# Wavelet Decomposition-Based Genomic Analysis of the Human Electrocardiogram

**DOI:** 10.64898/2026.05.20.26353725

**Authors:** Salma Zainana, Larissa Lauer, Tuomo Kiiskinen, Robert Tibshirani, Trevor Hastie, Euan A. Ashley, Jack W. O’Sullivan, Manuel A. Rivas

**Affiliations:** Department of Biomedical Data Science, Stanford University, Stanford, CA, USA, 94305; Department of Statistics, Stanford University, Stanford, CA, USA, 94305; Department of Medicine, Stanford University, Stanford, CA, USA, 94305; Institute for Computational & Mathematical Engineering, Stanford, CA, USA, 94305

## Abstract

The electrocardiogram (ECG) encodes the electrical activity of the heart across multiple timescales, yet standard clinical analysis collapses this rich signal into a handful of scalar measurements that discard most of the waveform’s structure. Whether the frequency signals lost in this reduction carry heritable biological information relevant to cardiovascular disease risk remains unclear. Here we decompose resting 12-lead ECGs from 47,052 White British UK Biobank participants into 84 frequency-specific energy features using Daubechies-6 wavelet analysis across 12 leads and 7 decomposition levels, and perform independent genome-wide association analyses on each feature. We identify 67 independent loci (p < 5 × 10^−8^) and refine these to 101 high-confidence causal variants (posterior inclusion probability > 0.80) through Bayesian fine-mapping; associated loci converge on genes governing cardiac conduction and myocardial integrity, including SCN5A, TTN, KCNQ1, and DSP, alongside less-characterized cardiomyopathy candidates. SNP-based heritability estimates range from 0.03 to 0.26, with the strongest signals in mid-frequency bands (D6–D4, ∼4–32 Hz) of Lead I and aVR, and strong inter-lead genetic correlations indicate a coordinated genetic architecture underlying the waveform. Integrating these features with FinnGen R12 cardiovascular phenotypes reveals genetic correlations reaching 0.56 with heart failure, driven predominantly by energy in the highest-frequency band (D1, 125–250 Hz) — a spectral range routinely filtered from clinical ECGs and previously regarded as acquisition noise. These results reframe the electrocardiogram as a multi-frequency genetic phenotype, expand the set of cardiac loci discoverable from ECG data, and implicate high-frequency cardiac electrical activity as an underexplored dimension of cardiovascular disease risk.

## Introduction

Electrocardiograms (ECGs) provide a unique, raw representation of cardiac physiology and pathophysiology. Recent genome-wide association studies (GWAS) of ECGs have primarily focused on standard parameters, which quantify the duration, amplitude, or timing of specific segments of the heartbeat cycle, such as the QRS complex [1], P wave [3], and T wave [2][25]. However, focusing solely on these traits may miss subtle morphological, spatial and frequency changes in ECG waveforms, including those not detectable to the human eye. Characterizing more granular and subtle waveforms provides the opportunity to identify unique genetic influences on granular ECG waveform morphology and frequency components. Incorporating time-frequency analysis has been shown to enhance the detection and classification of cardiovascular abnormalities by capturing both temporal and spectral characteristics of the ECG signal [3]. Although deep learning models can efficiently process raw ECG data across time and frequency domains, they often suffer from a lack of interpretability, functioning as ’black-box’ systems that obscure biological insights [4][5][27]. In contrast, a wavelet-based approach offers the advantage of generating interpretable features that can be directly linked to specific genetic loci through GWAS [5][28]. Such discoveries could deepen our understanding of genetic loci associated with ECG signals and advance our knowledge of cardiac diseases.

Here, we present a wavelet-based framework that derives frequency-specific representations of standard 10-second resting 12-lead ECGs from 47,052 UK Biobank individuals using discrete wavelet decomposition with Daubechies-6 (db6) wavelets [6]. This decomposition yields 84 energy features per individual (12 leads × 7 frequency levels), quantifying the contribution of distinct frequency bands—from slow baseline drift captured in the approximation level A6 (∼0–4 Hz) to rapid high-frequency activity in D1 (125–250 Hz)—to the overall cardiac waveform. These interpretable, physiologically grounded features serve as quantitative traits for genetic analysis. We perform 84 independent GWAS to identify loci associated with frequency- and lead-specific ECG variation, apply statistical fine-mapping using SuSiE-inf to resolve causal variants within associated loci, and use LD Score Regression to estimate SNP-based heritability and genetic correlations both across ECG features and with nine major cardiovascular disease phenotypes from the FinnGen R12 biobank [21]. This integrative approach enables the discovery of shared genetic architecture between ECG frequency components and cardiovascular disease risk, and reveals that high-frequency ECG bands, traditionally excluded from clinical interpretation, carry structured, heritable biological signals with direct relevance to heart disease.

## Materials and Methods

### Electrocardiogram signal

We performed our analysis on a standard 10-second resting 12-lead ECGs from 47,052 UK Biobank’s individuals[6]. Each recording is sampled at 500 Hz (5,000 samples per lead) and uses 10 electrodes placed on different locations on the human body to provide 12 different measurements of cardiac electrical signals (see **Figure 2)** .

The 12 leads consist of six limb leads and six chest leads. Of the six total limb leads, three are bipolar, standard limb leads and three are unipolar, augmented limb leads. The standard leads (I, II, and III) view the heart from the front of the body (the frontal plane) and are called bipolar because they are measurements of the potential difference between a positive electrode on one limb and a negative electrode on another limb. Lead I measures the difference between the positive electrode on the left arm and the negative electrode on the right arm. Lead II measures the difference between the positive electrode on the left leg and the negative electrode on the right arm. Lead III measures the difference between the positive electrode on the left leg and the negative electrode on the left arm. The augmented limb leads (aVL, aVR, and aVF) are readings from the same electrodes used for leads I through III, but only use a singular positive electrode, located on the left arm (aVL), the right arm (aVR), and the left leg (aVF). The six chest leads, labeled V1 through V6, like the augmented limb leads, are unipolar in that they are each measured from one singular positive electrode placed on the surface of the chest. The chest leads capture electrical activity from a horizontal plane, perpendicular to the frontal plane captured in the limb leads.

### Energy in ECG analysis

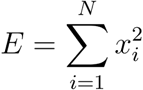

Quantifying cardiac electrical activity requires measuring signal energy, the cumulative intensity of deflections over time. For a discrete ECG signal *x* with N samples, energy is defined as the sum of squared amplitudes:

Squaring eliminates sign differences and emphasizes large deviations: tall R-waves dominate while baseline noise contributes minimally. This yields a scalar metric of heartbeat “strength.”

However, raw energy conflates distinct physiological processes. The QRS complex (∼10–40 Hz) carries high energy during ventricular contraction, while slow drift (∼0–3 Hz) accumulates energy slowly across seconds. To isolate these contributions, we must partition the signal’s total energy into frequency-specific bands—achieved through wavelet decomposition.

### Wavelet decomposition

Wavelet decomposition is a signal processing technique that breaks down complex data, such as an electrocardiogram (ECG) signal, into simpler components at different levels [8]. Applied to an ECG, this isolates rapid spikes from gradual shifts using filters that define the wavelets. In signal processing, a filter is a mathematical operation that selectively highlights certain waveforms from a series. The Haar wavelet, the simplest possible wavelet, illustrates this principle visually: it computes averages and differences of adjacent pairs using a step function. However, the Haar wavelet is discontinuous, producing blocky artifacts when analyzing smooth signals like ECGs. For this reason, smoother wavelet varieties are preferred.

Among these smoother alternatives, we selected Daubechies-6 (db6) because its scaling function resembles the QRS complex: a sharp central peak with smooth, oscillating tails (see **Figure 1**). This shape match preserves heartbeat morphology during analysis [8][10]. Concretely, db6 applies two complementary filters to the signal: a low-pass filter (*h*), averaging nearby points to capture slow trends; and a high-pass filter (*g*) computing weighted differences to isolate rapid changes. The filters are orthogonal thus they partition information without overlap or loss [7][9].

**Figure 1.**
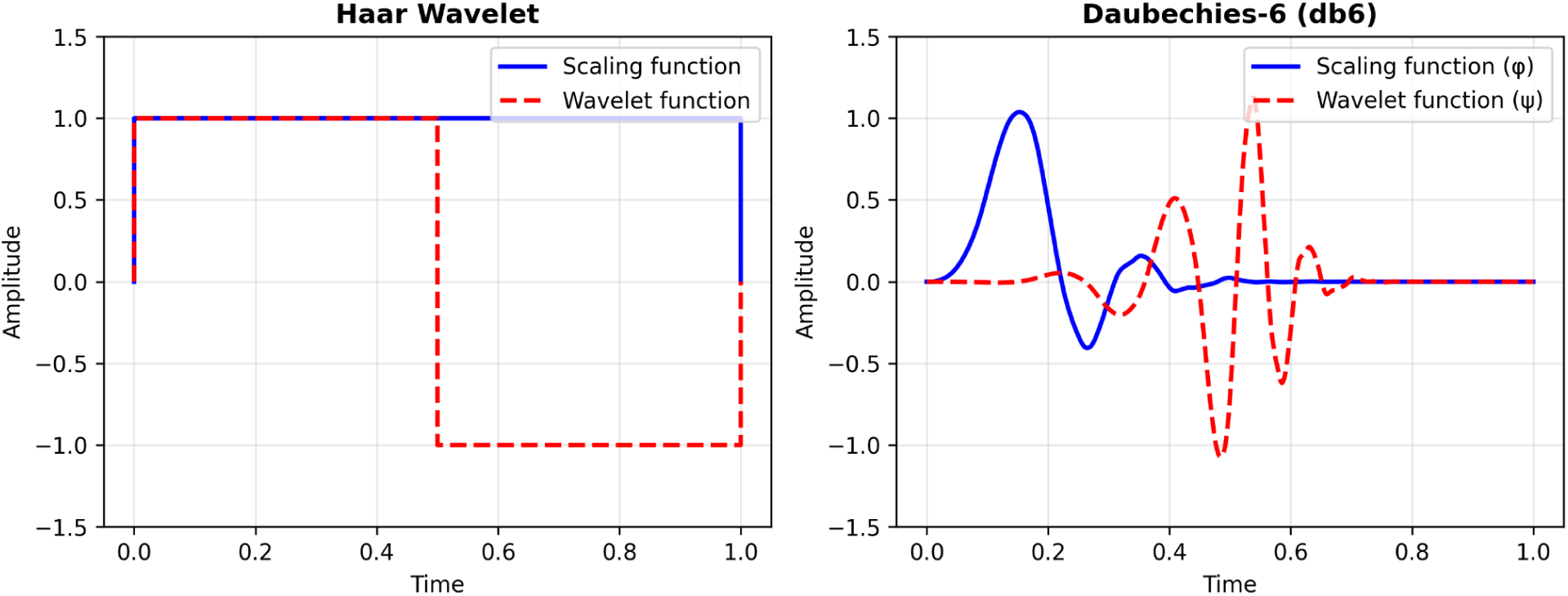
Wavelet basis functions for Haar and Daubechies-6 (db6). Each wavelet family is characterized by a scaling function φ (blue, solid) and a wavelet function ψ (red, dashed). These shapes are not chosen arbitrarily; they are the geometric fingerprint of the filter coefficients: φ emerges from repeatedly applying the low-pass filter *h* to itself, and ψ from the complementary high-pass filter *g* thus φ and ψ reveal what each filter is sensitive to. Left: the Haar scaling function is a flat step, discontinuous by construction, producing blocky artifacts when applied to smooth cardiac signals. Right: db6’s scaling function exhibits a sharp central peak with smooth, decaying oscillations that closely resemble the QRS complex morphology, making it geometrically well-suited to capture heartbeat energy in the approximation coefficients. X-axis: time; Y-axis: amplitude.

Each filter is applied through convolution—sliding a window across the signal and computing a weighted sum of neighboring points at each position. The low-pass filter *h* convolves with the signal to produce the approximation coefficients A, capturing low-frequency content; the high-pass filter *g* convolves with the same signal to produce the detail coefficients D, isolating high-frequency changes. Both outputs are then downsampled by keeping every other point, halving the length.

The algorithm proceeds iteratively. Starting from the full signal, it produces an approximation A1 (low frequencies, covering 0–125 Hz) and detail D1 (high frequencies, 125-250 Hz), then discards half of the sample [11]. Repeating this on A1: low-pass yields A2 (0–62.5 Hz), high-pass gives D2 (62.5–125 Hz), and downsample. After six levels, we also obtain A6 (0–3.91 Hz for slow trends) and details D3 (31.25–62.5 Hz), D4 (15.625–31.25 Hz), D5 (7.81–15.625 Hz), D6 (3.91–7.81 Hz) [10][11]. We do not need to keep intermediate approximations A1 through A5 because their information is captured in the subsequent details (D2 through D6) and the final A6 [9]. The original ECG can be perfectly rebuilt by summing these pieces as energy is fully preserved throughout (see **Figure 2)**. All wavelet decompositions were computed in Python using the PyWavelets package [32].

**Figure 2.**
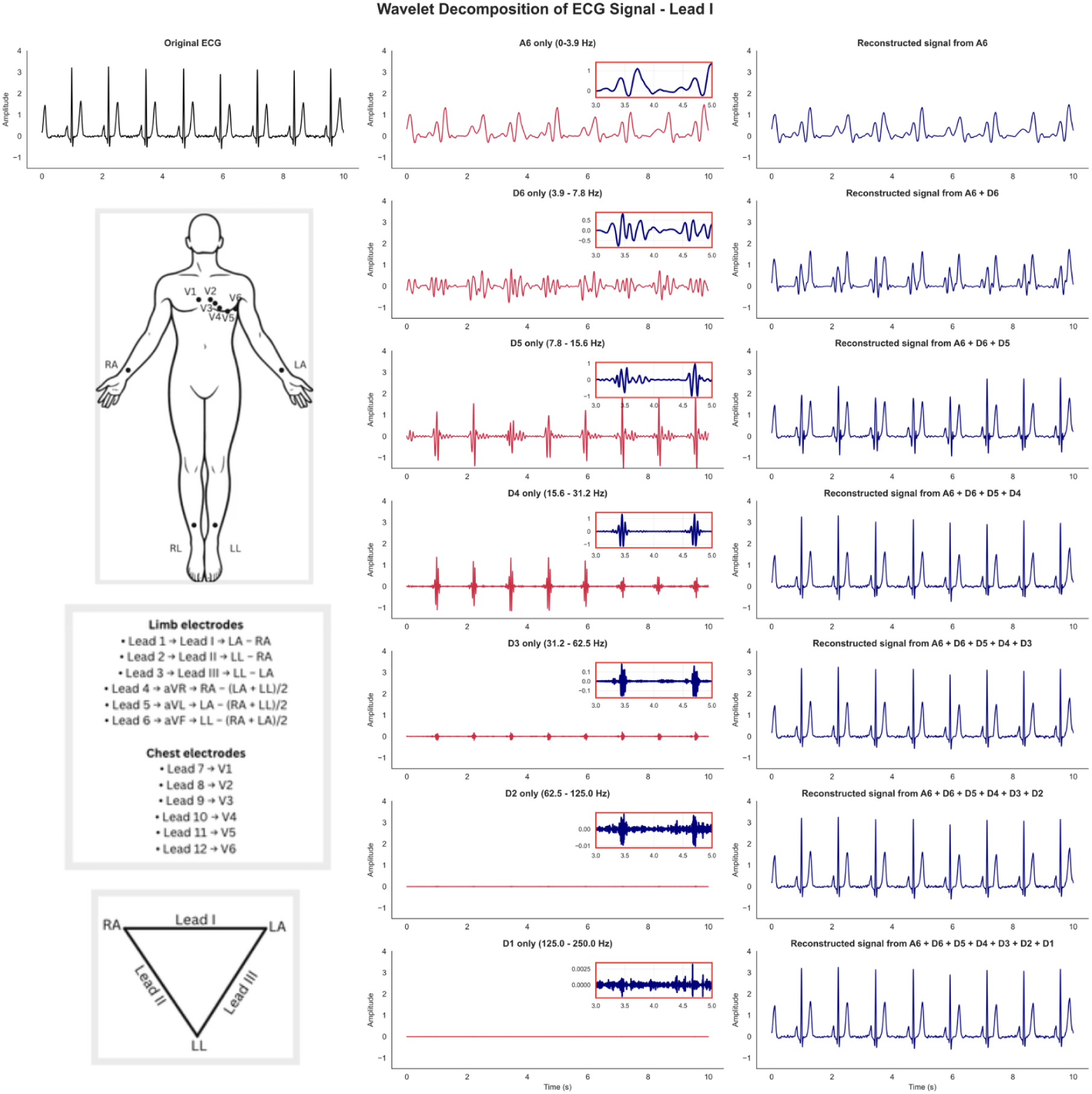
Wavelet decomposition of a single-lead ECG signal and overview of electrodes placements. The ECG signal (Lead I) is decomposed using the Daubechies-6 (db6) wavelet at 6 levels. **Column 1** shows the original ECG and the standard electrode configuration used to record 12-lead ECGs. **Column 2** displays the reconstructed signal of each individual level (A6, D6–D1), obtained by inverse wavelet transform with all other coefficients set to zero, and including a zoomed inset highlighting a 2-second segment. **Column 3** illustrates reconstruction of the signal by cumulatively adding wavelet levels starting from the approximation A6 (lowest frequency) and progressively including detail coefficients D6 to D1 (increasing frequencies). X-axis: time; Y-axis: amplitude.

### Energy distribution across levels

Following decomposition, we calculate energy at each level:

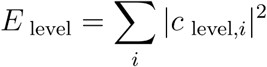

where *c_level_*, is the *i^th^* coefficient at the chosen *level* (e.g., A6, D6, D5, …,D1) .

For multi-lead ECG data, we compute this energy separately for each lead and each level using the corresponding coefficients. This calculation measures the energy contribution of each frequency band per lead without emphasizing any single coefficient. Parseval’s theorem for the discrete wavelet transform (DWT) states that the signal’s energy stays identical before and after decomposition. As a result of this energy-preserving property, no details vanish during the breakdown, and we can rebuild the ECG from its coefficients by reversing the steps as shown in **Figure 2**.

### ECG phenotype preparation

Resting 12-lead ECGs were obtained from UK Biobank field 20205, which provides raw waveform data digitised at 500 Hz over a 10-second recording window. Each of the twelve leads was processed independently. Raw signals were first denoised with a notch filter to suppress powerline interference, and were then submitted to db6 wavelet decomposition as described above. Recordings in which any lead contained a decomposition level with zero coefficient inputs, indicating a flat or absent segment, were excluded from downstream analysis. For each retained lead, signal energy was computed separately at each of the seven decomposition levels (A6, D1–D6) yielding 84 lead-by-level energy phenotypes (12 leads × 7 decomposition levels). Each of the 84 phenotypes was rank-based inverse-normal transformed (INT) prior to genome-wide association testing.

### Genotyping, imputation, and sample quality control

Genotyping, imputation, and quality control of UK Biobank samples have been described in detail by Bycroft et al. [6] and are summarised here. Participants were genotyped on the UK Biobank Axiom array. We restricted the analysis to participants of self-reported White British ancestry (data field 21000), confirmed by principal component analysis, and to unrelated individuals as defined by the UK Biobank kinship file. After applying these genotypic filters together with the ECG flat-signal exclusion described above, the final GWAS sample comprised n = 47,052 individuals.

### Genome-wide association analysis

We performed an independent genome-wide association study (GWAS) for each of the 84 wavelet energy phenotypes in the n=47,052 unrelated individuals retained after the genotypic and ECG quality control mentioned above. Using PLINK2, we analyzed SNPs across chromosomes 1-22, adjusting for age, sex, and population structure (via the first 10 principal components) to account for potential confounders. For every lead–level combination, we fitted an additive linear model regressing the inverse-normal-transformed (quantile-normalized) energy phenotype on allele dosage at each variant, with age, sex, and the first ten genetic principal components included as covariates. The genome-wide significance threshold for each individual GWAS was set at the conventional p = 5 × 10^−8^. To account for multiple testing across the 84 correlated traits, we estimated the effective number of independent phenotypes (M_eff) empirically via a permutation procedure adapted from [20] . We shuffled phenotype IIDs once across all 84 traits simultaneously, preserving inter-trait correlations while breaking the genotype–phenotype link, and ran 84 permuted GWAS on an LD-pruned SNP set (451,310 SNPs, r² < 0.1). For each SNP, the minimum p-value across the 84 null GWAS was retained; the 5th percentile (α_FWER = 0.05) of this null distribution defines the per-trait empirical significance threshold maintaining a 5 % family-wise error rate across the 84 traits. M_eff was derived from p via the Šidák relation M_eff = log(1 − α_FWER_) / log(1 − p). The phenotype-corrected genome-wide significance threshold was then set to α_GW_ / M_eff, with α_GW_ = 5 × 10^−8^ the conventional single-GWAS threshold [20].

### Statistical fine-mapping

We performed statistical fine-mapping for each of the 84 genome-wide association studies (GWAS) of wavelet-energy traits using a multi-stage Bayesian framework, SuSiE-inf [14] . Genome-wide significant variants (p < 5 × 10^−8^) were first identified and pruned by LD–based clumping (r² < 0.1, window = 500 kb) to define independent lead loci. For each lead SNP, we extracted a ±500 kb window centered on the index variant and retained significant or LD-correlated variants within this interval. Fine-mapping was then performed using the SuSiE-inf algorithm (moments method; L = 5; maxiter = 500), applied to the vector of z-scores and the corresponding LD matrix. This yielded posterior inclusion probabilities (PIPs) and 95% credible sets for each locus.

### SNP-based heritability estimation

SNP-based heritability (h²) was estimated for each of the 84 wavelet energy phenotypes independently using LDSC v1.0.1 [12], with European LD scores from the 1000 Genomes Project [13] computed on HapMap3 variants as a reference panel. For each phenotype, h² was obtained from the slope of the regression of GWAS χ² statistics on per-SNP LD scores.

### Genetic correlation analysis

Genetic correlations (rg) were estimated within the LDSC framework via cross-trait regression (--rg flag) [12], using European LD scores from the 1000 Genomes Project [13] as reference panel. Three sets of correlations were computed. First, all pairwise rg among the 84 wavelet energy phenotypes were estimated; the Lead I × aVR pair is reported in detail in the main text as a representative example, with the full pairwise matrix presented in Supplementary Figures 3. Second, rg between each of the 84 phenotypes and seven I9-prefix cardiovascular disease phenotypes from the FinnGen R12 release [21] were computed and reported at a significance threshold of p < 0.005. Third, the rg between each phenotype and body mass index were estimated using publicly available BMI GWAS summary statistics generated in-house with PLINK2 on the same unrelated White British UK Biobank cohort used for the wavelet-energy GWAS, applying identical covariate adjustment (age, sex, PC1–10) and quantile-normalized BMI as the outcome. Partial genetic correlations conditioning on BMI were computed analytically as

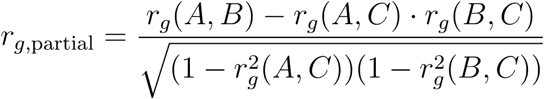

where C denotes BMI.

### Gene and pathway enrichment analysis

Lead SNPs identified from per-GWAS LD clumping were mapped to genes using FUMA SNP2GENE [30], with positional mapping based on genome build GRCh38. The resulting gene list was submitted to Enrichr [31] for over-representation testing against three curated libraries: Jensen DISEASES Curated 2025, CellMarker 2024, and Reactome Pathways 2024. Enrichment significance was assessed using Enrichr’s standard Fisher’s exact test.

### Sensitivity analyses

#### Reverse causation

To assess whether the genetic correlation between V1 D1 energy and heart failure and coronary heart disease could be attributed to reverse causation, whereby having an established heart condition changes the ECG signal, which then artificially inflates the genetic correlation, we performed a sensitivity analysis restricted to participants free of prevalent heart failure (HC299), cardiomyopathy (HC414), and myocardial infarction (HC326), yielding n = 46,108 phenotypes remaining after exclusions.

GWAS were re-run for all 12 D1 leads in this disease-free subset, and LDSC genetic correlations were re-estimated against FinnGen R12 cardiovascular phenotypes.

#### BMI independence

To further quantify the independence of D1 cardiac signal from adiposity genetics, we performed two complimentary sensitivity analyses. First, we repeated all 12 D1 GWAS with BMI included as an additional covariate alongside age, sex, and PC1–10, and compared effect sizes at genome-wide significant loci between the base and BMI-adjusted models. Second, we computed the partial genetic correlation between D1 wavelet energy and heart failure conditioning on BMI analytically, using the formula given in Genetic correlation analysis above.

### Positive control validation

To validate that wavelet-derived energy features recapitulate known cardiac electrophysiology, and to confirm their frequency-band specificity prior to interpreting novel findings, we tested their association with five standard machine-reported ECG metrics available in UK Biobank: QRS duration (field 12340), QT interval (field 22331), P-wave duration (field 12338), heart rate (field 22426), and R-wave axis (field 22336). For each of the 84 wavelet energy features, we fitted a linear regression model of the form: ECG metric ∼ wavelet energy + age + sex + PC1–PC10. Both the ECG metric and wavelet energy feature were rank-based inverse-normal transformed (INT) prior to regression, such that regression coefficients (β) are interpretable as partial correlations, consistent with the INT normalization used in the main GWAS. Analyses were restricted to White British individuals (n ≈ 11,800–17,200 depending on metric availability).

## Results

We analyzed mean energy across leads and levels (see **Figure 3**). As expected from cardiac electrophysiology, the majority of signal energy is concentrated in the low- and mid-frequency components (A6–D4), which correspond to large-scale deflections such as the P wave, QRS complex, and T wave [10]. Yet a consistent minority of energy remained in the high-frequency detail bands (D2–D1), historically regarded as noise within conventional clinical recordings. These components, especially D1 (125–250 Hz), are filtered out or visually imperceptible on standard ECGs but are preserved in raw trace data and wavelet coefficients.

**Figure 3.**
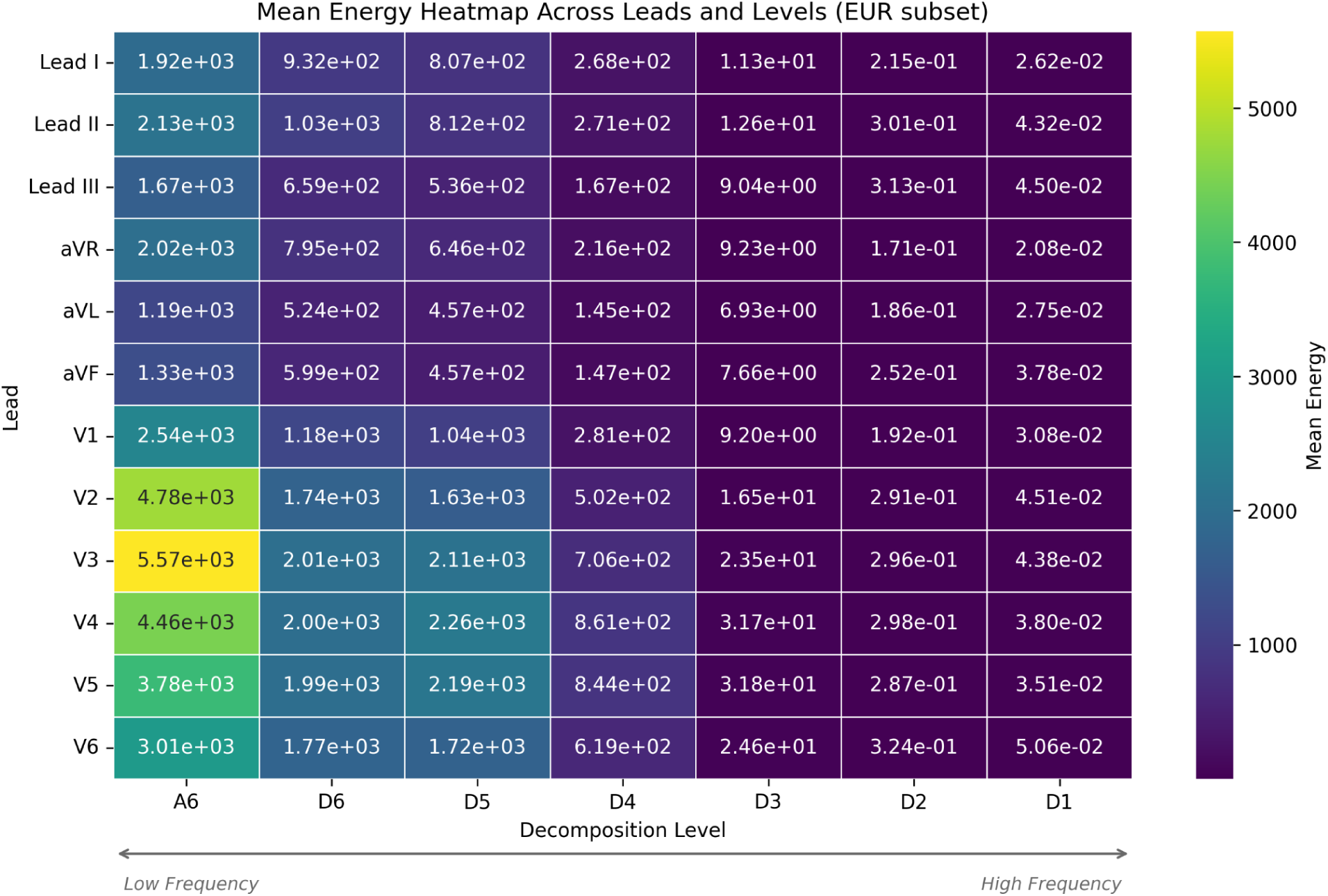
Heatmap of energy features across ECG leads and decomposition level. X-axis: decomposition levels; Y-axis: 12 leads. Each row represents a lead and each column represents a decomposition level (detail or approximation). The color intensity corresponds to the magnitude of the energy at that lead and level.

For each of the 84 lead–level combinations we ran an independent genome-wide association analysis (Methods, Genome-wide association analysis; Supplementary Figures 1). To synthesize results across the 84 analyses, we aggregated the minimum p-value for each SNP across all lead–level combinations as a visual summary of the overall association landscape (**Figure 4A**); because this aggregation takes the minimum across 84 tests, it inflates type I error relative to any individual GWAS and is therefore an exploratory visualization rather than a formal discovery procedure. Independent loci were defined from by applying LD clumping to this combined GWAS, yielding 67 genome-wide significant loci at the conventional genome-wide significance threshold (p < 5 × 10^−8^). Applying the empirical permutation procedure described in Methods (Genome-wide association analysis) to the 84 correlated phenotypes, we obtained a per-trait empirical threshold of p =1.21×10^−3^, corresponding to M_eff = 42.4 effective independent traits and a corresponding corrected genome-wide threshold is 5×10^−8^ / 42.4 = 1.18×10^−9^. Of the 67 lead variants, 56 exceed this corrected threshold, confirming that our findings are robust to multiple testing correction [20]. **Figure 4B** shows the 67 independent lead variants derived from the clumping procedure, as well as the 56 loci derived from the corrected threshold.

**Figure 4A.**
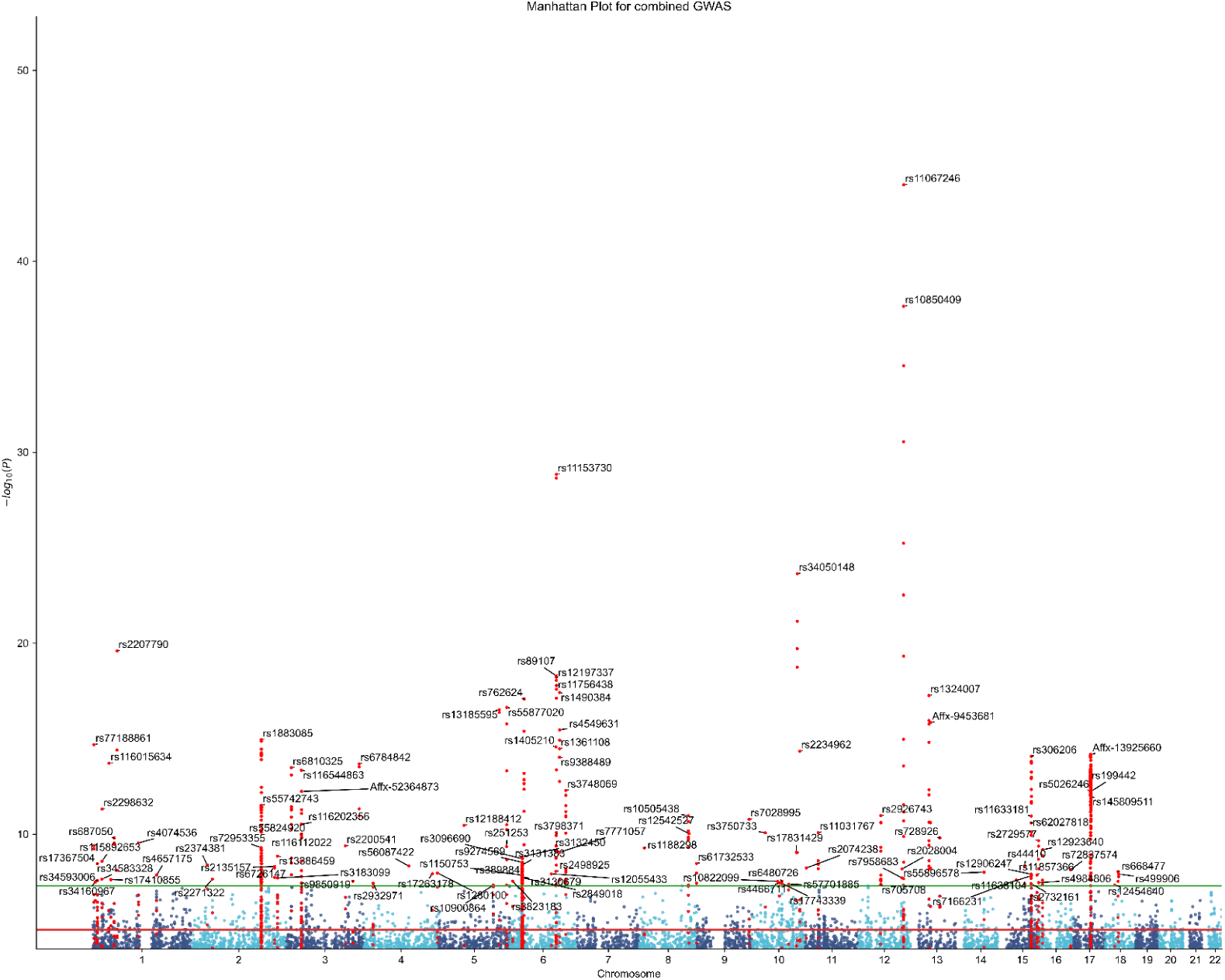
Manhattan plot displaying genome-wide association results aggregated across 84 waveform energy phenotypes (12 ECG leads × 7 wavelet decomposition levels), each analyzed in a separate GWAS. Each point represents a SNP plotted by chromosomal position (x-axis) against its association significance expressed as –log₁₀(p-value) (y-axis). P-values were derived from ordinary least squares linear regression (PLINK2), testing the additive effect of allele dosage on quantile-normalized waveform energy at each lead–level combination, with adjustment for age, sex, and 10 genetic principal components. Horizontal lines indicate suggestive (p = 1×10^−5^, red) and genome-wide significance thresholds (p = 5×10^−8^, green).

**Figure 4B.**
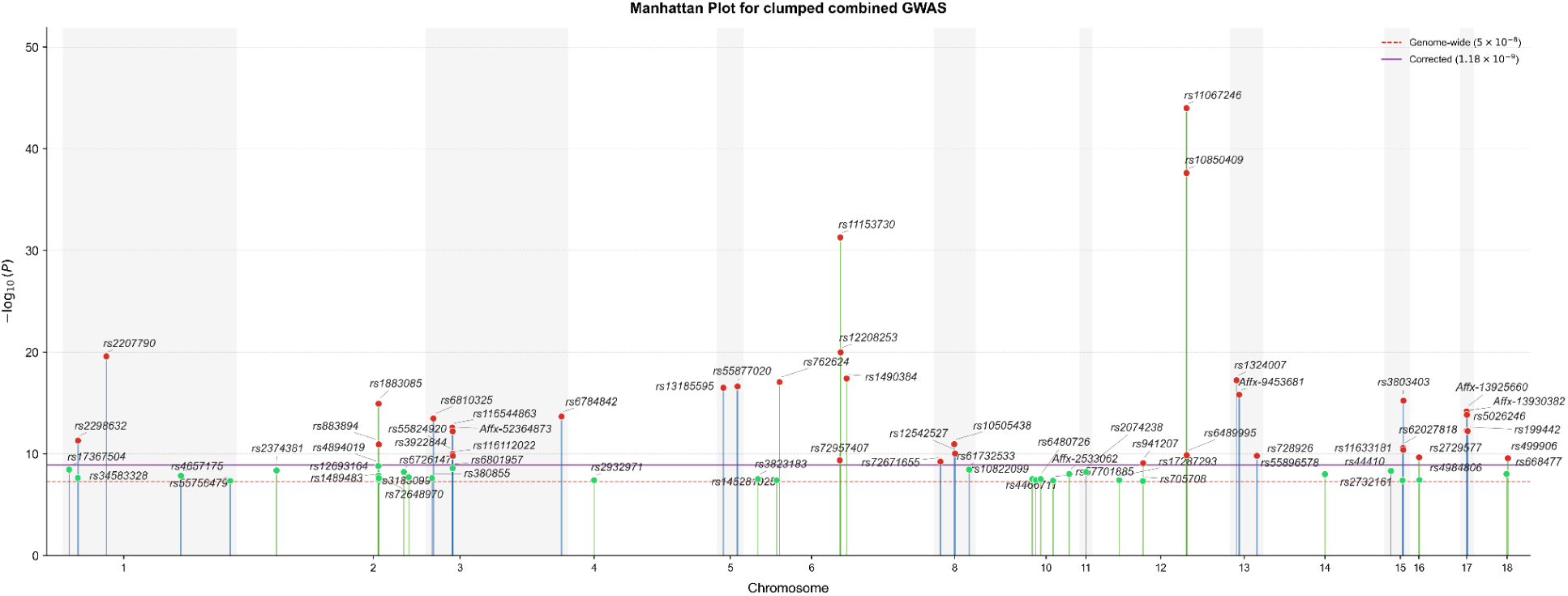
Manhattan plot of 67 independent lead SNPs identified after LD clumping (r² < 0.1, 500 kb window) applied to each of the 84 GWAS separately. Each point represents the index variant with the strongest association within each correlated SNP set, plotted by chromosomal position (x-axis) against –log₁₀(p-value) (y-axis). SNPs are colored by significance tier: red for variants surpassing the Bonferroni-corrected threshold (p = 1.18 × 10^−8^), and green for variants meeting the conventional genome-wide significance threshold (p = 5 × 10^−8^).

As a positive control (Methods, Positive control validation), we tested whether the 84 wavelet energy features recapitulate known cardiac electrophysiology. Mid-frequency wavelet bands (D3–D6, 7–62 Hz) showed consistent, highly significant, and physiologically interpretable associations with all five standard ECG metrics (see **Figure 10**). QRS duration was most strongly associated with D3 energy in limb leads (β up to 0.25, p < 10^−^200), consistent with ventricular depolarization generating maximal power in this frequency range. QT interval showed widespread negative associations with D3–D5 energy across all leads (β ≈ −0.15 to −0.28, p < 10^−50^), reflecting the inverse relationship between repolarization duration and mid-frequency power concentration. Heart rate showed positive associations with D3–D4 energy across all 12 leads (β ≈ 0.15–0.26, p < 10^−^100), consistent with faster heart rates concentrating more energy per unit time in mid-frequency bands. P-wave duration showed significant associations predominantly with D2 energy in limb leads (β ≈ −0.17, p < 10^−100^), reflecting atrial depolarization activity in the corresponding frequency range. R-wave axis showed the strongest associations in the dataset, with large opposing effects between Lead I (β ≈ −0.56, p < 10^−300^) and Lead II (β ≈ +0.45, p < 10^−300^) at mid-frequency bands, directly recapitulating the vector nature of cardiac depolarization along orthogonal leads. Critically, the highest-frequency band D1 (125–250 Hz) showed markedly attenuated associations with all five standard ECG metrics across all 12 leads (|β| < 0.13 across all lead-metric combinations), with substantially weaker significance compared to mid-frequency bands.

The estimated SNP-based heritability (h²) for the 84 energy features ranges from ∼0.03 to 0.26 (see **Figure 5**, **Supplementary Figure 2**), with several features reaching h² ∼0.2-0.25, comparable to levels previously associated with meaningful genetic contribution [15]. Leads I and aVR exhibit higher heritability in mid-detail levels (D6–D4, h² ∼0.20-0.25) than leads placed directly over the cardiac surface, consistent with their alignment along the heart’s normal electrical conduction axis. Precordial leads V5 and V6 exhibit elevated h² (∼0.13–0.17) in the D6 level, aligning with genetic contributions to left ventricular repolarization. Finer details (D2–D1) show lower h² (∼0.05–0.10) across leads, while the A6 level varies (e.g., higher in aVR at 0.17), suggesting genetic bases for overall waveform shape.

**Figure 5.**
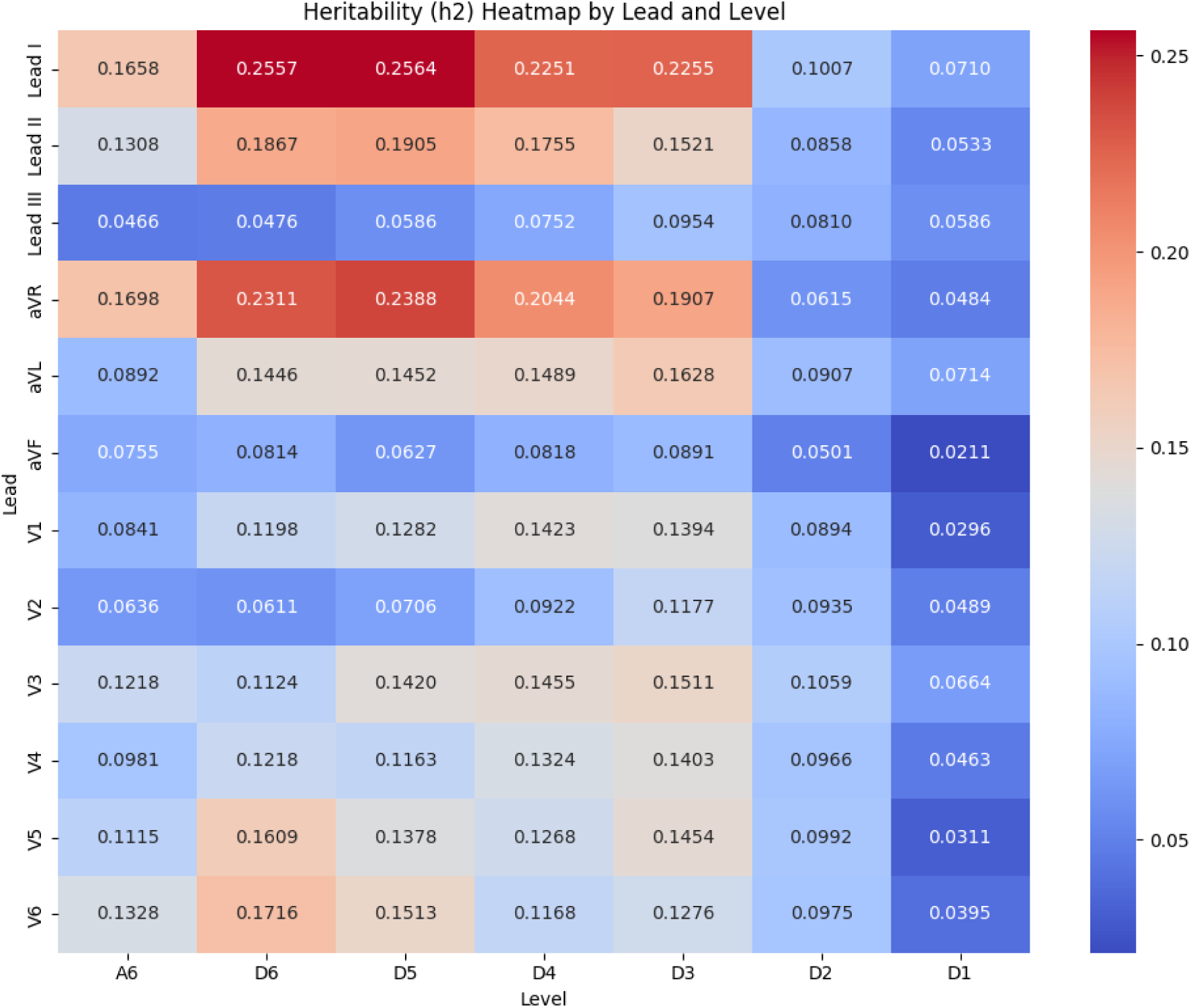
Heatmap of the heritability (h²) estimates of energy features across ECG leads and decomposition level. X-axis: decomposition levels; Y-axis: 12 leads. Each cell shows the SNP-heritability estimated using LD Score Regression (--h2) with European LD scores from the 1000 Genomes Project [13]. For each lead-level energy phenotype, GWAS summary statistics were regressed on LD scores, and h2 was computed from the slope of this regression.

Given their elevated heritability in mid-detail levels (h² ∼0.20–0.25, see **Figure 5**), we focused on the genetic correlation between Lead I and aVR (see **Figure 6**). Both leads show strong positive rg scores in mid-levels (rg ∼0.5–0.8) suggesting shared associated variants for mid-frequency components. Genetic correlations across all lead–level combinations are shown in **Supplementary Figures 3**.

**Figure 6.**
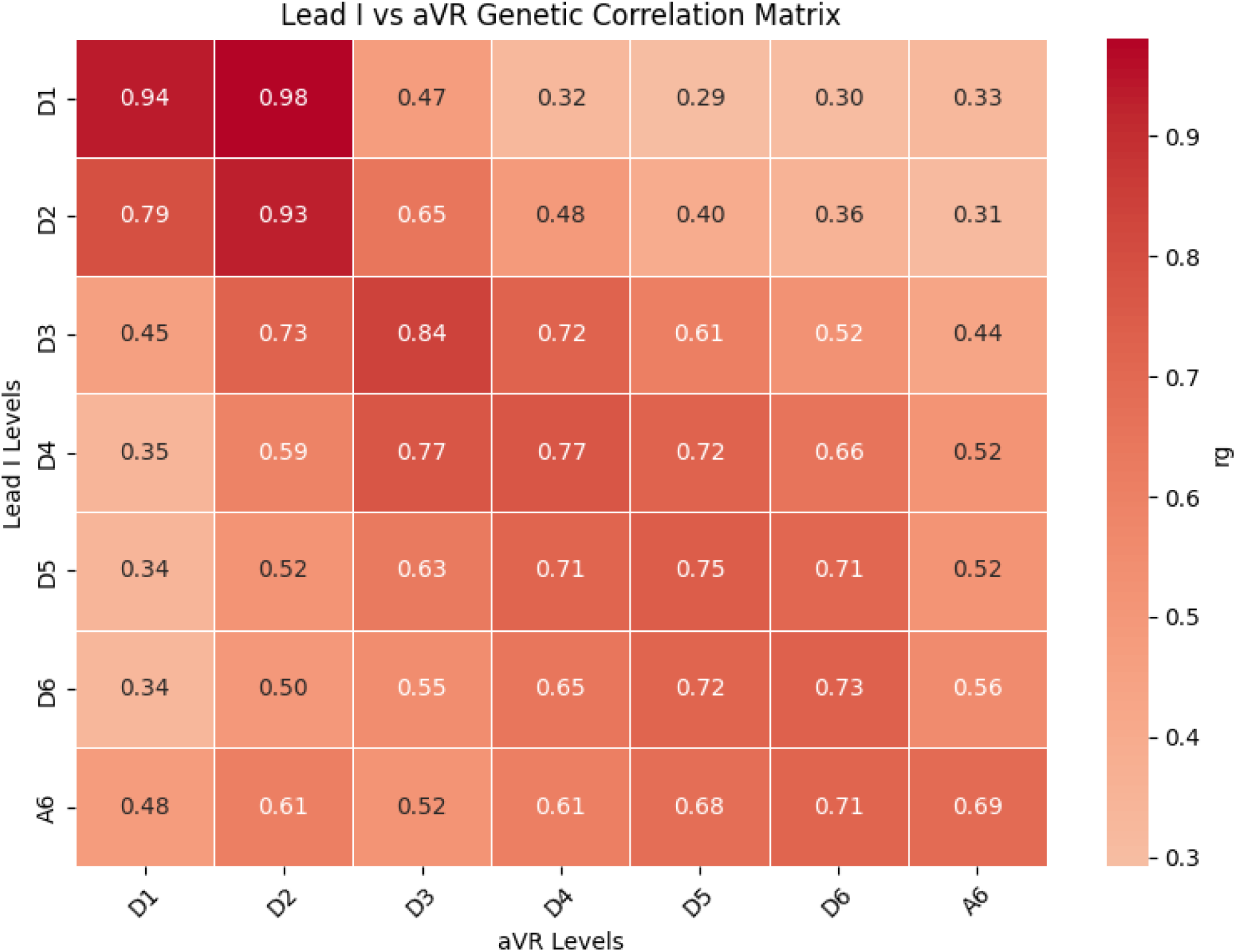
Heatmap of genetic correlations (rg) for energy features between Leads I and aVR across wavelet decomposition levels. X-axis: energy levels for aVR lead; Y-axis: energy levels for Lead I. Each cell shows the genetic correlation estimated using LD Score Regression (--rg) with European LD scores from the 1000 Genomes Project reference panel. For each pair of lead–level energy phenotypes, GWAS summary statistics were jointly analyzed by regressing the product of SNP z-scores across traits on LD scores, and the genetic correlation (rg) was derived from the slope of this cross-trait regression, scaled by the square roots of the corresponding SNP-heritability estimates.

To validate and contextualize our ECG-derived energy features within broader clinical phenotypes, we computed LDSC genetic correlations with seven I9 cardiovascular disease phenotypes from the FinnGen R12 release (Methods) and report associations at p<0.005 (see **Figure 7**). Heart failure and coronary heart disease showed the broadest and strongest genetic links, with correlation values reaching a peak of 0.5591 for V1 at high-frequency detail level D1 (p = 0.0039), followed by notable genetic correlations such as 0.3760 for V1 at low-frequency approximation A6 (p = 0.0001), 0.3322 for V1 at mid-frequency detail D5 (p = 3.8 × 10^−5^), and 0.3164 for V1 at lower-frequency detail D6 (p = 0.0002). These were accompanied by consistent patterns in Lead I and aVL across various frequency levels D1–D6 and A6 (for example, Lead I at D4: rg = 0.22, p = 9.4 × 10^−5^; aVL at D5: rg = 0.24, p = 0.0027). Coronary atherosclerosis showed strong genetic links primarily in right precordial and lateral leads, with correlation values reaching a peak of 0.3894 for V1 at high-frequency detail level D1 (p = 0.0027), followed by notable genetic correlations such as 0.3778 for V6 at D1 (p = 0.0004), 0.2967 for V1 at low-frequency approximation A6 (p = 4.1 × 10^−5^), and 0.2292 for V1 at mid-high frequency detail D2 (p = 0.0003). Additional patterns appeared in aVR and Lead II, such as aVR at mid-frequency detail D5 (rg=0.15, p = 0.0004) and Lead II at D1 (rg= 0.25, p = 0.004). Left bundle branch block revealed focused genetic links with aVL, peaking at 0.4233 for low-frequency approximation A6 (p = 0.0026) and including 0.3744 for lower-frequency detail D6 (p = 0.0021).

**Figure 7.**
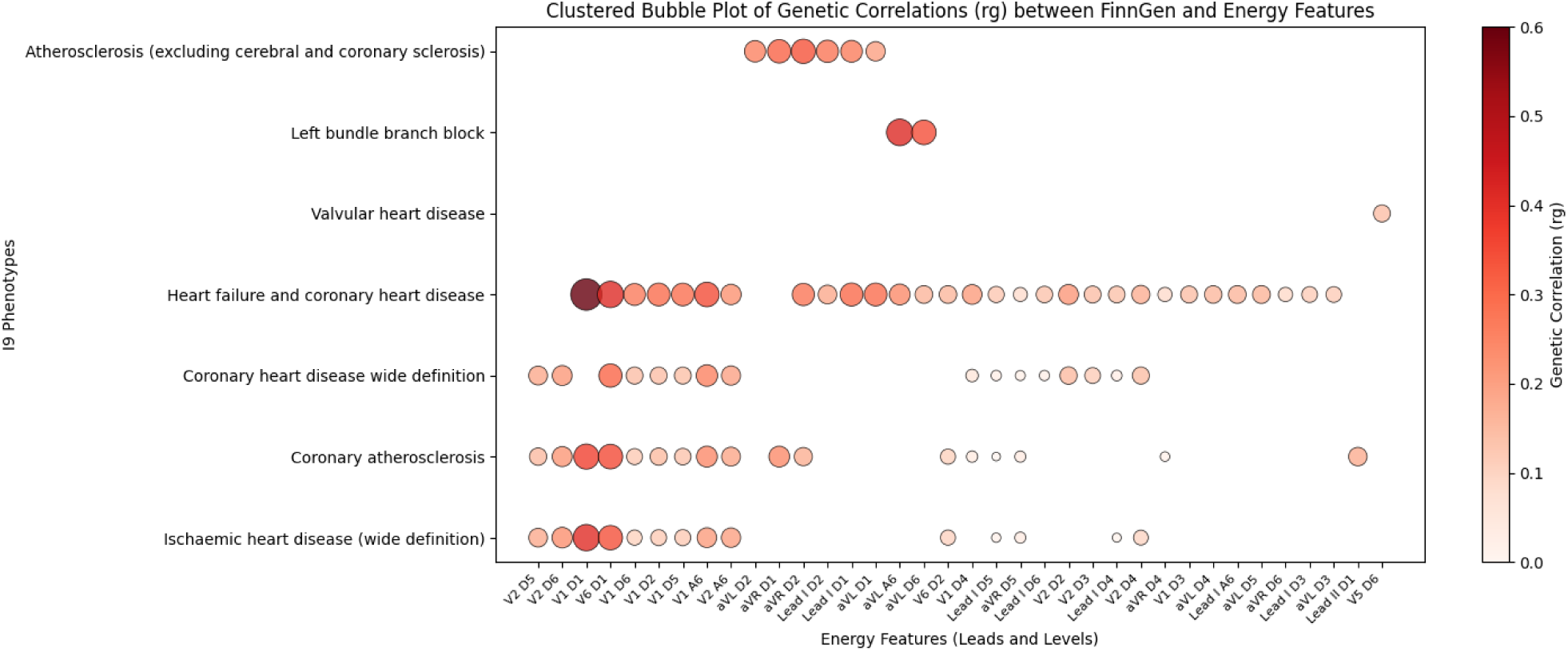
Clustered bubble plot of genetic correlations (rg) between 84 ECG energy features (12 leads × 6 detail levels + 1 approximation level) and 7 FinnGen cardiovascular phenotypes. Each cell shows the genetic correlation estimated using LD Score Regression. X-axis: energy features ; Y-axis: FinnGenn diseases. Only associations with p < 0.005 are shown. Larger and darker bubbles indicate stronger genetic correlations. Lead and level combinations such as Lead I D6 and V1 D5 exhibit the strongest associations, particularly for heart failure and coronary heart disease. These spatial patterns reflect disease-specific electrophysiological and genetic signatures across the 12-lead ECG.

As a sensitivity analysis for reverse causation (Methods, Sensitivity analyses), LDSC genetic correlations against FinnGen R12 cardiovascular phenotypes were re-estimated in a subset of UK Biobank participants free of prevalent cardiac disease at baseline. The genetic correlation between V1 D1 energy and heart failure and coronary heart disease was attenuated from rg = 0.56 (p = 0.0039) in the full sample to rg = 0.38 (p = 0.0026) in the disease-free subset, remaining statistically significant. This attenuation pattern was consistent across other D1 leads showing significant correlations in the primary analysis **(Supplementary Table 4).**

To assess the biological relevance of the loci identified in the combined GWAS, we performed pathway and gene ontology enrichment analyses (Methods, Gene and pathway enrichment analysis) on the 110 genes mapped from clumped leading SNPs (see **Supplementary Table 2**). We find significant enrichment in pathways and disease terms related to cardiac electrophysiology and muscle function. In Reactome Pathways 2024, top enriched terms included “Muscle Contraction” (p =1.91×10^−6^) and “Cardiac Conduction” (p = 7.6×10^−6^) [25]. In Jensen DISEASES (see **Figure 8**), prominent enrichments were found for “Cardiomyopathy” (p = 2.66 × 10^−12^) and “Heart Disease” (p = 3.3 × 10^−13^). Additionally, cell type enrichment analysis using CellMarker 2024 revealed significant overrepresentation in cardiac-related cell types, with the strongest signals in “Cardiomyocyte Heart Mouse” (p = 1.47 × 10^−4^) and “Cardiomyocyte Heart Human” (p = 3.84 × 10^−4^) (see **Figure 8**, **Supplementary Table 3**). **Figure 8** highlights specific genes driving these enrichments, such as ALPK3, ANKRD1, CTNNA3, DSP, FHOD3, KCNQ1, NKX2-5, NPPA, PLN, SCN10A, SCN5A, TTN, VCL, and ZFPM2, associated with ontology terms like cardiomyopathy, dilated cardiomyopathy, and arrhythmogenic right ventricular cardiomyopathy, as well as cell types including cardiomyocytes and cardiac progenitors.

**Figure 8.**
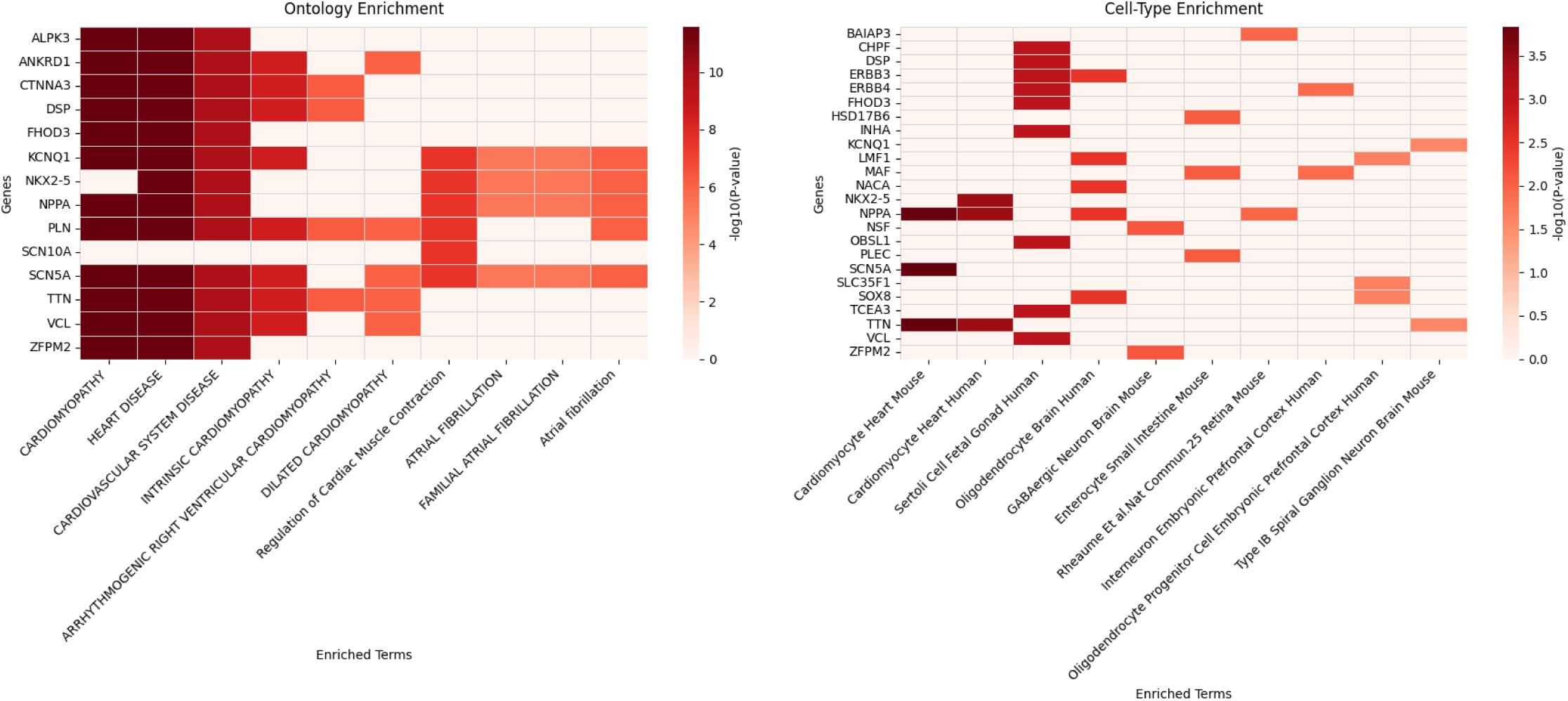
Gene enrichment analysis of 110 genes mapped from the clumped leading SNPs via FUMA snps2genes, submitted to Enrichr against the **Jensen DISEASES Curated 2025** (ontology enrichment, left) and **CellMarker 2024** (cell-type enrichment, right) libraries. X-axis: enriched terms; Y-axis: genes. For each term, enrichment significance was assessed by testing whether the overlap between submitted genes and the term’s gene set exceeded what would be expected by chance. Color intensity reflects −–log₁₀(p-value), with darker red indicating greater statistical enrichment. Left: enriched disease ontology terms highlight convergent genetic associations with cardiomyopathy, arrhythmia, and related cardiac pathologies. Right: cell-type enrichment reveals preferential expression of associated genes in cardiomyocytes, consistent with a cardiac-specific genetic architecture.

To characterize the relationship between our ECG energy features and adiposity, and to assess whether D1 signal reflects skeletal muscle EMG artifact modulated by subcutaneous fat, we computed genetic correlations between all 84 wavelet traits and BMI (Methods; see **Figure 9**). The resulting pattern is spatially coherent and maps onto the canonical ECG signature of obesity-related leftward axis deviation: left-oriented limb leads (Lead I, aVL) show strongly positive rg with BMI across mid-frequency bands (Lead I D1: rg = 0.40, p = 9.7×10^−11^; aVL D1: rg = 0.33, p = 3.7×10^−7^), inferior leads show strongly negative to near zero rg (aVF D1: rg = −0.01, p = 0.90; Lead II D1: rg = 0.02, p = 0.71), and Lead III was near zero across all levels. Right precordial leads (V1–V2) were weakly positive while left precordial leads (V3–V6) were moderately negative, consistent with horizontal plane posterior displacement of the cardiac mass from diaphragmatic elevation. Notably, the axis-deviation signal is concentrated in mid-frequency bands (D6–D3) and collapses at D1 for the inferior leads most sensitive to axis shift (aVF: rg = −0.47 at D5 → rg = −0.01 at D1). Two complementary sensitivity analyses (Methods, Sensitivity analyses; **Supplementary Analysis 2**) further quantified the independence of the D1 cardiac signal from adiposity genetics. After BMI conditioning, the lead SNP in the SCN5A region (Affx-89018181), showed β attenuation of less than 2% across all 12 D1 leads, and genetic correlations between BMI-adjusted D1 energy and heart failure remained significant (V1 D1: rg = 0.36, p = 0.0045). Second, the partial rg remained substantial across all leads with significant D1–HF associations.

**Figure 9.**
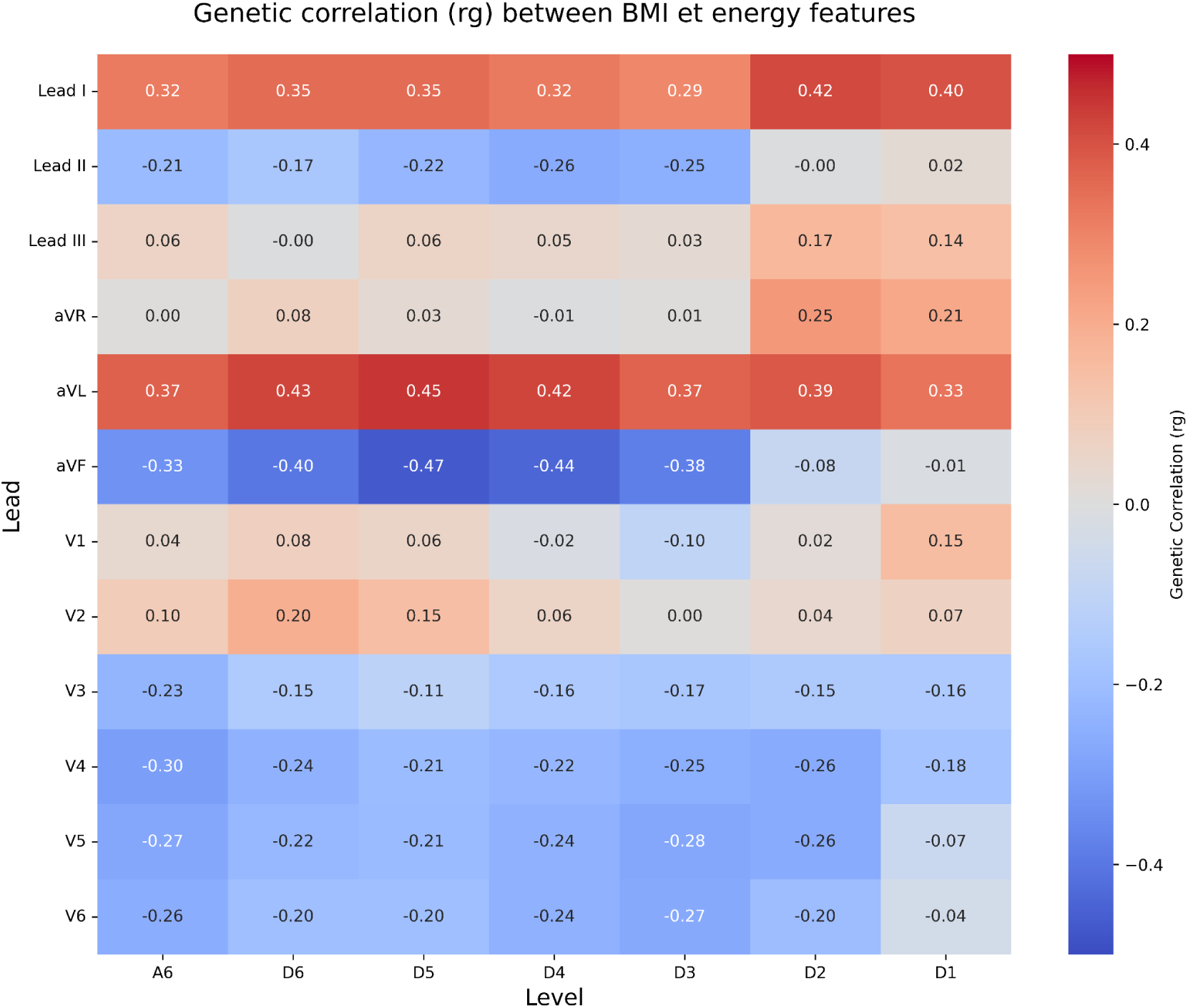
Heatmap of genetic correlations (rg) between 84 ECG energy features (12 leads × 6 detail levels + 1 approximation level) and BMI. Each cell shows the genetic correlation estimated using LD Score Regression. X-axis: decomposition levels (A6, D6–D1); Y-axis: 12 ECG leads. Color intensity reflects the magnitude and direction of rg, with red indicating positive correlations and blue indicating negative correlations.

**Figure 10.**
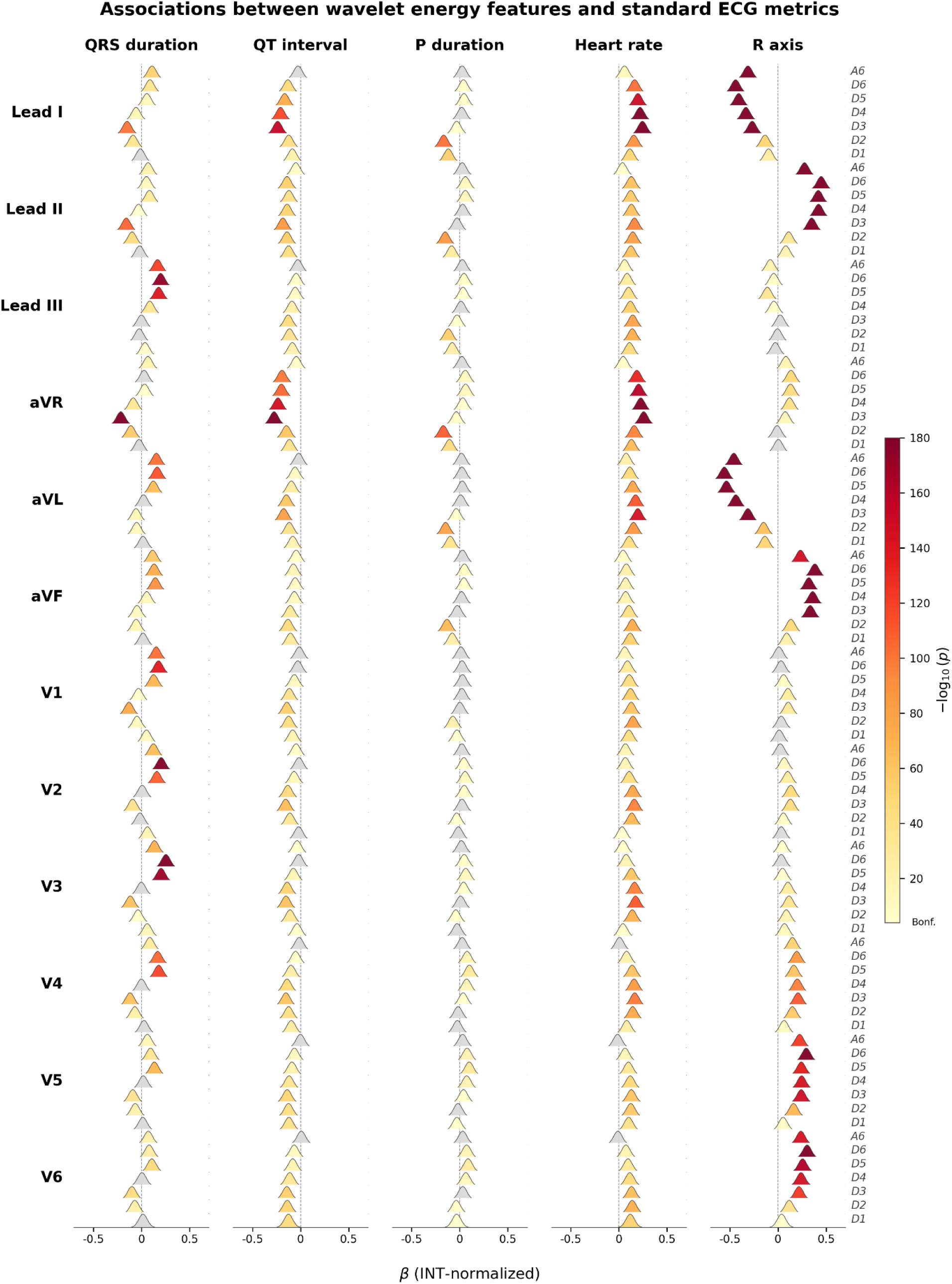
Forest plot of regression coefficients (β) for the 84 ECG wavelet energy features on five standard UK Biobank machine-reported ECG metrics (QRS duration, QT interval, P-wave duration, heart rate, R-wave axis). Each cell shows β as a Gaussian density centred on the point estimate with width proportional to the standard error; distributions not overlapping zero indicate effects reliably distinct from zero. X-axis: β. Y-axis: 12 ECG leads, each containing 7 distributions for wavelet levels A6, D6–D1. Colour intensity reflects –log₁₀(p-value), with bright yellow indicating high significance and dark blue weak association.

Statistical fine-mapping of the 84 wavelet-energy GWAS, conducted using SuSiE-inf as described in Methods (Statistical fine-mapping), identified top-ranked variants at loci previously implicated in cardiovascular and heart disease phenotypes, including rs11153730 (PIP = 1, p = 9.57 × 10^−23^), rs10850409 (PIP = 1, p = 2.28 × 10^−38^), and rs1324007 (PIP = 1, p = 5.53 × 10^−38^). A full summary of fine-mapping results, including PIP scores and credible sets across all loci, is available at https://finemap-app.streamlit.app.

## Discussion

In this work, we introduce a frequency-resolved framework for mapping the genetic architecture of ECG waveforms identifying 56 independent genome-wide significant loci in a sample of 47,052 individuals. By decomposing 12-lead ECGs into energy components across six wavelet detail levels and one approximation level, we uncover genetic influences that extend beyond conventional ECG intervals. Most heritable signals are concentrated in mid-frequency bands consistent with large-scale depolarization and repolarization features. However, we also observe measurable SNP-based heritability in finer-scale wavelet components and disease-associated genetic correlations in the highest-frequency band (D1, 125–250 Hz). This is notable because D1 components are not part of routine clinical ECG interpretation and are often regarded as noise or artifact.

The magnitude of heritability observed for these wavelet-derived traits supports their use as genetically informative phenotypes. Prior research, including twin studies, has established moderate to high heritability for standard ECG traits like PR, QRS, and QT intervals [22][23]. Twin studies estimates, which capture total genetic variance including contributions beyond common SNPs, typically range from 0.3-0.43 [1][19], and thus exceed SNP-based estimates. In this context, the SNP-based heritability estimates observed are substantial and comparable to other traits used successfully in GWAS. The lower h² in finer details (D2–D1) likely reflects greater environmental or artifactual influences, but it does not imply absence of genetic signal. Rather, the presence of measurable heritability and disease-associated genetic correlations in these bands suggests that even high-frequency ECG components can capture reproducible biological variation.

Beyond heritability, the wavelet-derived traits showed strong physiological face validity. The associations with standard ECG metrics recovered expected relationships between frequency leads and electrophysiological processes: QRS duration mapped most strongly to mid-frequency energy in limb leads, QT interval showed broad associations across repolarization-related bands, heart rate was associated with D3–D4 energy across leads, and R-wave axis produced opposing effects in Leads I and II consistent with cardiac vector orientation. Importantly, these relationships were markedly weaker in the D1 band. Together, these results establish two key points. First, the wavelet decomposition framework faithfully recapitulates known cardiac electrophysiology; each standard ECG metric is captured by the expected frequency band and lead combination, providing face validity for the approach. Second, and most importantly for interpreting the main findings, D1 energy appears statistically and physiologically distinct from standard ECG metrics. If D1 genetic correlations with cardiovascular disease were driven only by standard ECG features, we would expect strong overlap with established ECG metrics. Instead, the weak association between D1 and conventional ECG traits suggests that high-frequency wavelet energy captures a distinct component of ECG variation that is not routinely summarized by clinical ECG measurements. The significant genetic correlations between D1 energy and cardiovascular disease reported in the main results therefore cannot be explained by D1 proxying for established ECG phenotype, and instead reflect genuinely novel biological variation in high-frequency cardiac signals that is invisible to routine clinical interpretation.

Our findings raise several possible interpretations. One possibility is that high-frequency activity contains physiologically meaningful information that is lost during standard ECG filtering and visual inspection. Prior methodological work has shown that high-frequency components of the QRS complex can reflect subtle alterations in myocardial activation that are not visible in the time-domain waveform [18]. For example, studies of signal-averaged ECG have demonstrated that high-frequency “late potentials” (typically 40–250 Hz) can identify micro-reentrant conduction zones and risk of arrhythmia [16]. More recent work using ultra-high-frequency ECG has shown that frequencies well above the standard diagnostic band can track ventricular activation timing [17]. These studies collectively suggest that high-frequency ECG components, while visually imperceptible and traditionally omitted, may carry biological signals under certain conditions. Ultimately, the presence of significant genetic correlations with heart failure (rg = 0.56 in V1, p = 0.0039) and coronary atherosclerosis (rg = 0.39 in V1, p = 0.0027) in D1 signal suggests that at least part of the high-frequency content reflects consistent, biologically relevant variation rather than acquisition artifact. Notably, the strongest D1 signal for coronary atherosclerosis was observed in V1, a right precordial lead overlying the right ventricle and interventricular septum, rather than in the anterior and lateral leads (V3–V6) classically associated with atherosclerotic ECG changes. Combined with the weak association between D1 energy and standard ECG metrics, these findings support the interpretation that D1 captures a distinct high-frequency component of cardiac electrophysiology that is not represented by routine clinical ECG phenotypes, and that this component carries genetic signal of direct relevance to cardiovascular disease.

The BMI genetic correlation analysis addressed a key alternative explanation: that D1 signal reflects adiposity-related EMG artifact rather than cardiac biology. The observed BMI pattern followed the expected geometry of obesity-related leftward axis deviation: left-oriented limb leads, including Lead I and aVL, showed positive genetic correlations with BMI, whereas inferior leads showed negative or near-zero correlations. This triaxial signature is consistent with diaphragmatic elevation rotating the mean QRS vector leftward and superiorly, augmenting projection onto Lead I and aVL while diminishing it over the inferior leads. Recovery of this axis geometry by a genome-wide method, without any anatomical supervision, validates that wavelet energy features are sensitive to physiologically meaningful cardiac geometric variation. Three lines of evidence argue against contamination by skeletal muscle EMG artifact. First, the genetic correlation between D1 energy and BMI does not follow the pattern predicted by an EMG artifact model. Subcutaneous fat attenuates surface electrode EMG amplitude by increasing the distance and impedance between muscle and electrode, so if D1 energy were substantially driven by fat-modulated EMG, we would predict a negative rg between BMI and D1 energy; instead, the observed rg is positive for left-oriented leads and near zero for inferior leads, matching precisely the spatial signature of obesity-related leftward cardiac axis deviation. This is mechanistically inconsistent with EMG artifact and fully consistent with the known cardiac geometric consequences of increased adiposity, in which diaphragmatic elevation shifts the mean QRS vector superiorly and leftward. Second, the BMI–D1 rg collapses precisely at D1 for the inferior leads most sensitive to axis deviation, whereas a fat-artifact model would predict its strongest expression at the highest frequencies, where surface noise dominates, the opposite of what is observed. Third, the top GWAS loci in D1 map to cardiac conduction genes, most prominently SCN5A, encoding the primary cardiac sodium channel Nav1.5, rather than skeletal muscle structural genes, and their effect sizes are negligibly attenuated (< 2%) after conditioning on BMI. The partial genetic correlation between D1 energy and heart failure, after mathematically removing the contribution of shared BMI genetics, remains substantial across leads, confirming that the D1–cardiovascular disease genetic signal reflects a direct cardiac biological pathway rather than an adiposity-mediated artifact.

A robustness check excluding participants with prevalent heart failure, cardiomyopathy, and myocardial infarction ruled out reverse causation as the sole driver of this correlation (rg = 0.38, p = 0.003 in disease-free individuals), though the residual signal likely reflects a mixture of subclinical disease liability, a shared mediating risk factor, or both — completely independent pleiotropy is unlikely, given that both phenotypes are cardiac in nature. Several interpretations of the residual signal and its attenuation are possible. First, the attenuation may partly reflect a continuous disease liability gradient — individuals with high genetic risk for heart failure may already show subtle pre-diagnostic alterations in high-frequency ventricular conduction, consistent with high-frequency late potentials detected prior to clinical events in signal-averaged ECG studies [16][18]. Second, the correlation may be partly mediated by a shared causal risk factor that independently drives both D1 energy and heart failure risk. Completely independent pleiotropy is unlikely given that both phenotypes are cardiac in nature. Definitively separating these mechanisms would require Mendelian randomization or prospective longitudinal ECG data, which we leave for future work.

Setting aside the interpretation of D1, our findings demonstrate that wavelet-based ECG features capture a broad spectrum of heritable electrophysiological patterns. Many loci identified across leads and frequency levels map to genes implicated in cardiomyopathy, conduction disorders, and cardiac structural integrity, including ALPK3, DSP, FHOD3, SCN10A, SCN5A, KCNQ1, TTN, and others [3][25]. The appearance of less well-characterized genes (e.g., ANKRD1, ZFPM2) in enriched cardiac pathways further indicates that this approach may help refine candidate gene lists for electrophysiological phenotypes. Well-established genes like ALPK3 (linked to hypertrophic cardiomyopathy), DSP (associated with arrhythmogenic cardiomyopathy and myocarditis-like episodes), KCNQ1 (implicated in long QT syndrome and arrhythmogenic phenotypes), SCN5A (causing arrhythmogenic dilated cardiomyopathy and conduction disorders), and TTN (a major cause of dilated cardiomyopathy via truncating variants) show strong genotype-phenotype links to cardiac diseases. In contrast, less well-known genes such as ANKRD1, FHOD3, and ZFPM2 (appearing in cardiomyopathy gene panels with potential roles) are also enriched in these terms, suggesting novel contributions to cardiac pathology despite limited prior characterization. These results reinforce the connection between the associated genes and cardiovascular traits, complementing the genetic correlations observed with FinnGen phenotypes.

Large-scale ECG GWAS have to date relied overwhelmingly on visually interpreted, time-domain interval phenotypes (QRS duration, QT interval, PR interval, and heart rate) whose locus architectures are now relatively well characterized [3][26]. The present study demonstrates that this phenotyping strategy captures only a portion of the heritable variation embedded in the waveform: moving from discrete intervals to frequency-resolved energy features uncovers genetic signal, including disease-relevant associations in the highest-frequency band, that is structurally inaccessible to interval-based approaches [28]. Several questions and limitations define the immediate agenda for extending this framework. All analyses were restricted to White British UK Biobank participants, meaning the identified loci and fine-mapped variants reflect European allele frequencies and LD structure, and the wavelet energy distributions and derived heritability estimates may not transfer to other populations, as ECG morphology is known to vary systematically across ancestries; replication in ancestrally diverse cohorts therefore remains a prerequisite before clinical deployment can be considered. Whether high-frequency genetic signal carries causal clinical utility independent of established ECG traits requires formal causal inference, such as Mendelian randomization against incident cardiovascular endpoints. Whether the approach generalizes to higher-frequency or signal-averaged recordings — in which additional heritable components of myocardial activation may become resolvable, including high-frequency late potentials and rare events such as extrasystoles — is a technical question that motivates future data collection. And whether the decomposition framework could extend to other quasi-periodic physiological signals, such as EEG, to enable analogous multi-scale genetic mapping across organ systems remains an open question that would require independent validation in those domains.

## Conclusion

ECG genetics has largely been shaped by an assumption of clinical legibility that the traits most worth mapping are those visible to the cardiologist. This study challenges that assumption. By decomposing resting 12-lead ECGs from 47,052 UK Biobank individuals into frequency-specific energy components through Daubechies wavelet analysis, we show that the cardiac waveform contains heritable variation distributed across its full spectral range, much of which is invisible under standard clinical filtering. Across 84 wavelet-derived traits, we identified independent loci that converge on genes and pathways central to cardiomyocyte biology, cardiac conduction, and structural integrity, including SCN5A, KCNQ1, TTN, and DSP, as well as less-characterized candidates such as ANKRD1 and ZFPM2 whose electrophysiological roles merit further investigation. Most notably, energy in the highest-frequency detail band (D1, 125–250 Hz) showed significant disease-associated genetic correlation despite being largely absent from routine ECG interpretation and often attributed as noise. Its genetic correlation with heart failure, together with weak associations with standard ECG metrics and robustness to BMI-related sensitivity analyses, suggests that at least part of this high-frequency signal reflects reproducible cardiac biology rather than nonspecific artifact. More broadly, our findings suggest that the genetic architecture of the ECG waveform extends beyond clinically named segments and includes frequency-specific components that may capture previously under-characterized aspects of cardiac electrophysiology. The analytical framework and code are publicly available at https://github.com/rivas-lab/waveform_disease.git. Looking ahead, this approach could inform precision medicine by guiding targeted genetic testing based on lead- and frequency-specific signatures.

## AI Assistance Statement

During the preparation of this manuscript, the authors used Claude (Anthropic), ChatGPT (OpenAI), and Grok (xAI) to assist with code generation and debugging, brainstorming and structural organization, literature exploration, and editorial refinement. All AI-assisted content was reviewed, validated, and edited by the authors to ensure scientific accuracy and methodological integrity. The authors assume full responsibility for the final content of this manuscript. No AI tool was used to generate original research data or to interpret primary results independently of human oversight.

## Author Contributions

M.A.R. designed the study and provided overall supervision. S.Z. led the analysis and manuscript writing. J.W.O’S. and E.A.A. provided clinical and cardiological supervision. T.H., R.T. contributed to the statistical and methodological framework. L.L. contributed to manuscript writing. T.K. provided feedback on the manuscript and conducted statistical analysis. All authors reviewed the manuscript.

## Data Availability

The data used in this study are available from the UK Biobank resource upon application. Due to data access restrictions, the authors cannot share individual-level data.

## Acknowledgements

This research has been conducted using the UK Biobank Resource under Application Number 24983, “Generating effective therapeutic hypotheses” (PI: M.A.R.). Based on the information provided in Protocol 44532, the Stanford IRB has determined that the research does not involve human subjects as defined in 45 CFR 46.102(f) or 21 CFR 50.3(g). All participants in the UK Biobank provided written informed consent (more information is available at https://www.ukbiobank.ac.uk/2018/02/gdpr/). We thank all the participants in the UK Biobank study. Some of the computing for this project was performed on the Sherlock cluster. We would like to thank Stanford University and the Stanford Research Computing Center for providing computational resources and support that contributed to these research results. We would like to thank all participants of the FinnGen study for their generous participation.

## Supplementary Material

**Supplementary Figures 1.**
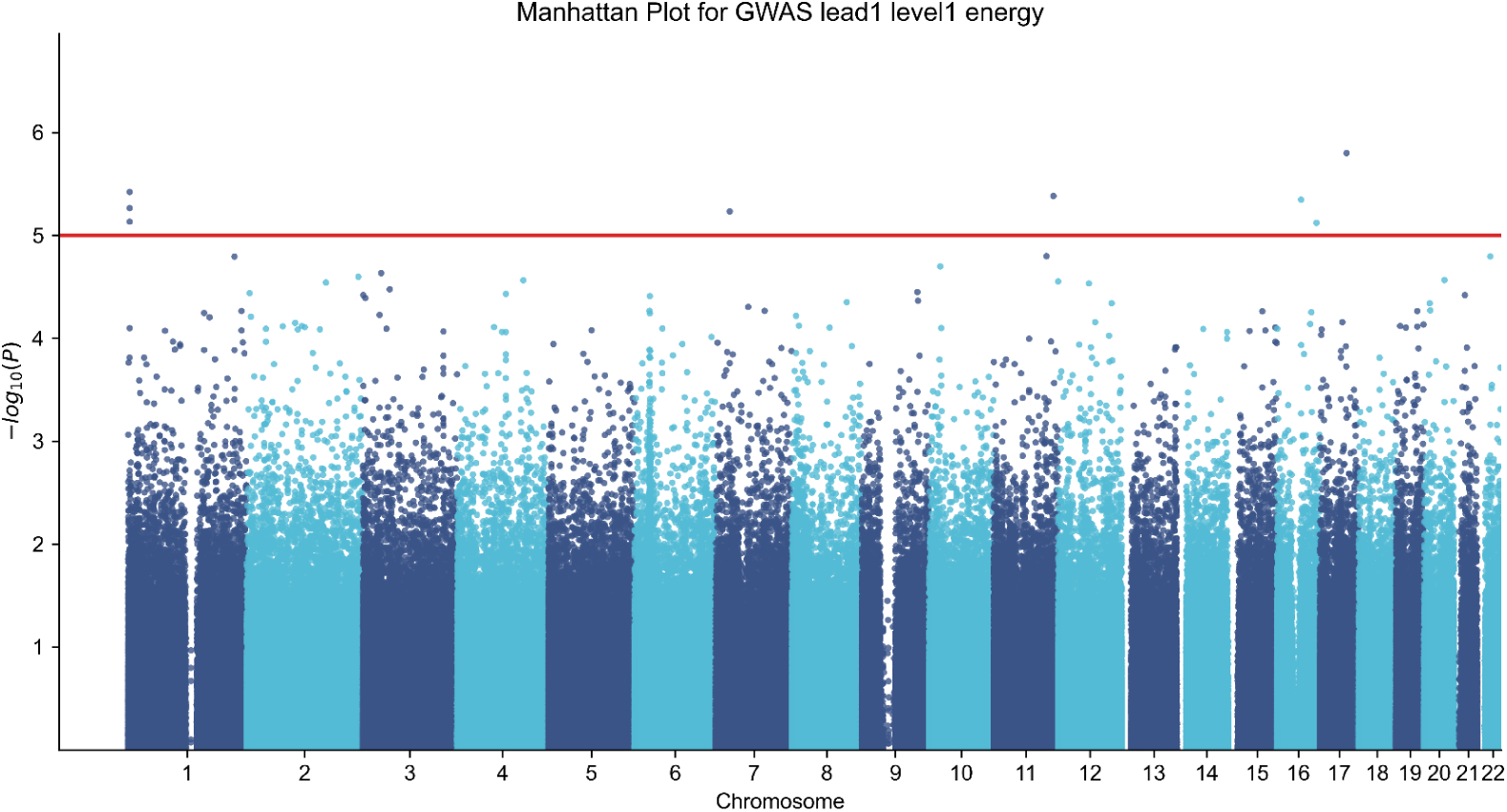
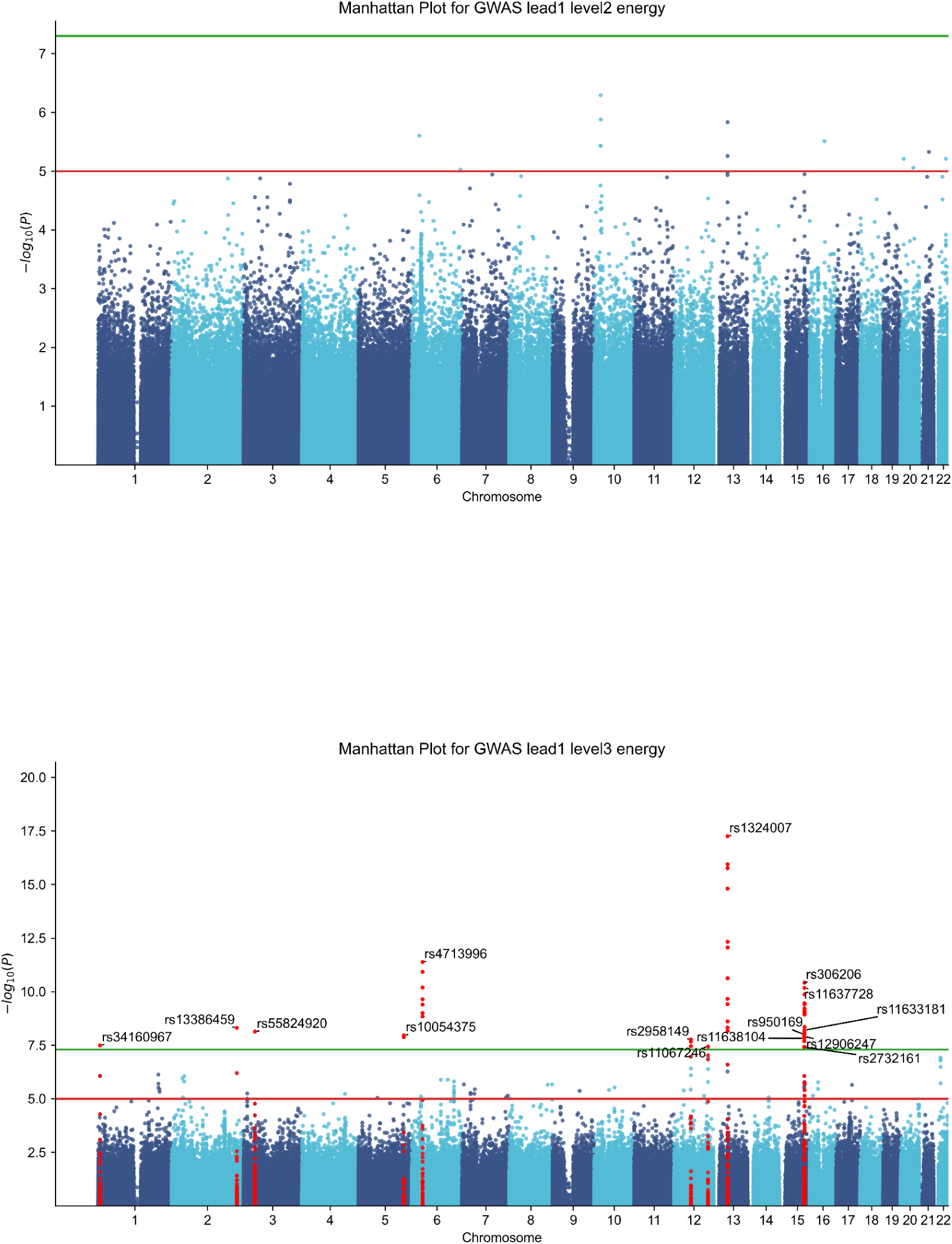
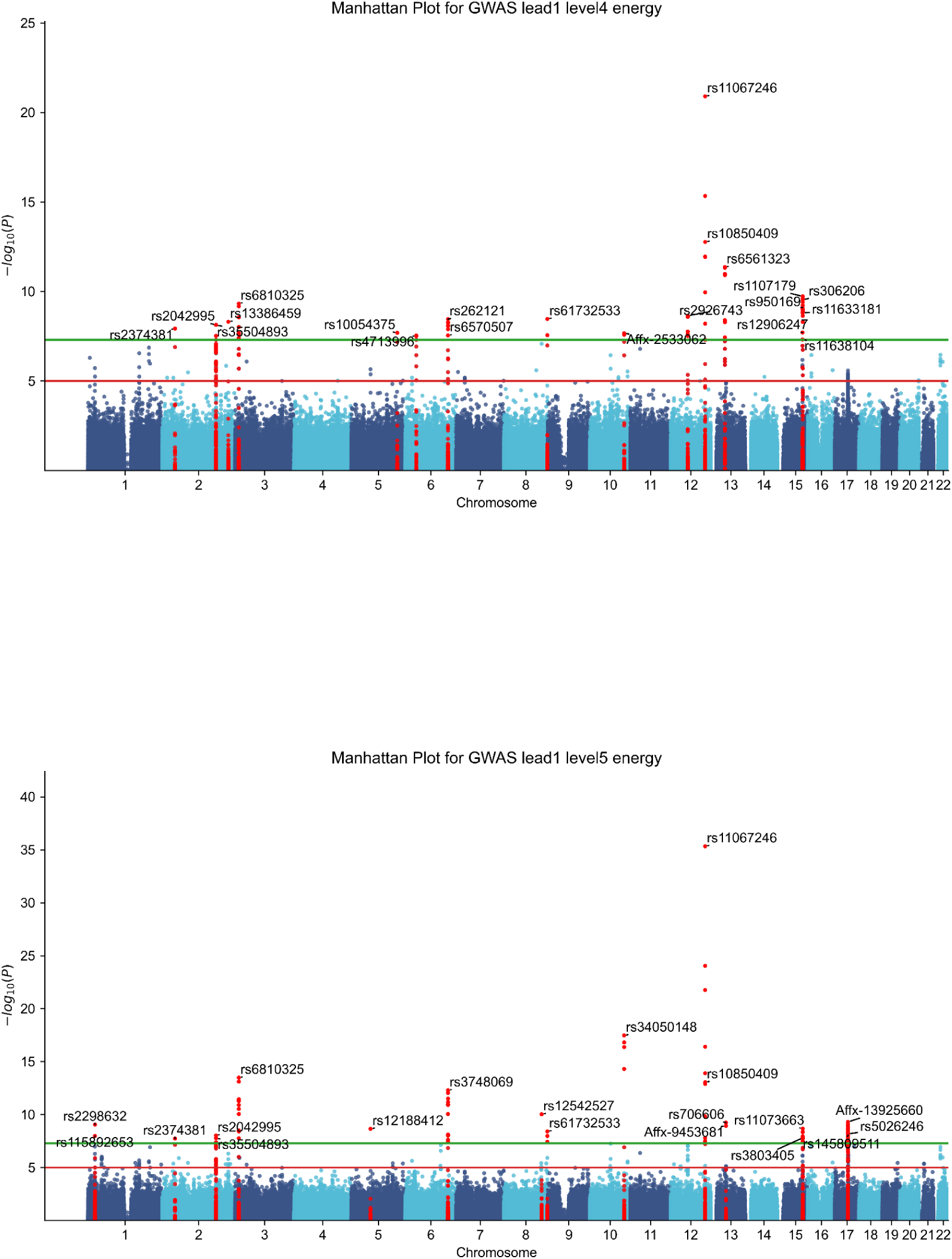
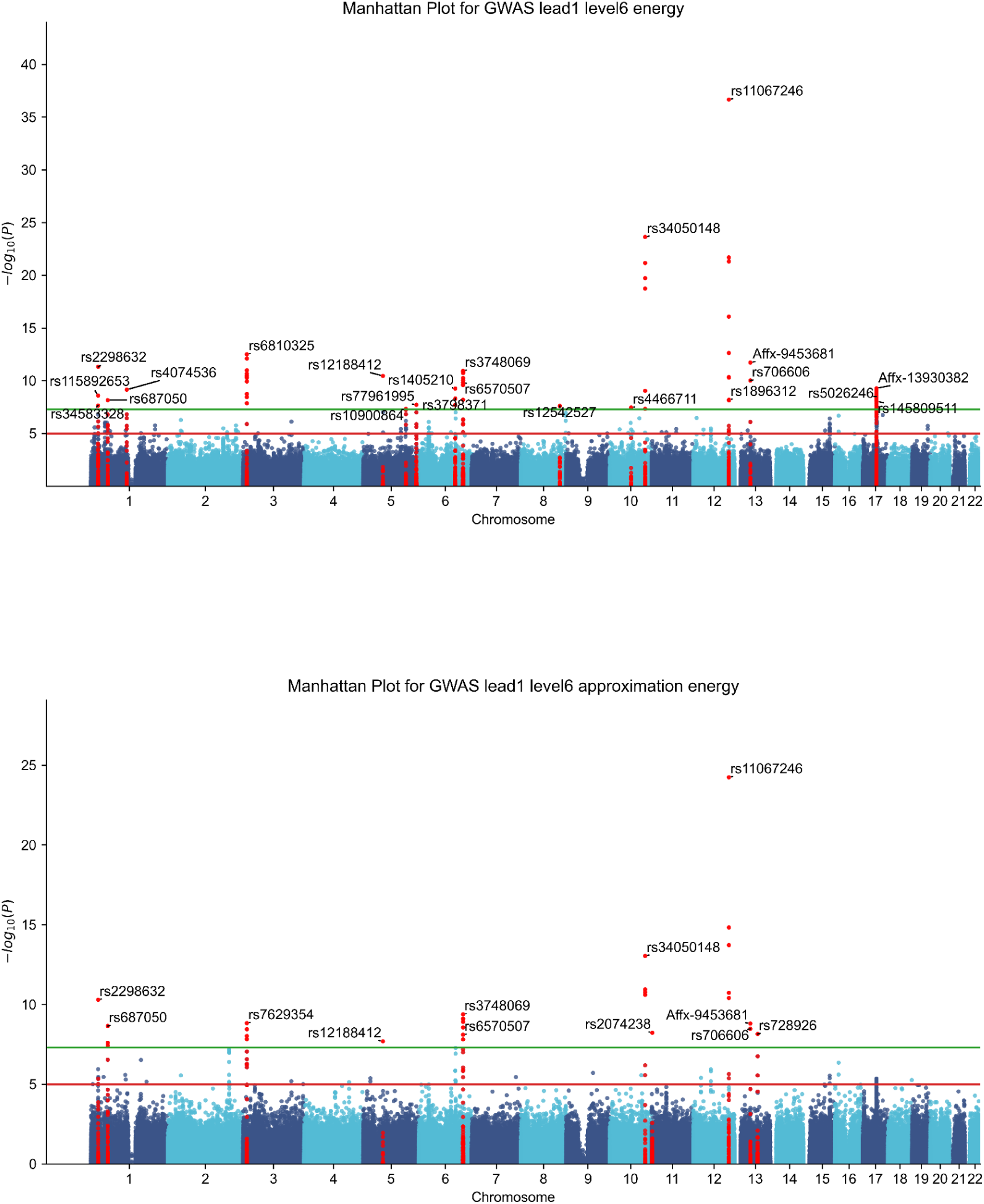
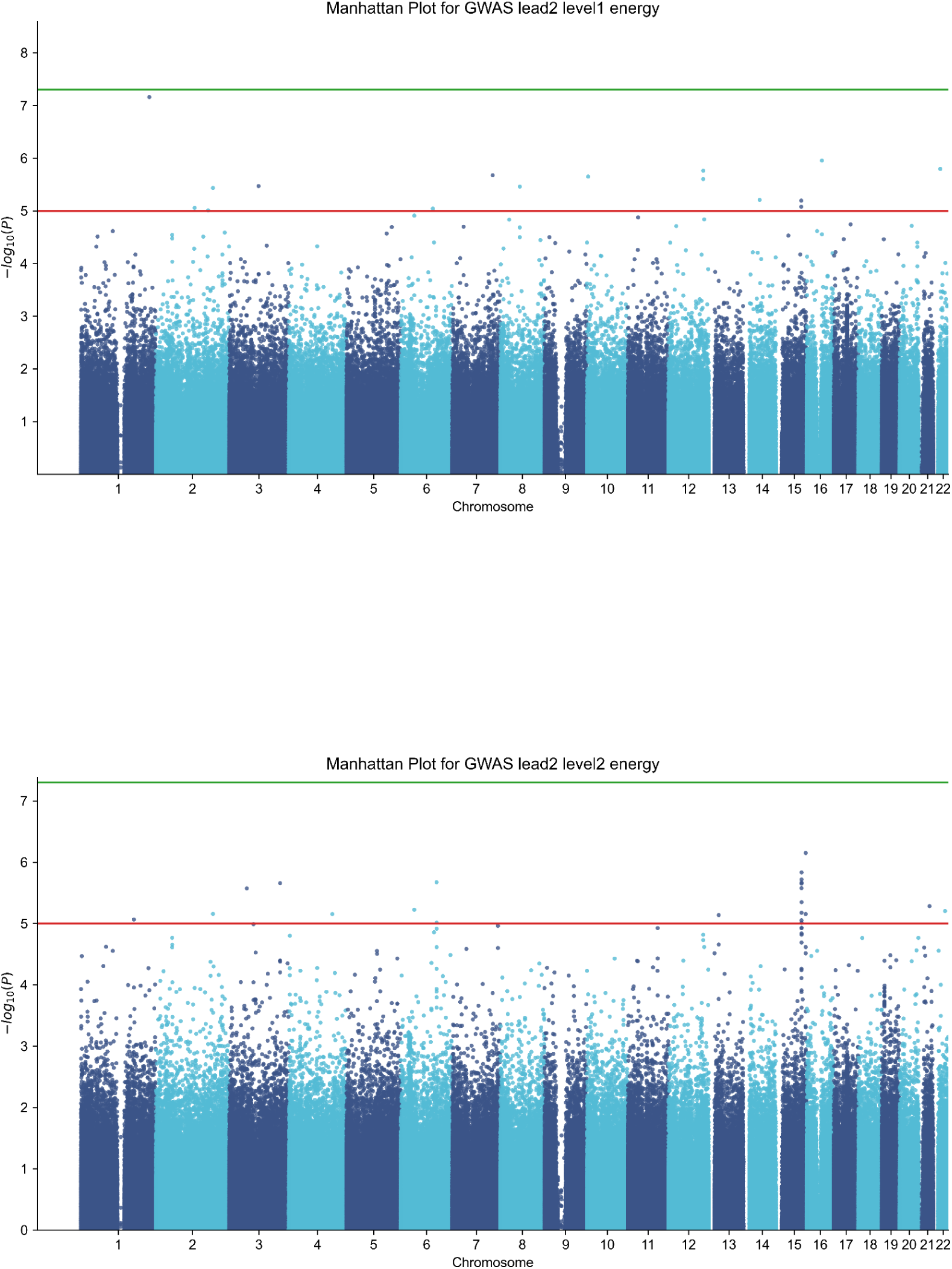
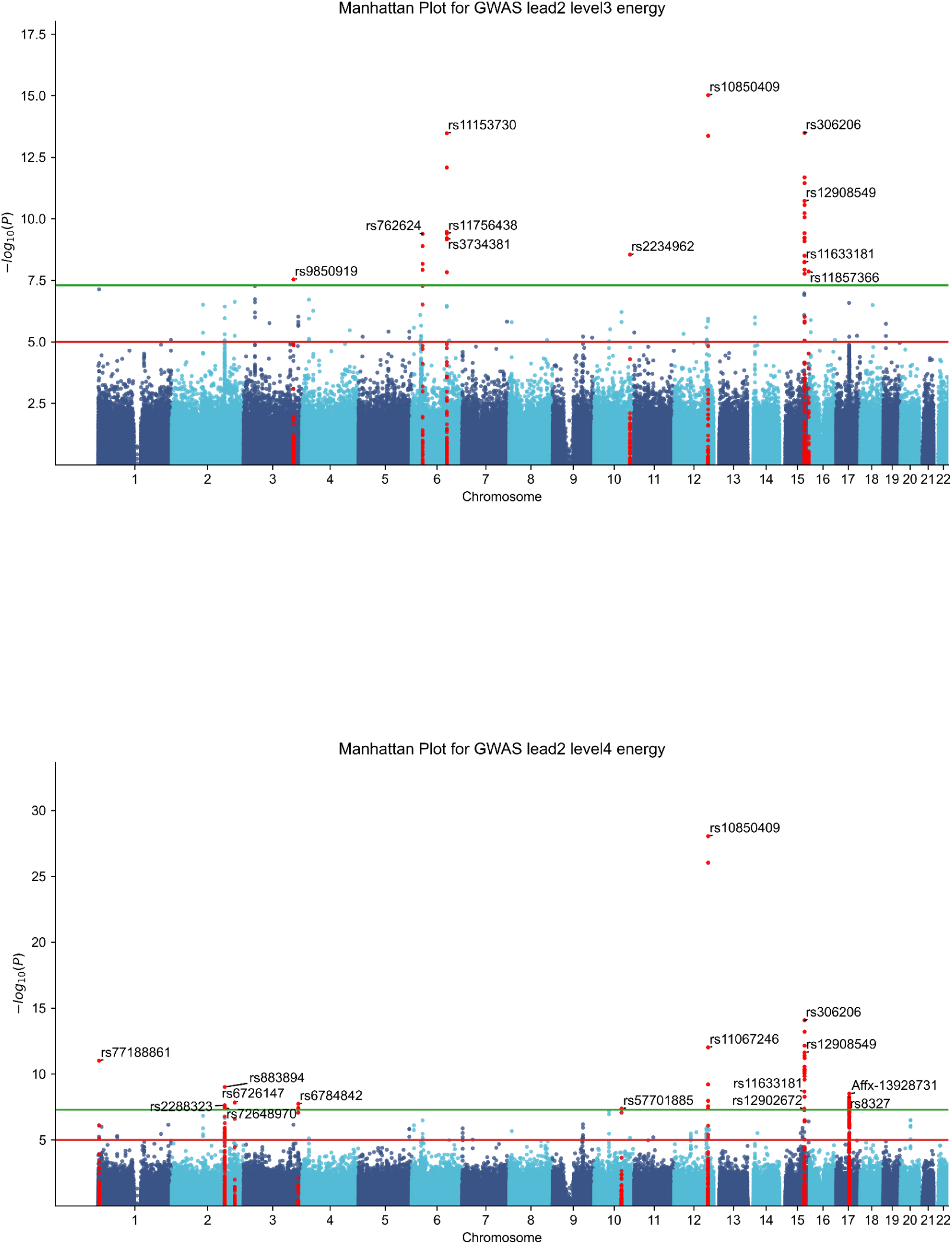
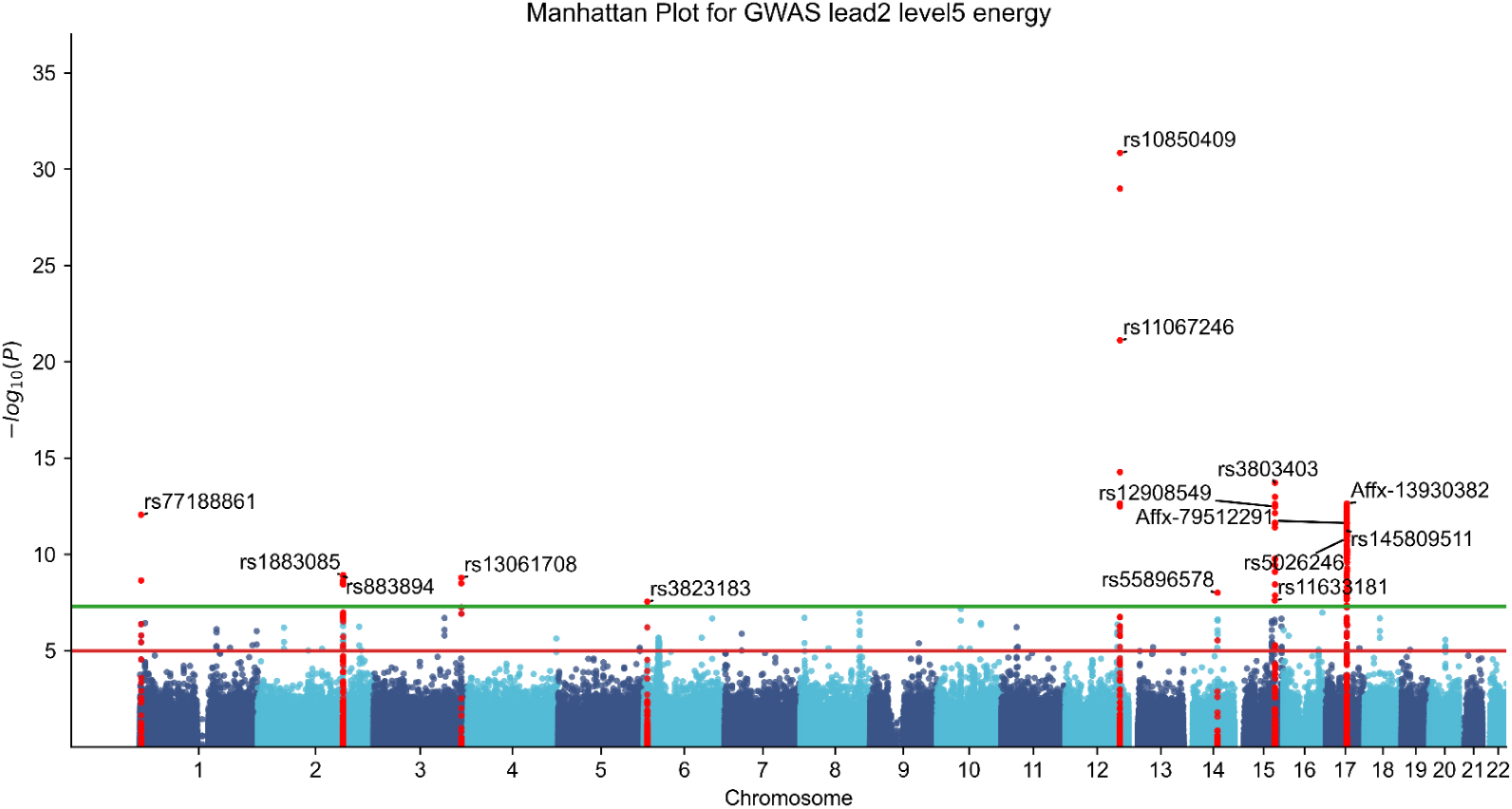
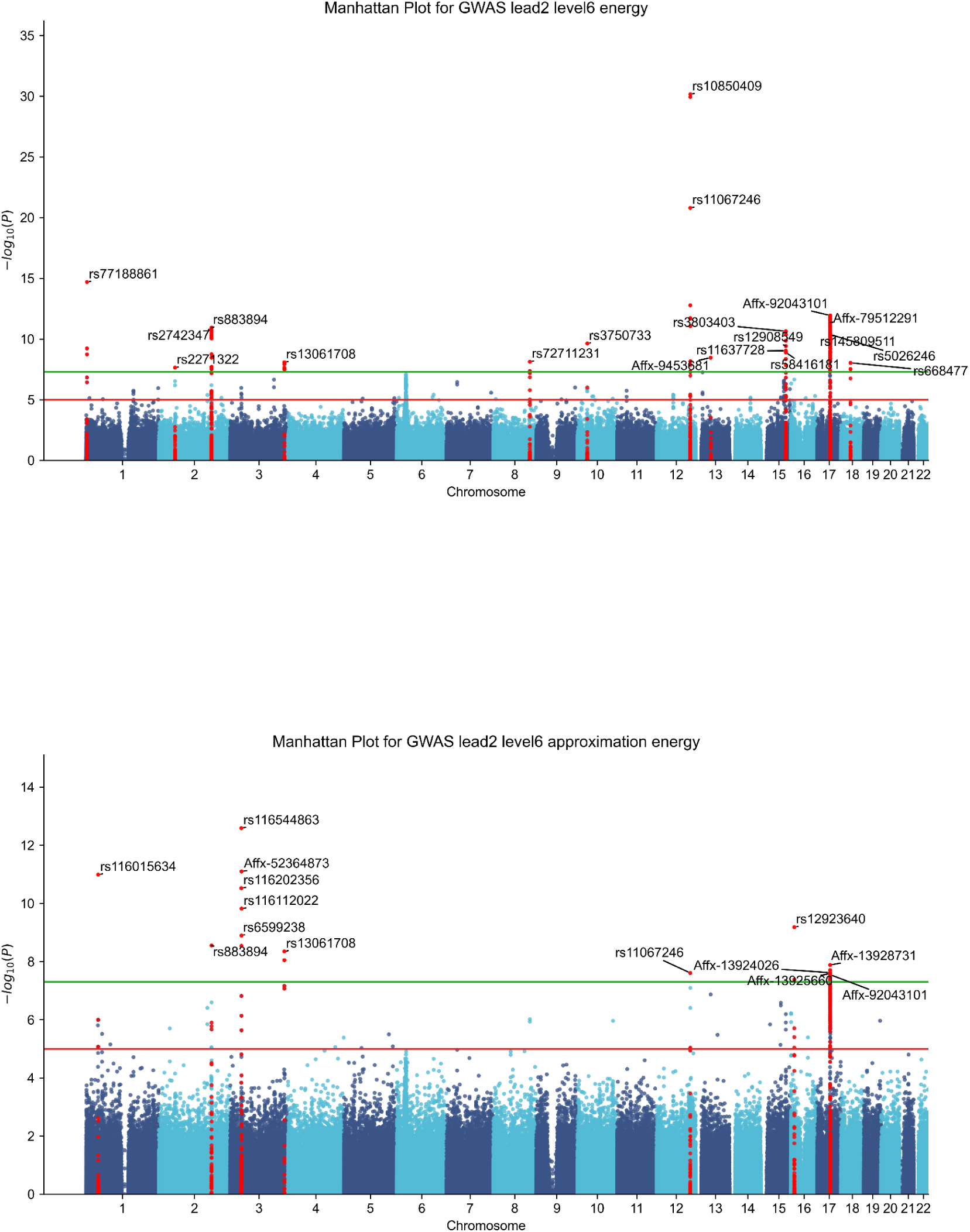
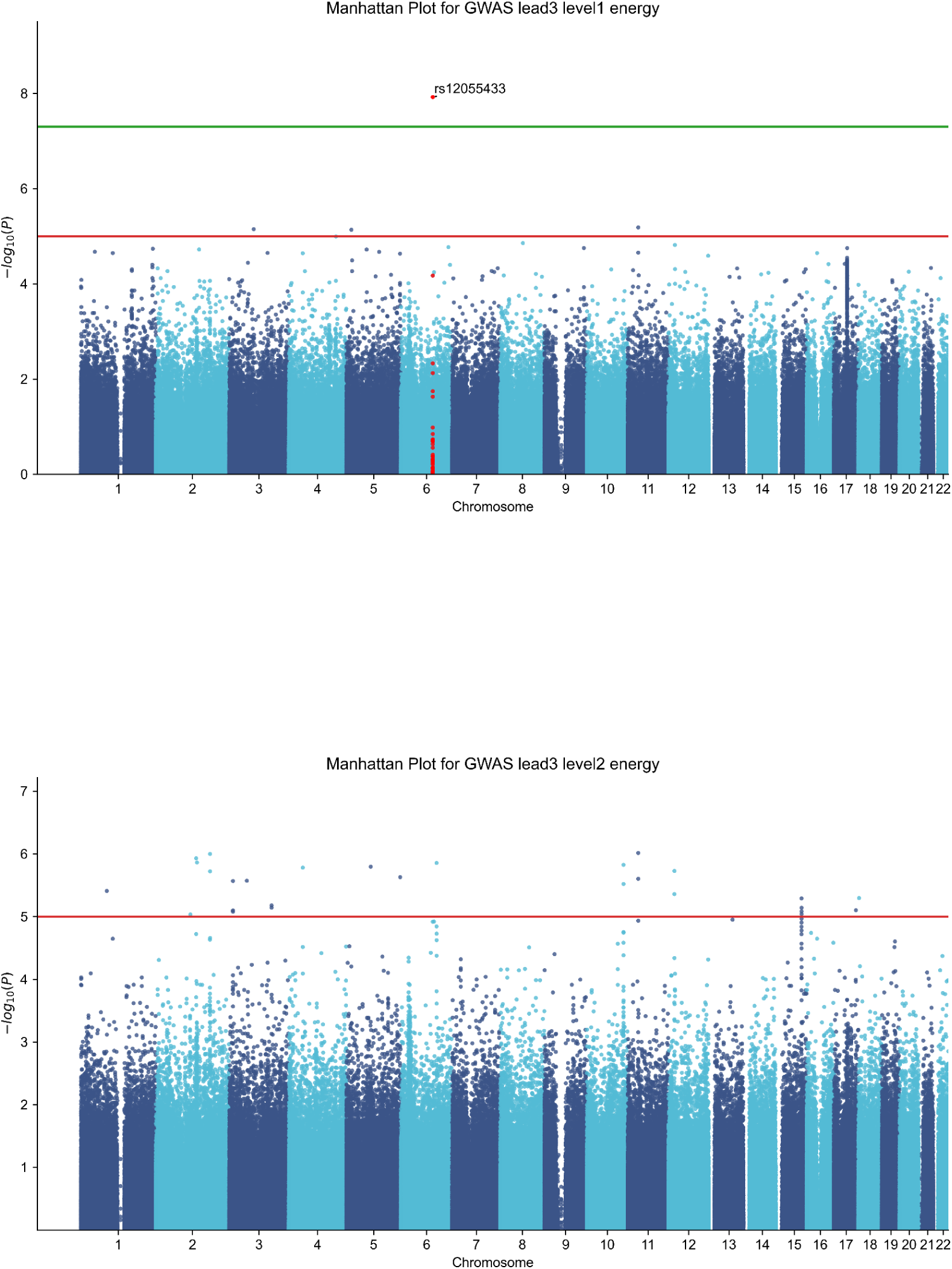
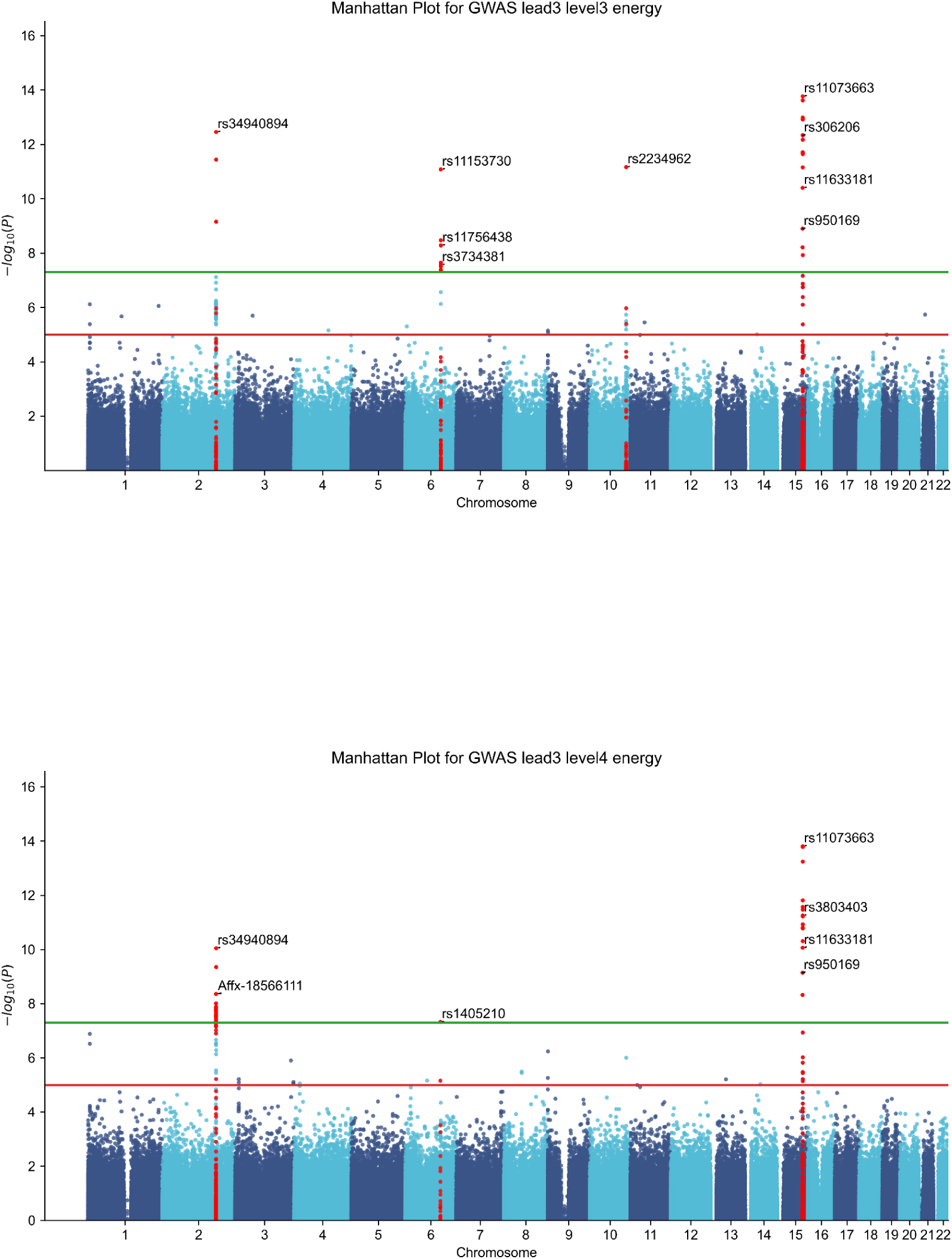
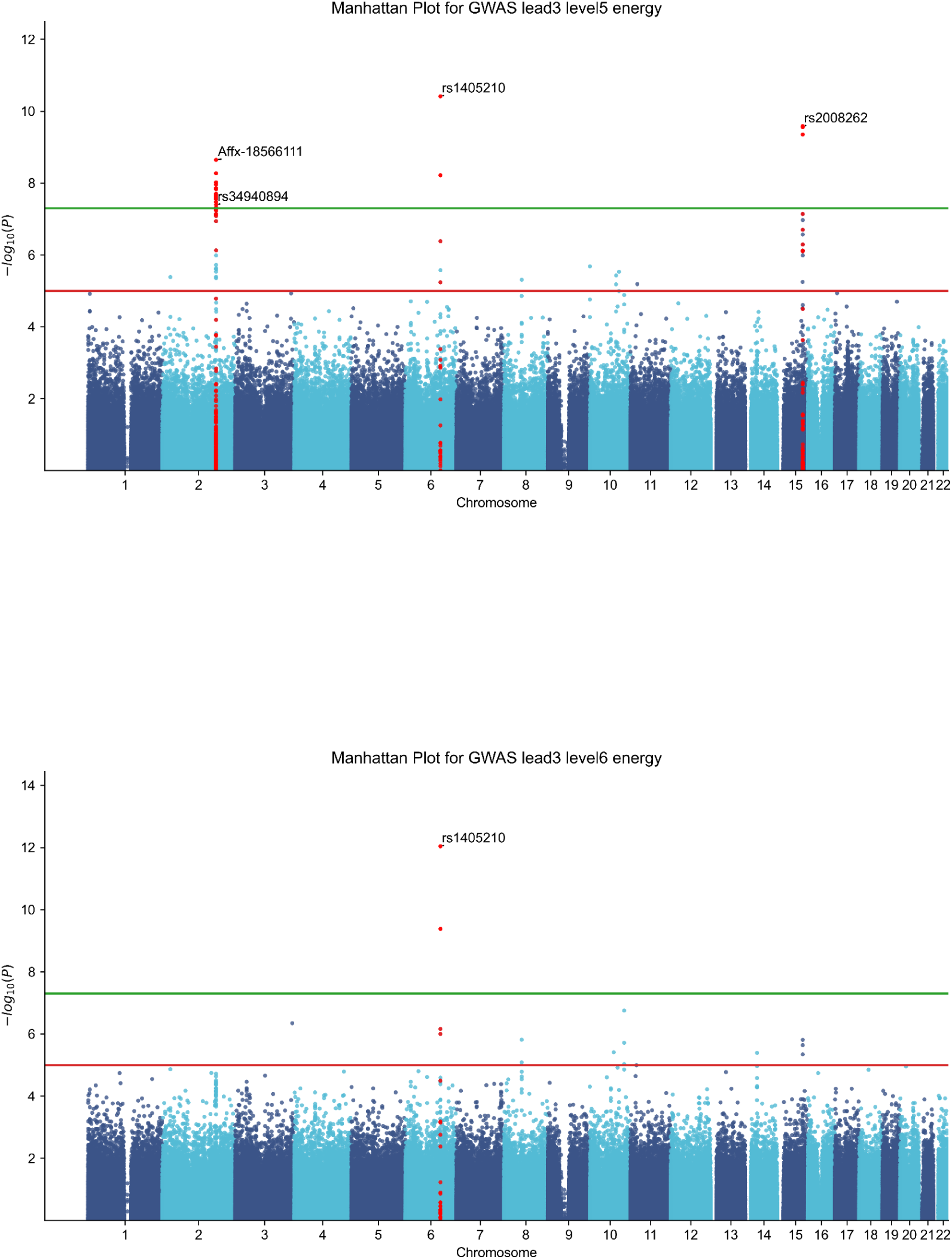
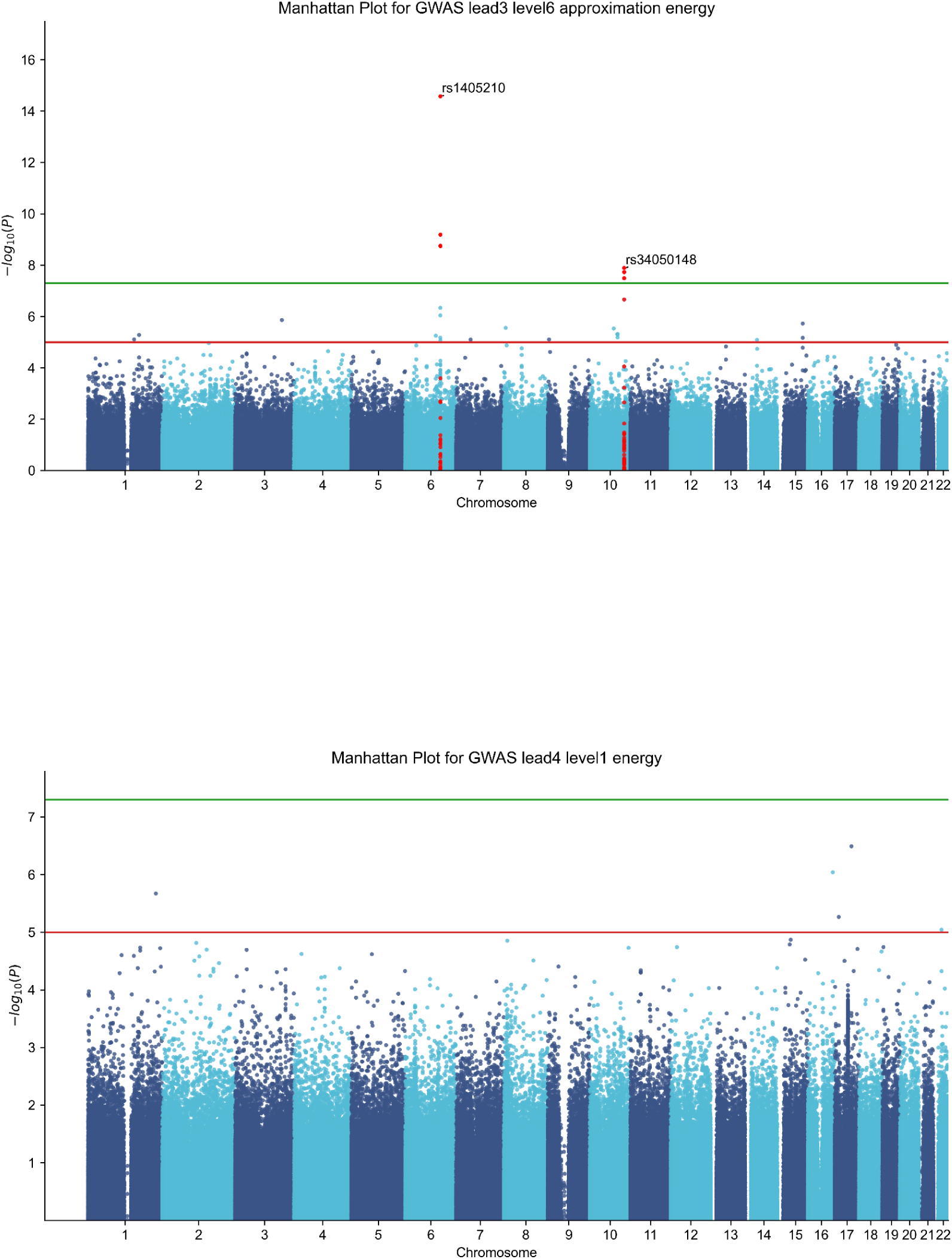
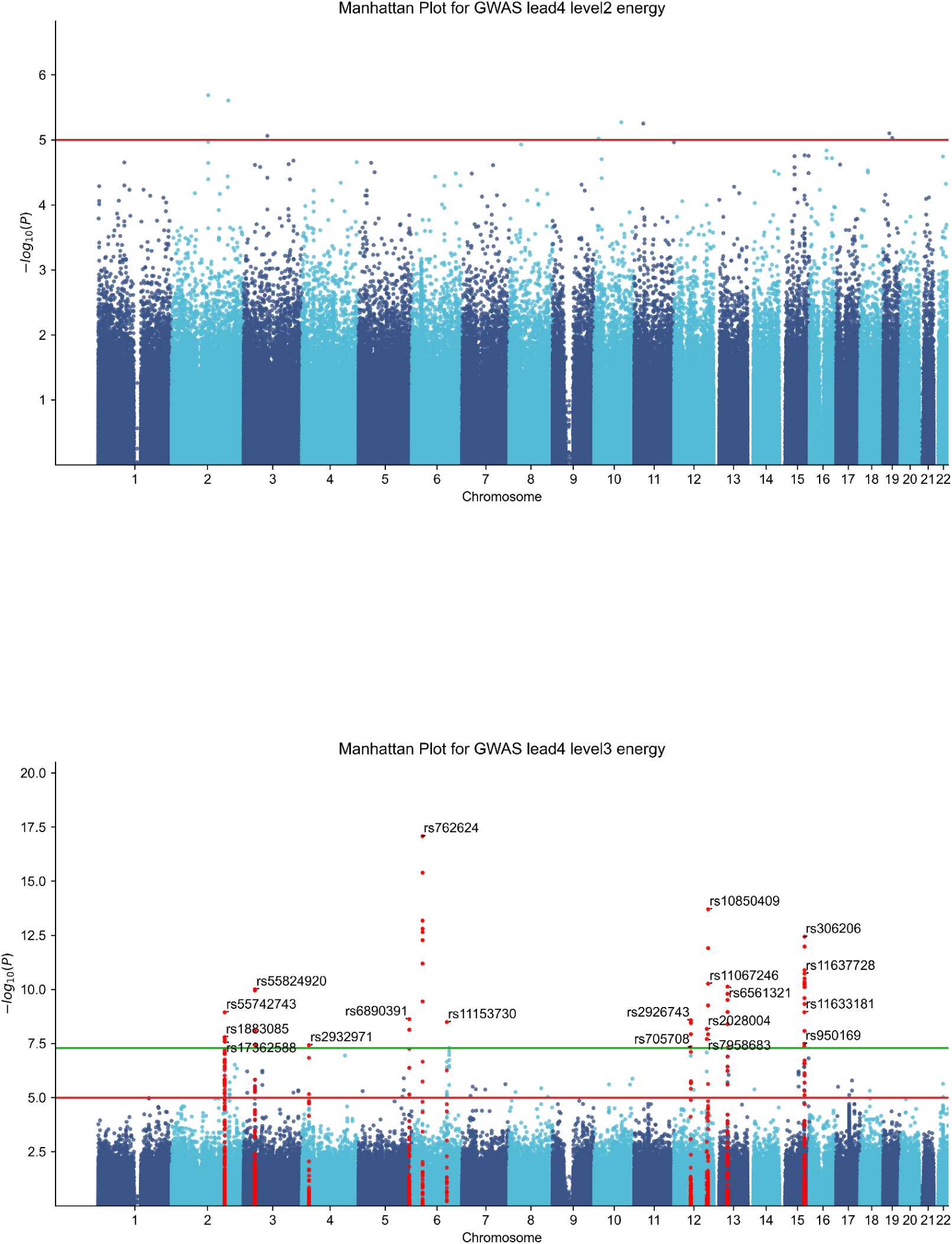
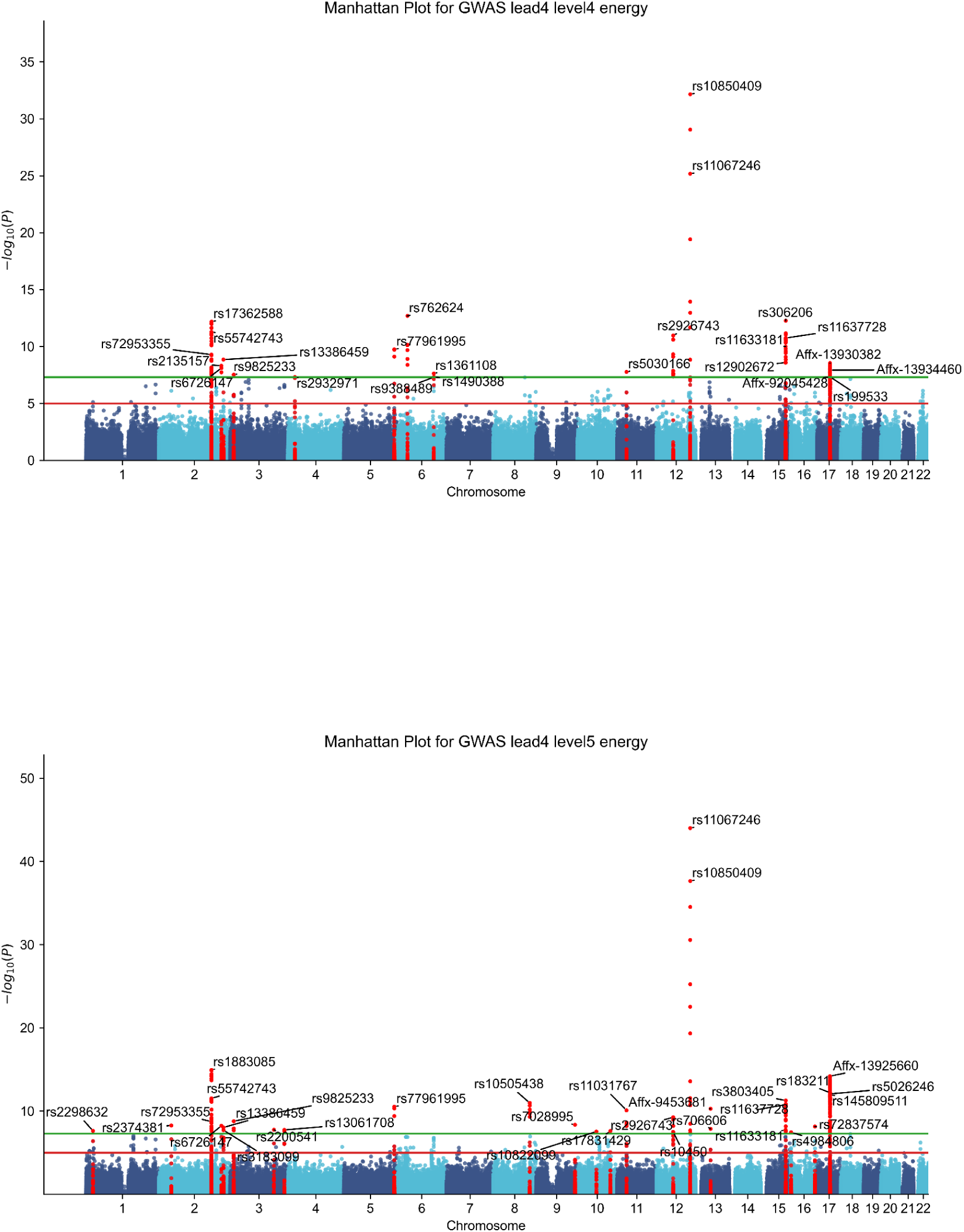
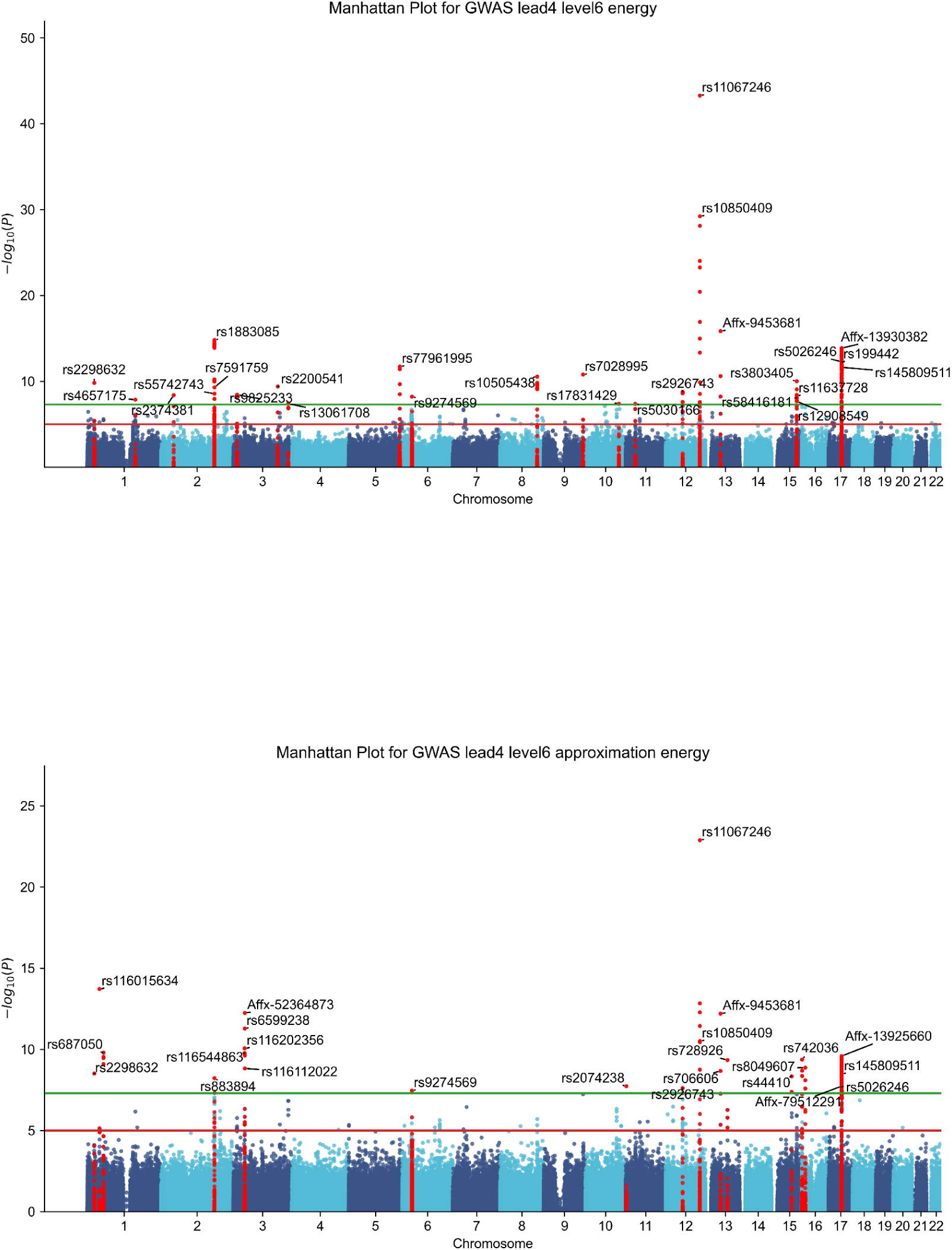
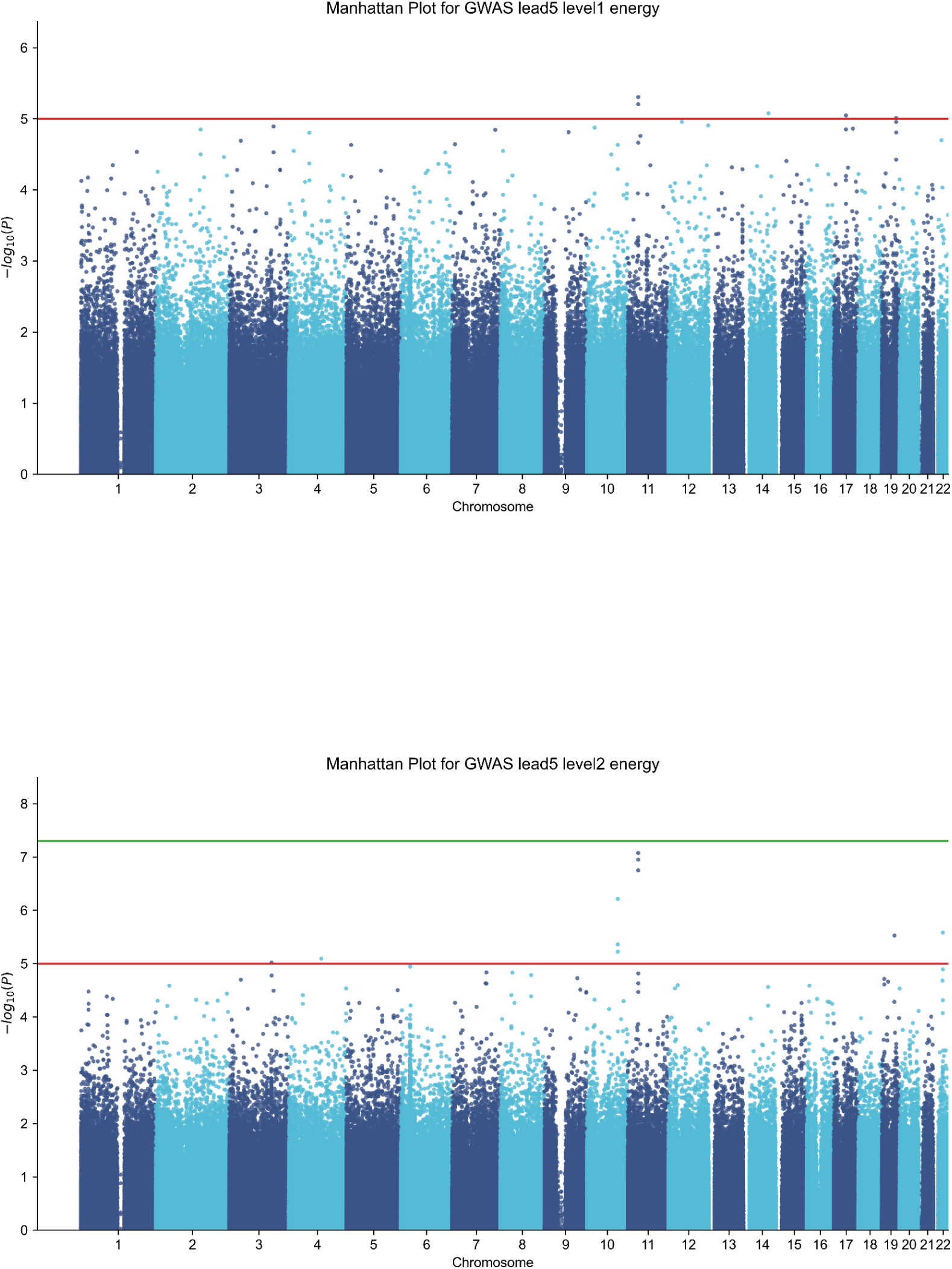
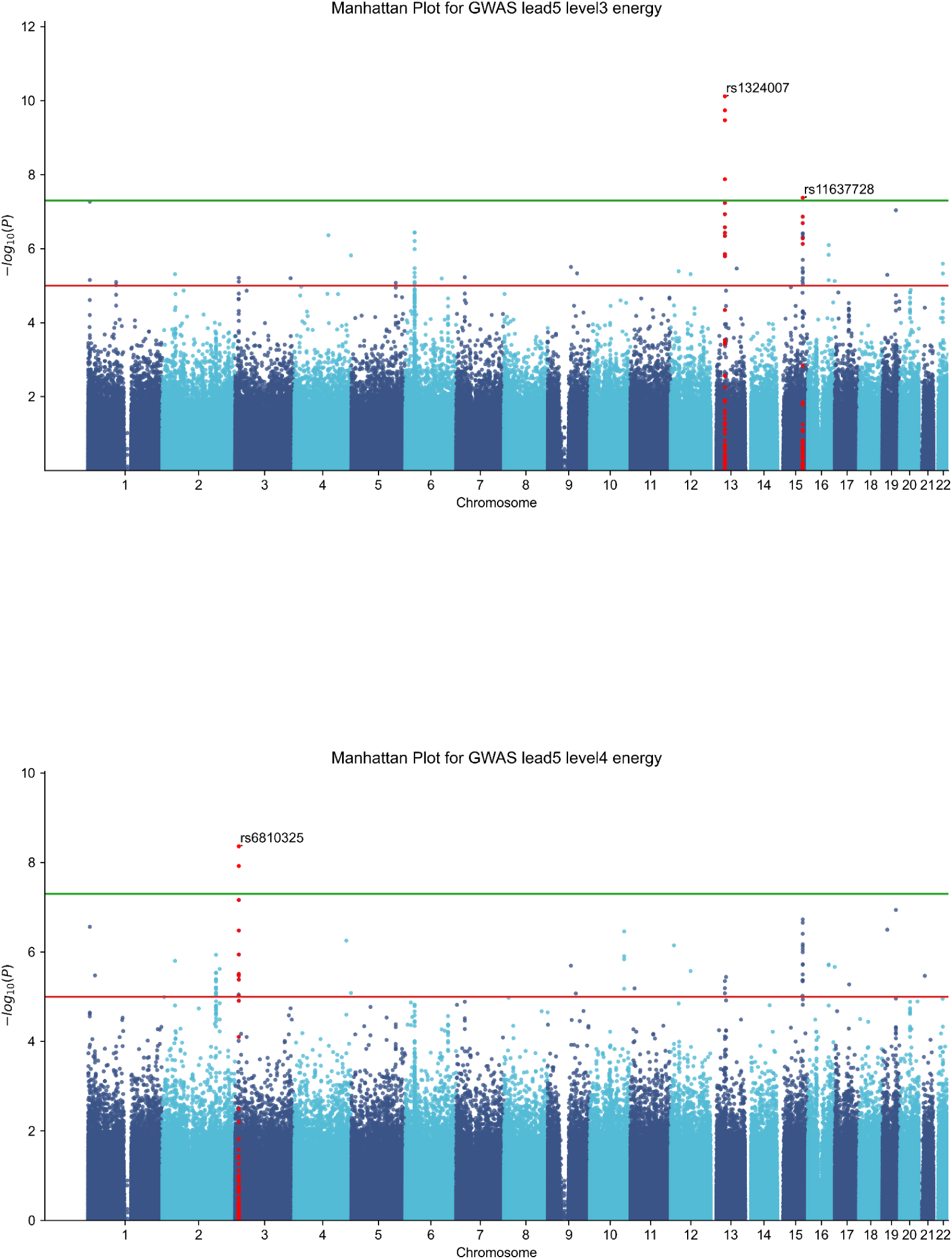
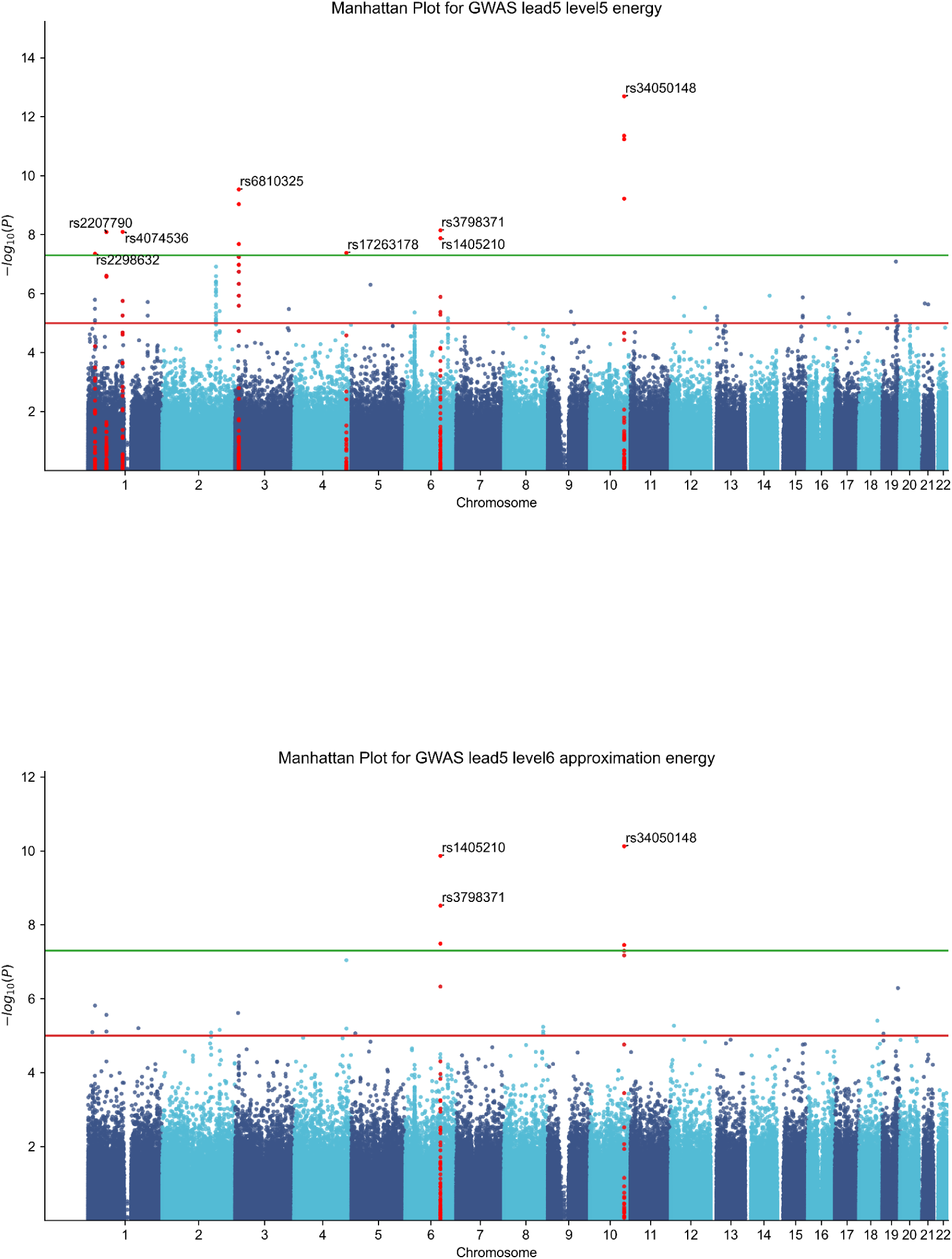
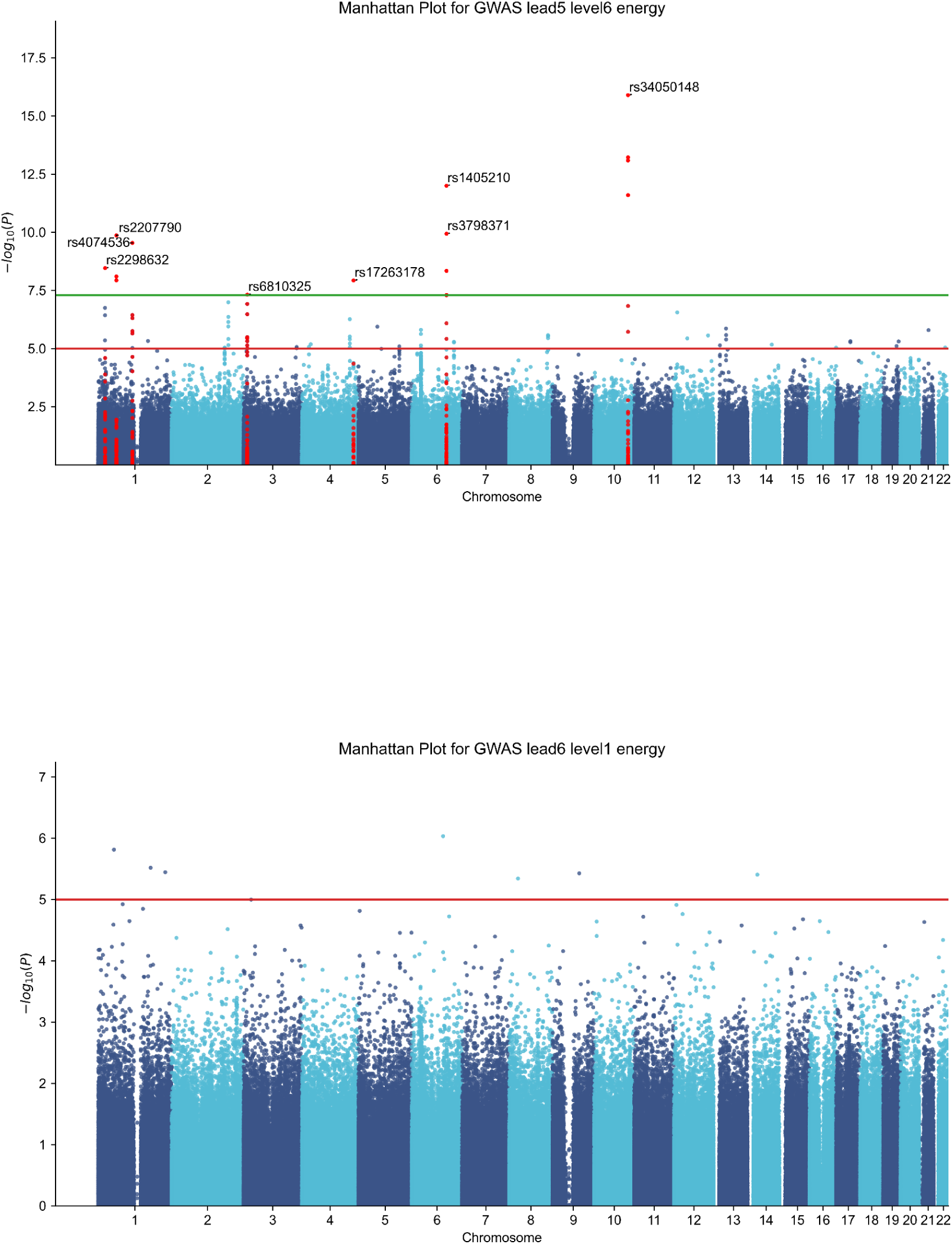
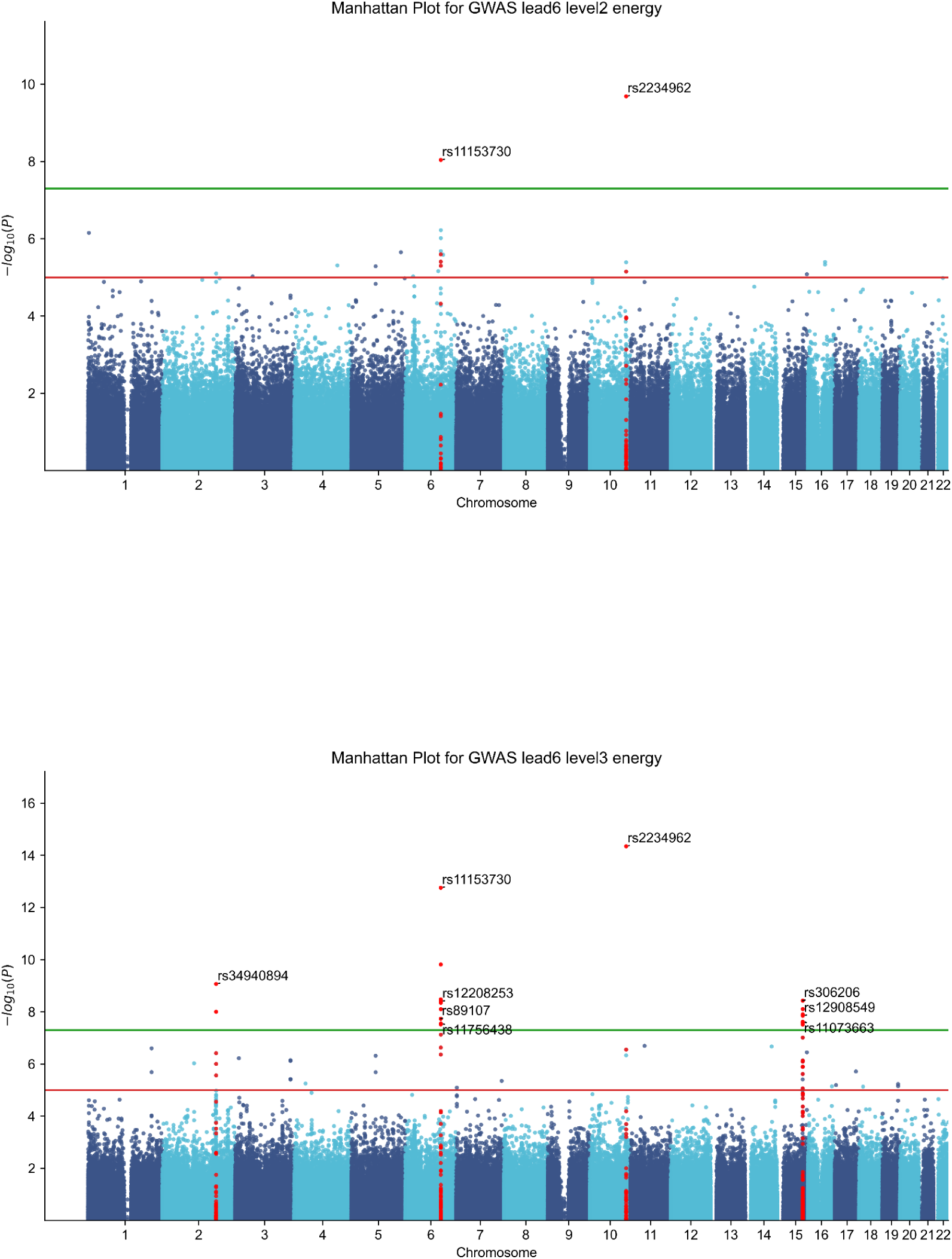
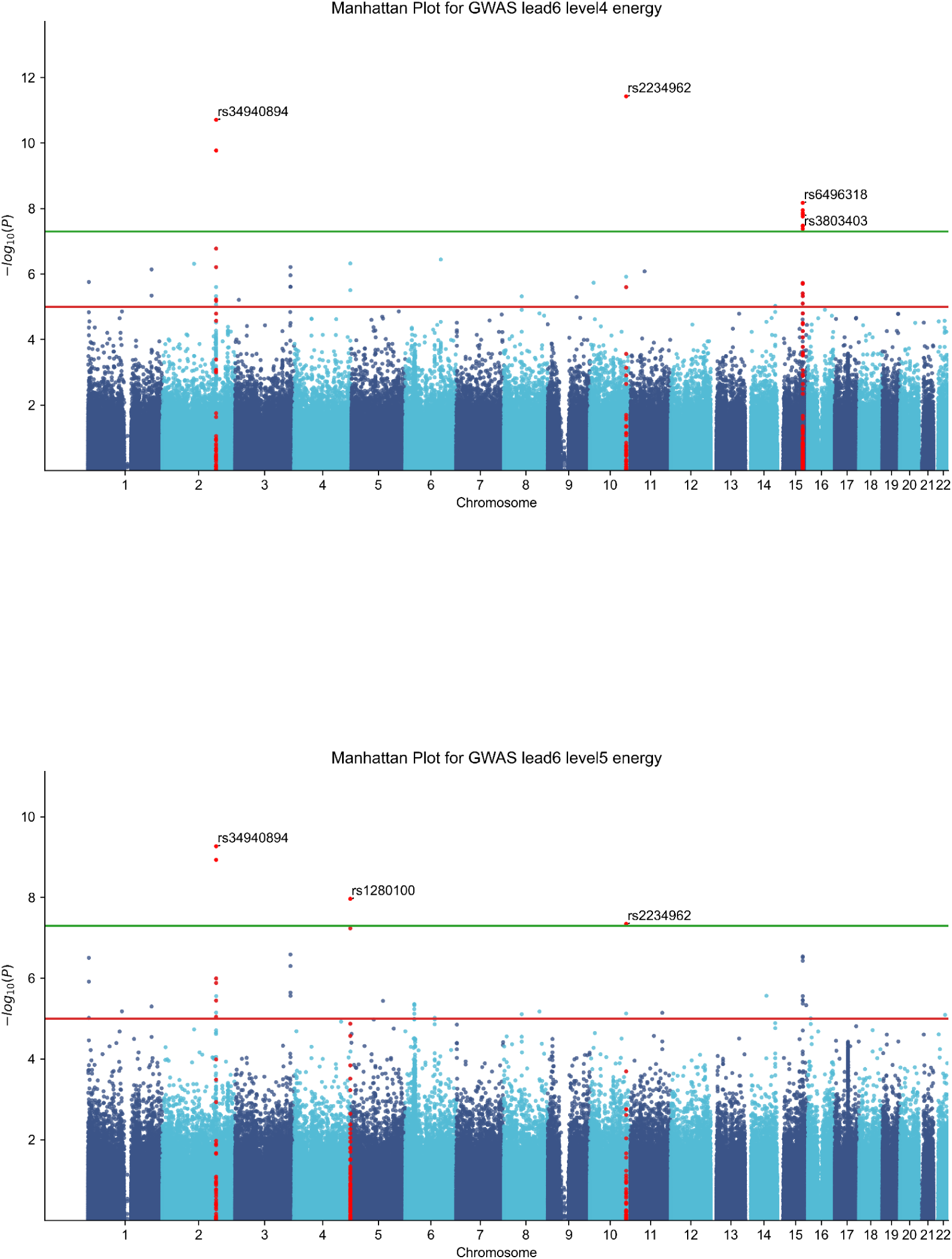
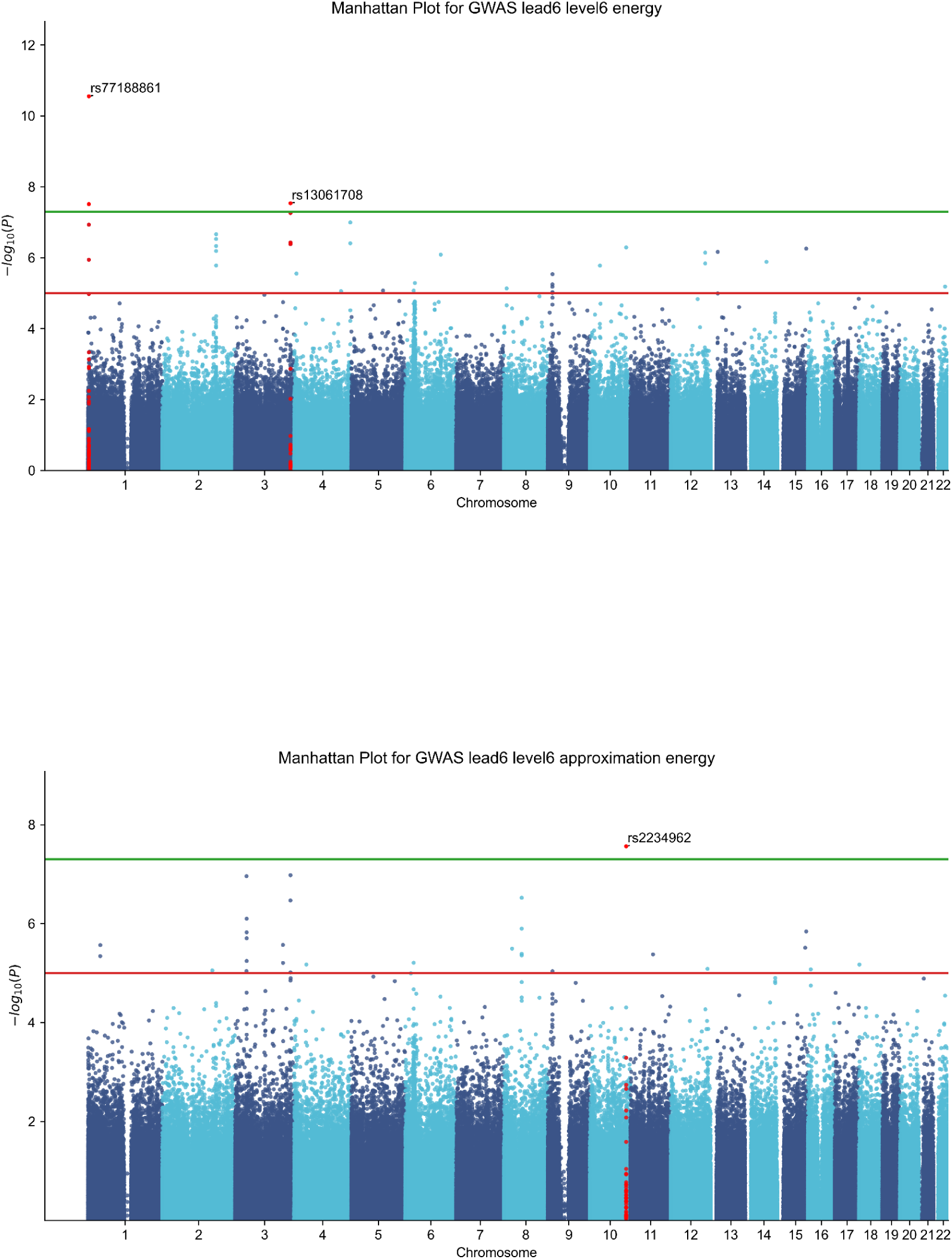
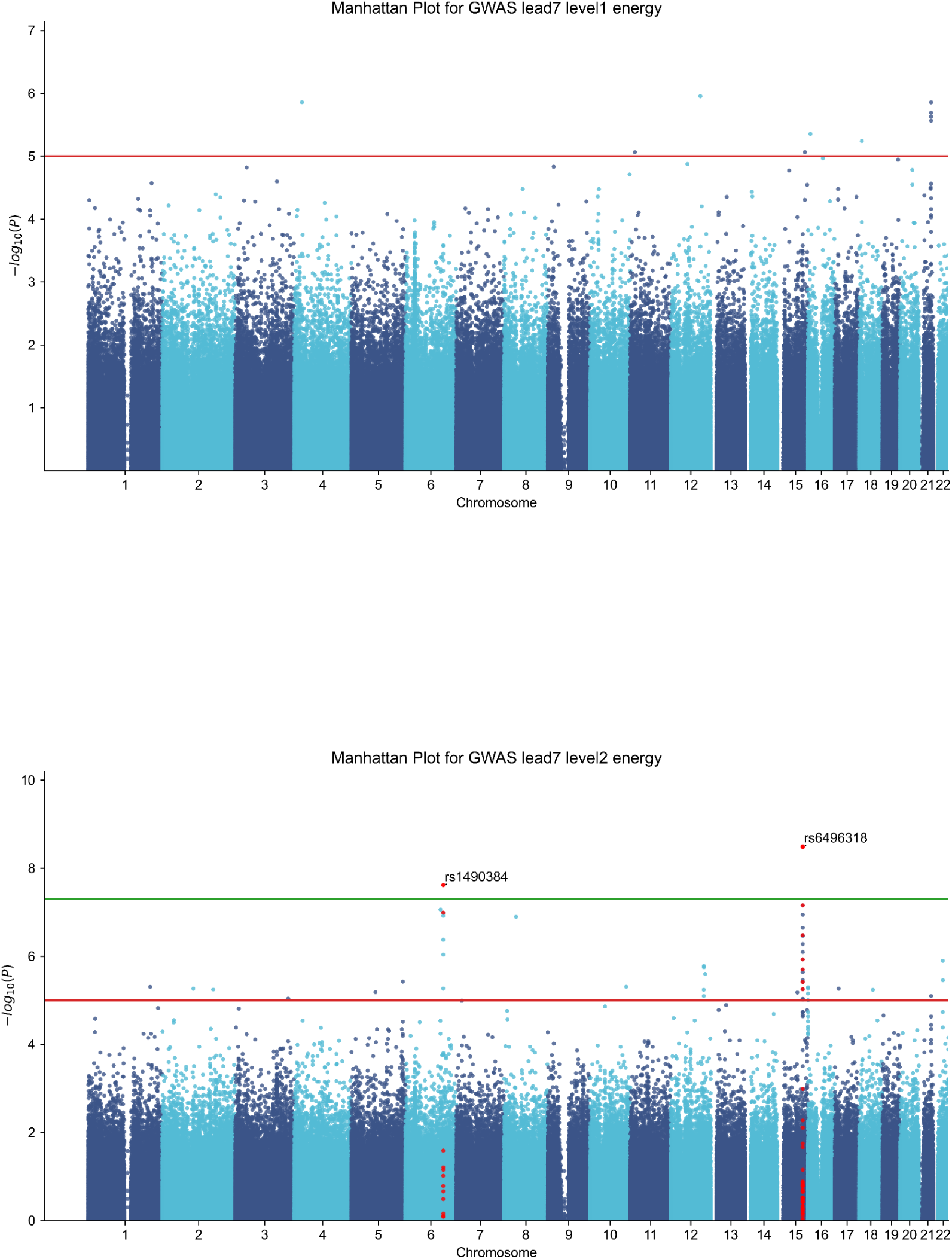
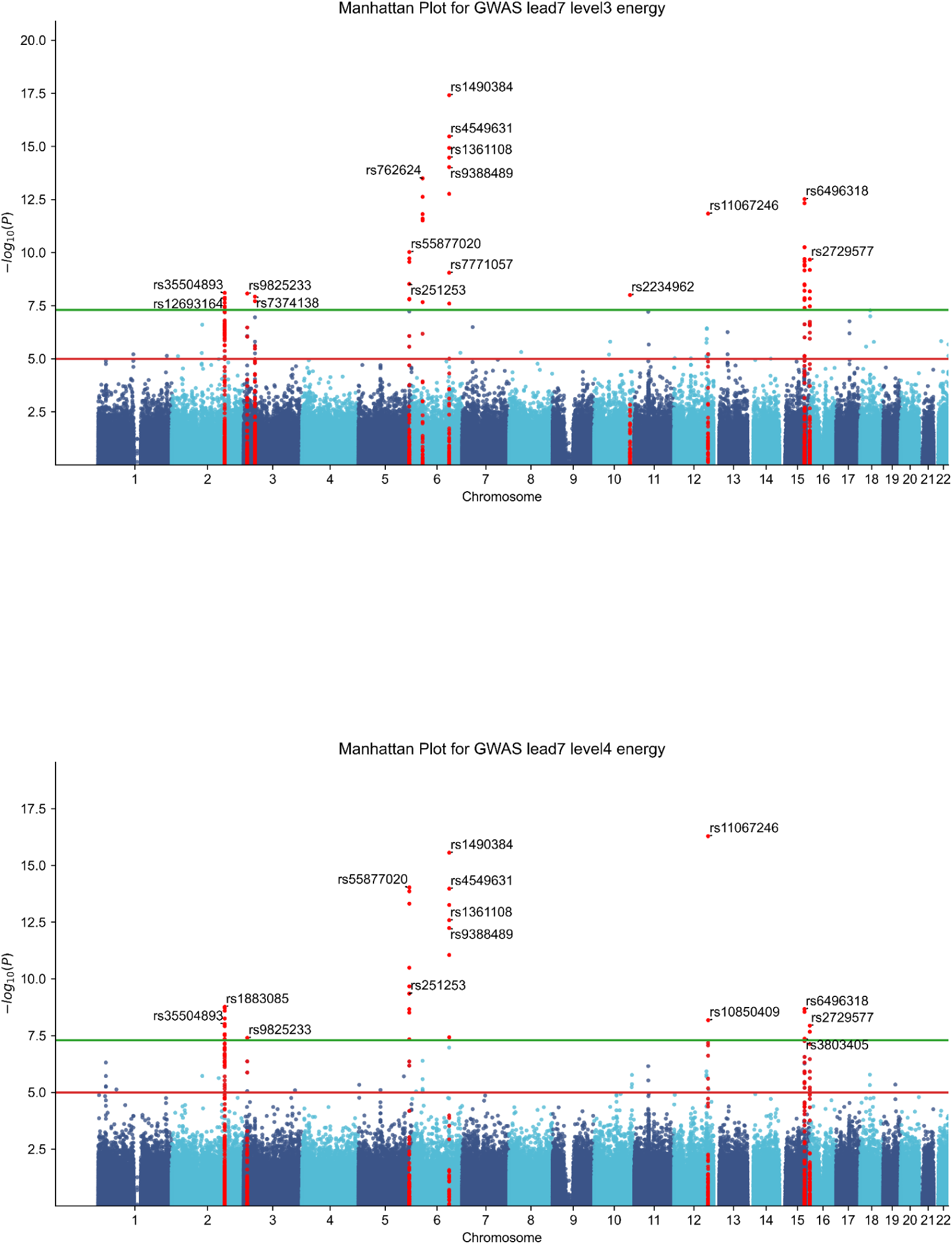
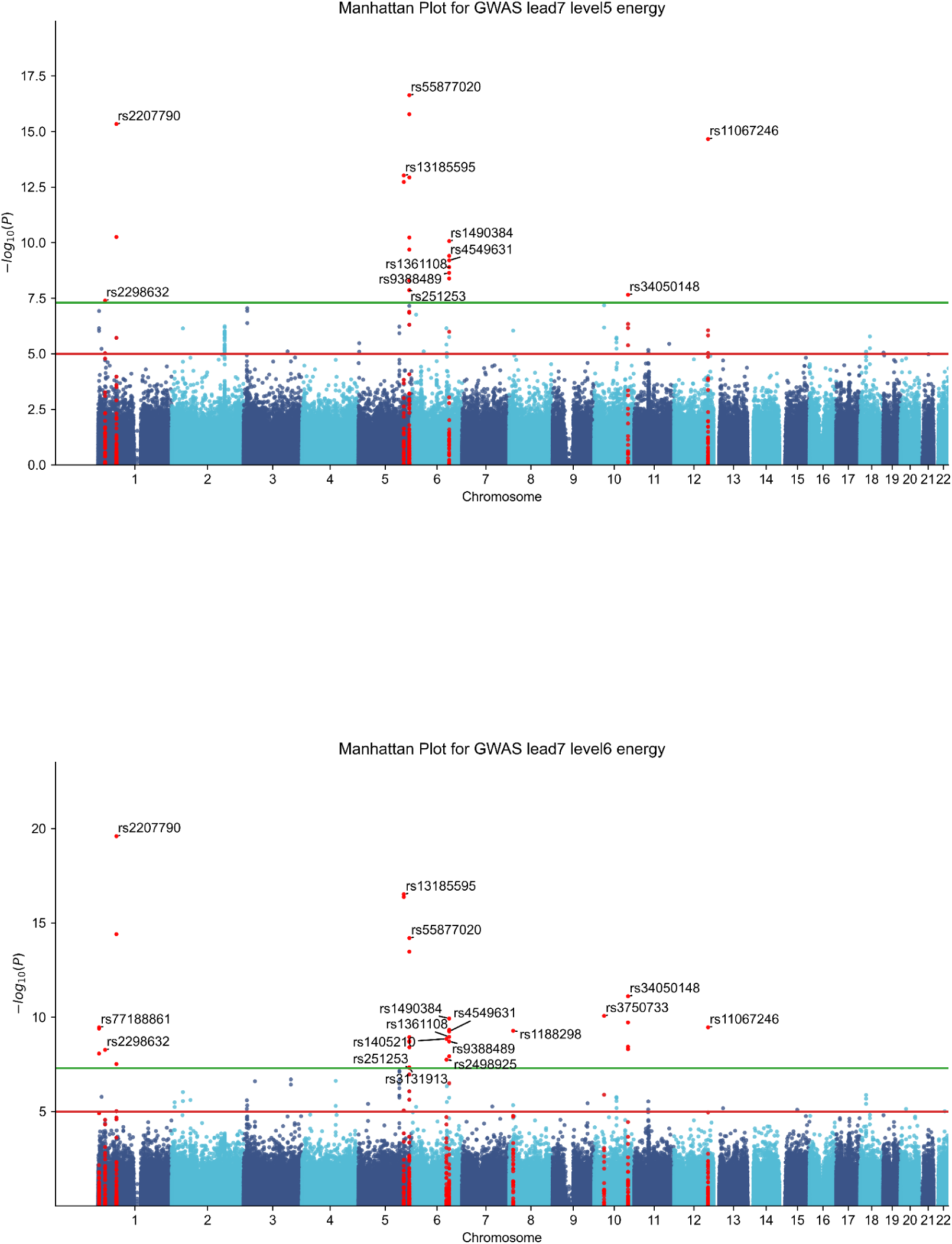
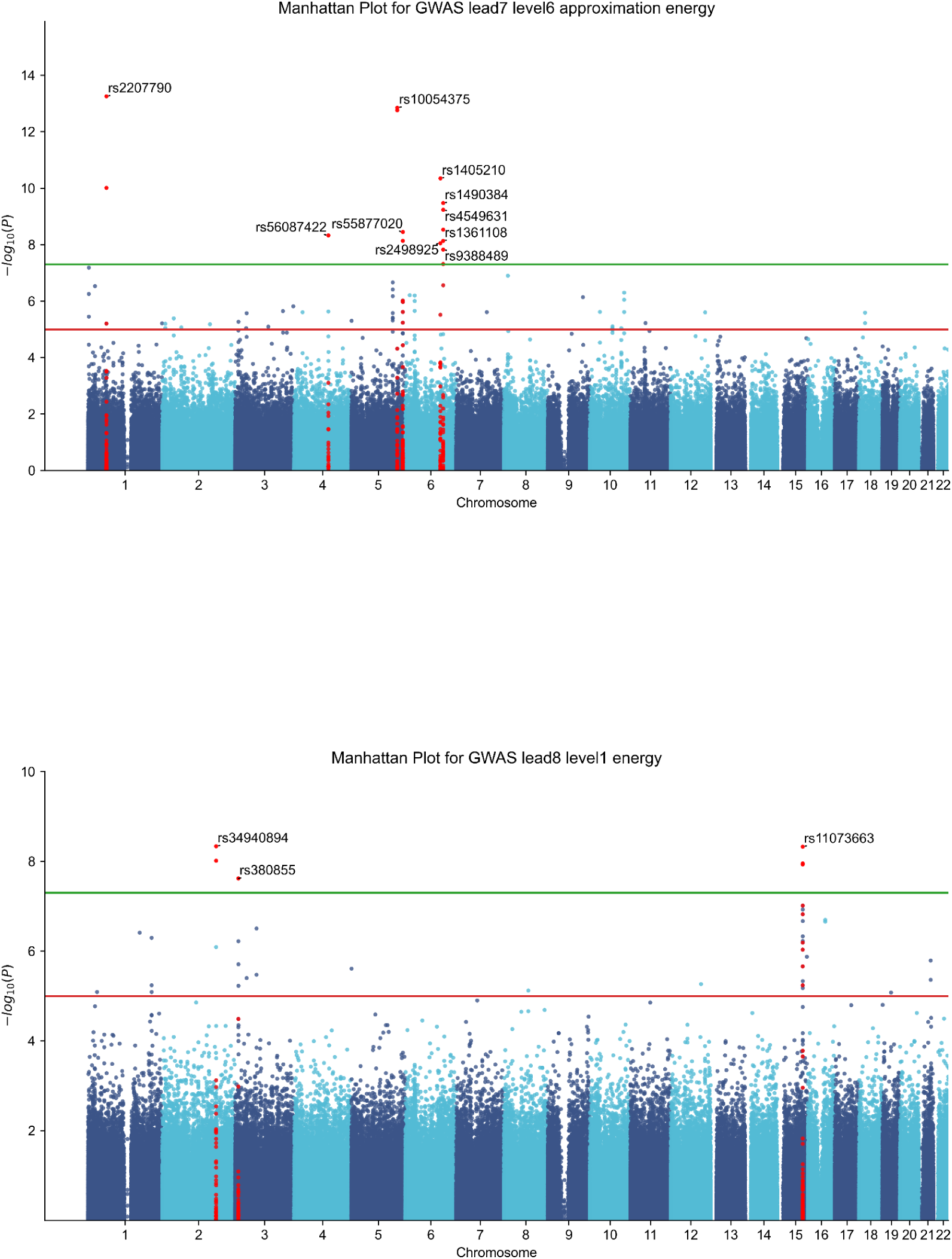
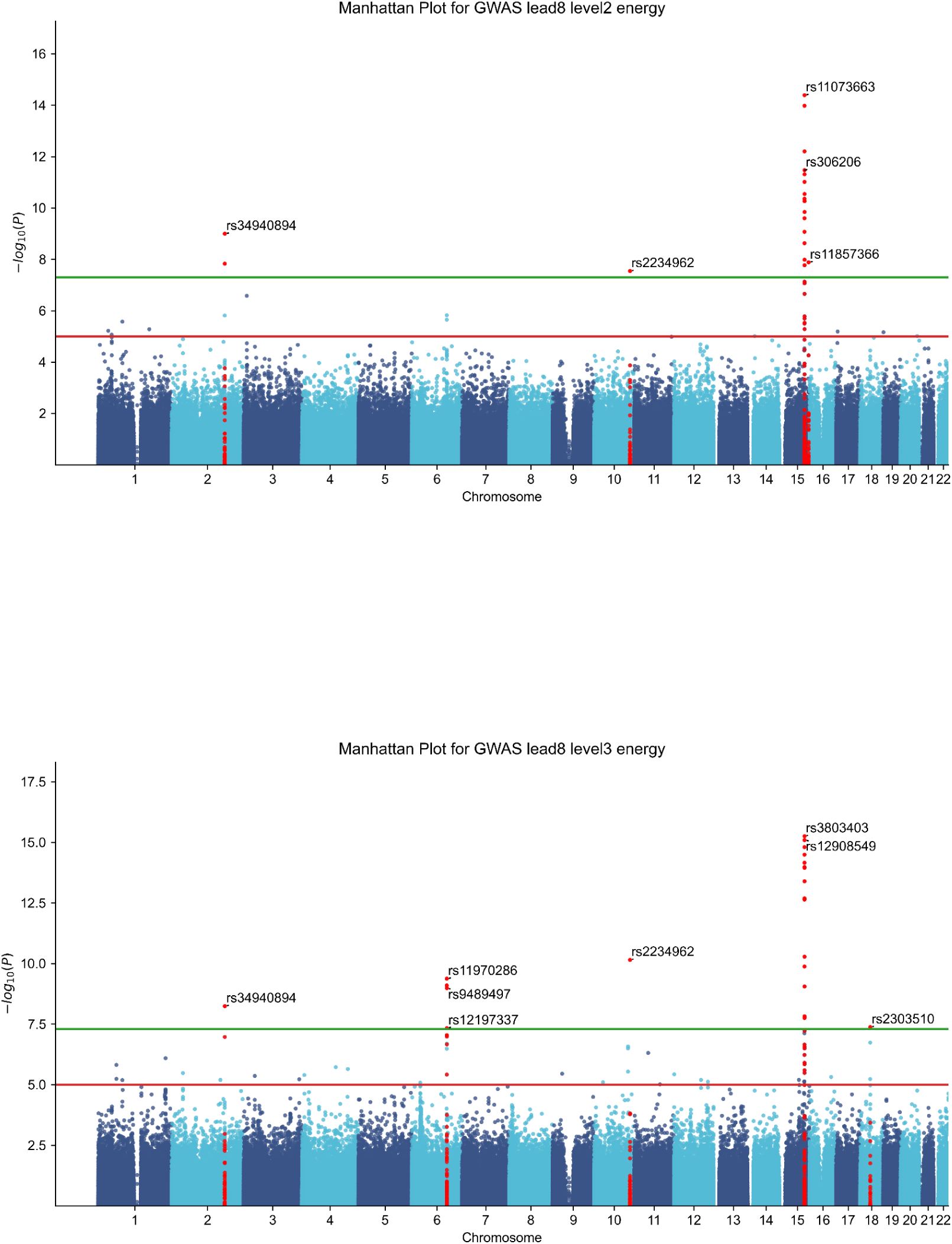
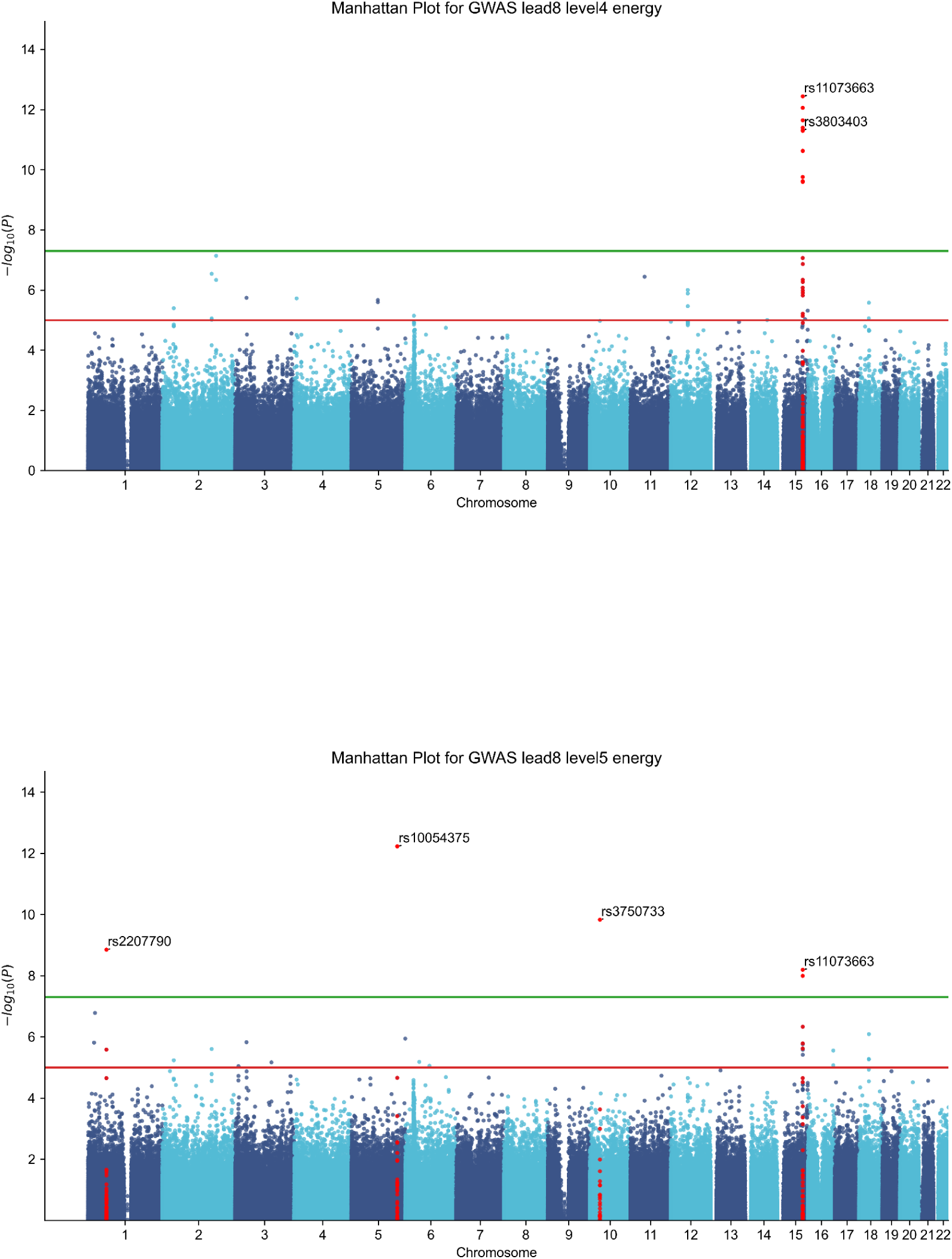
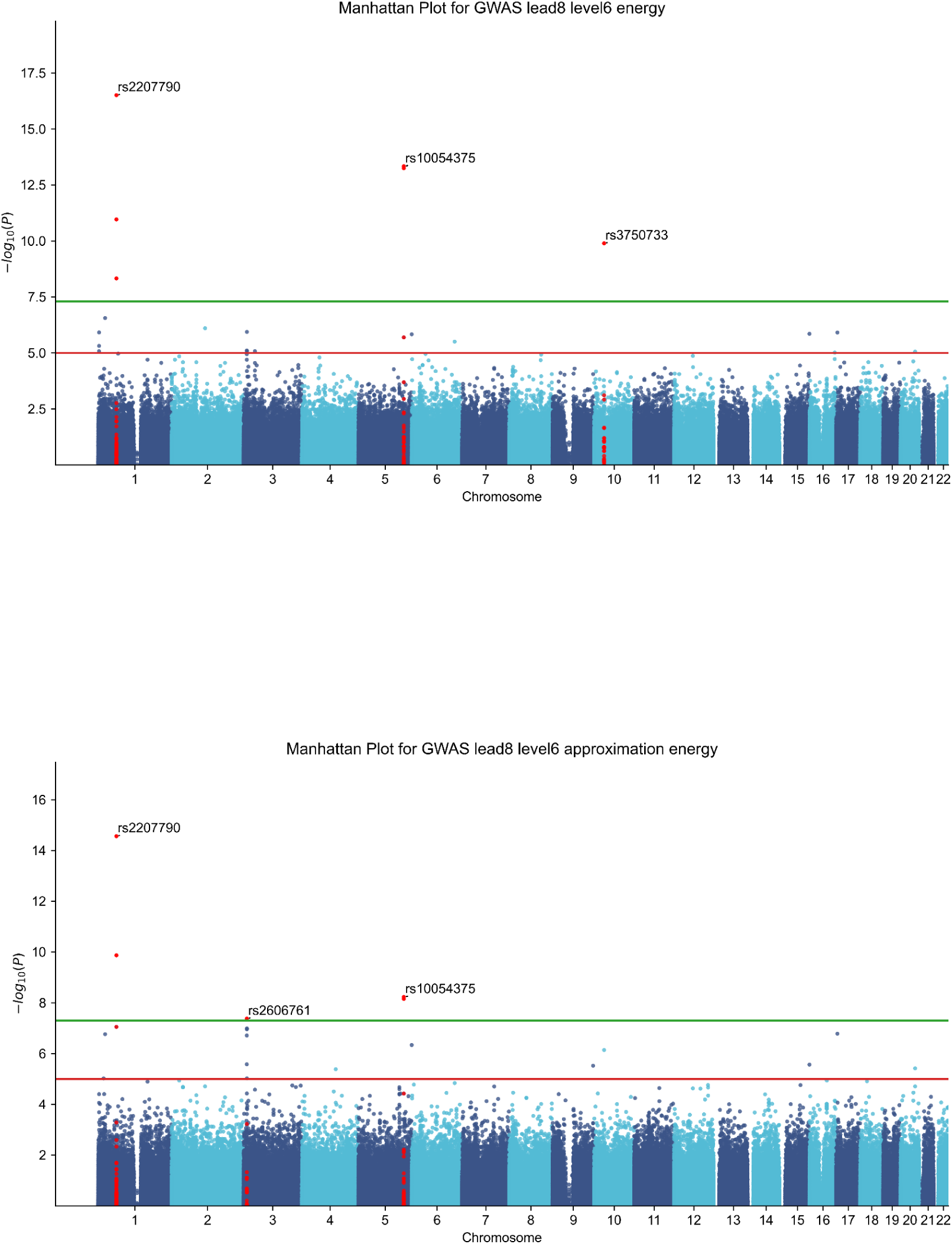
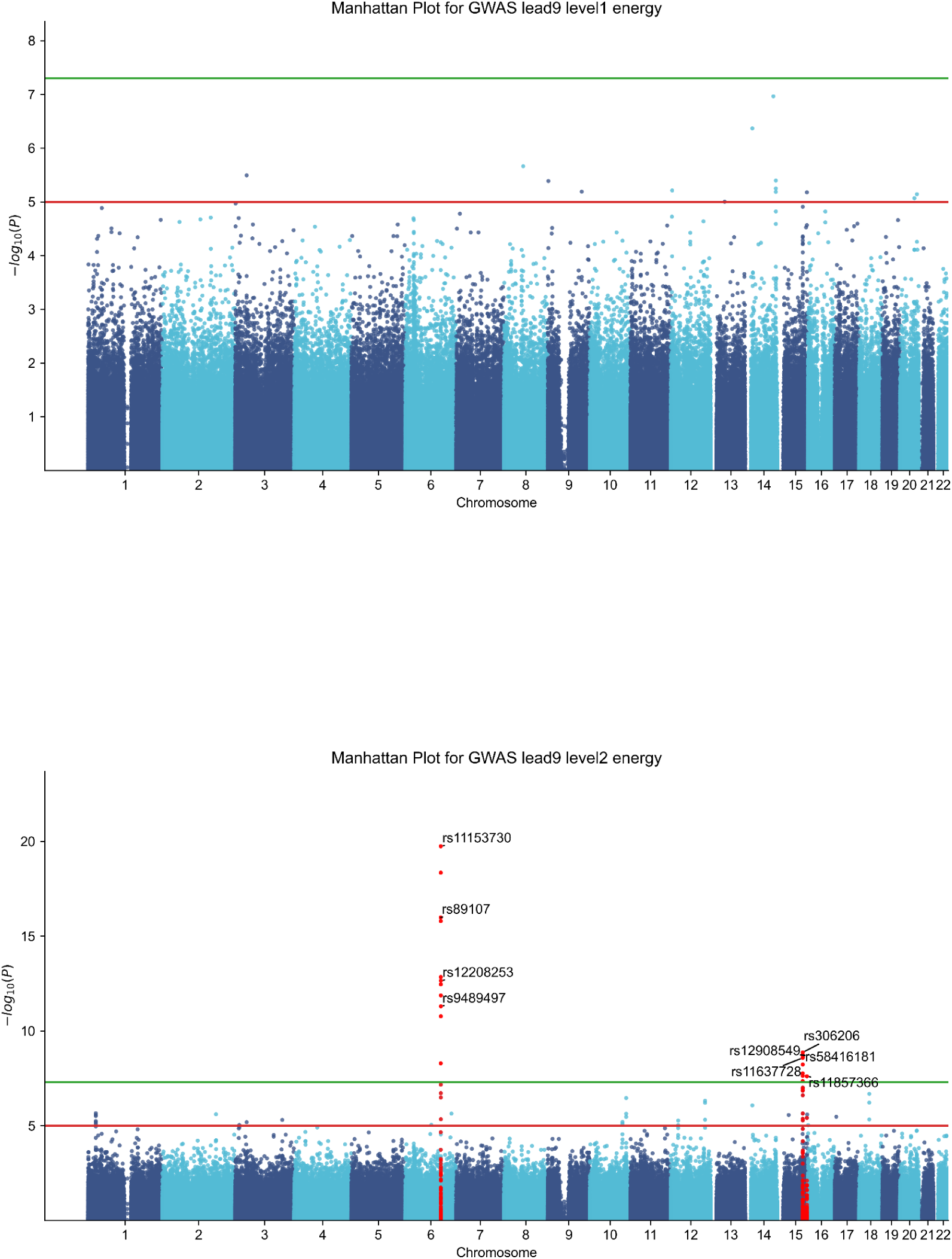
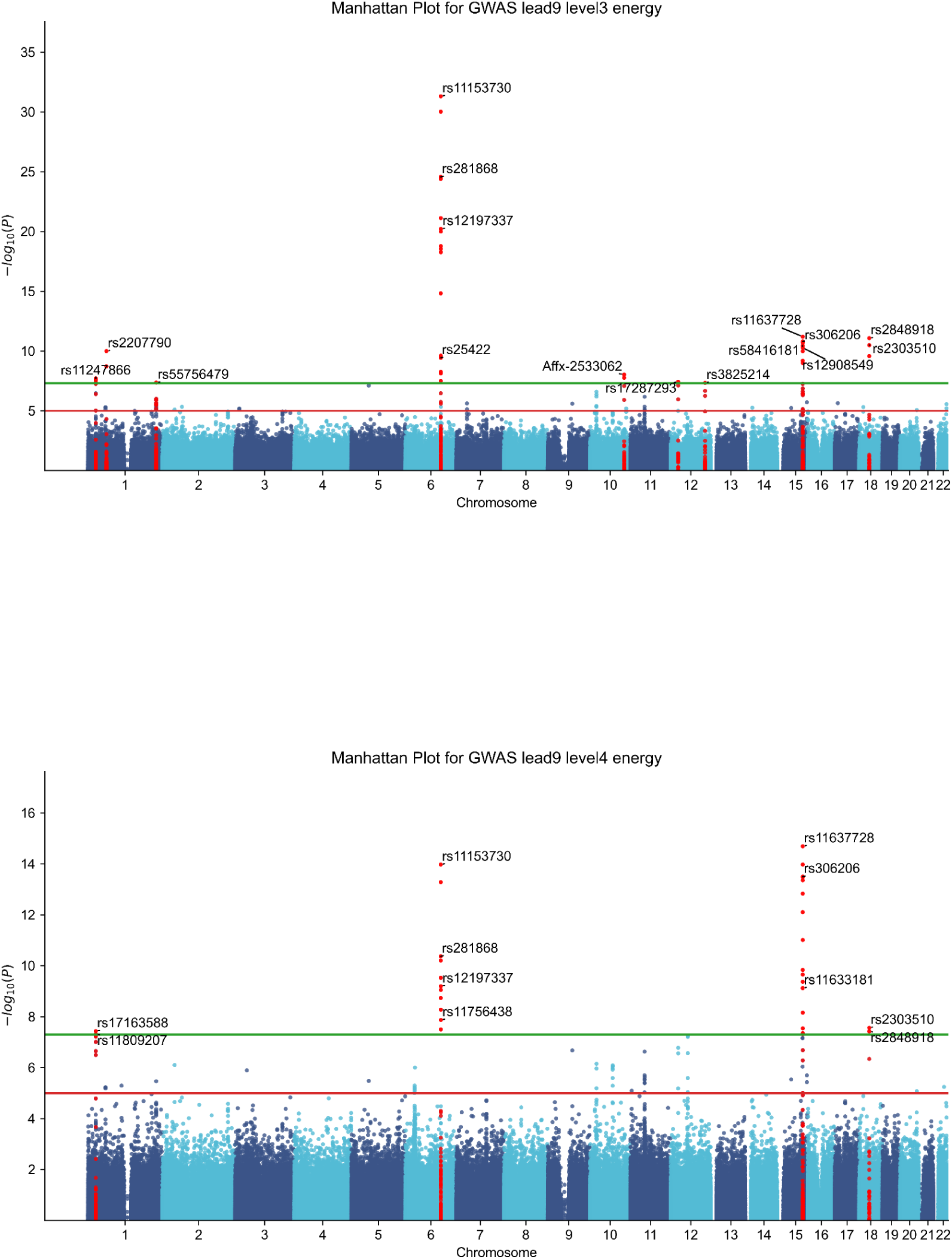
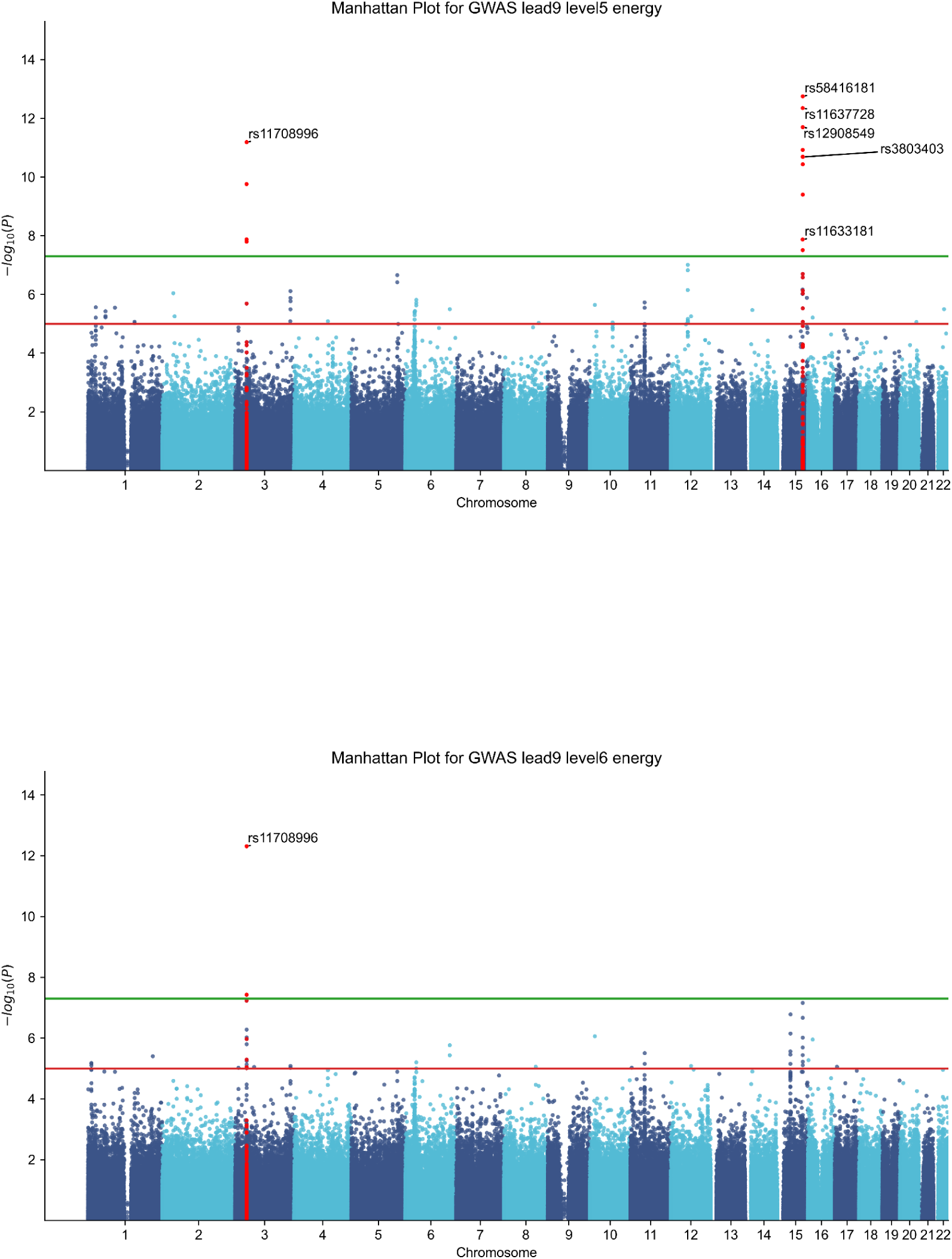
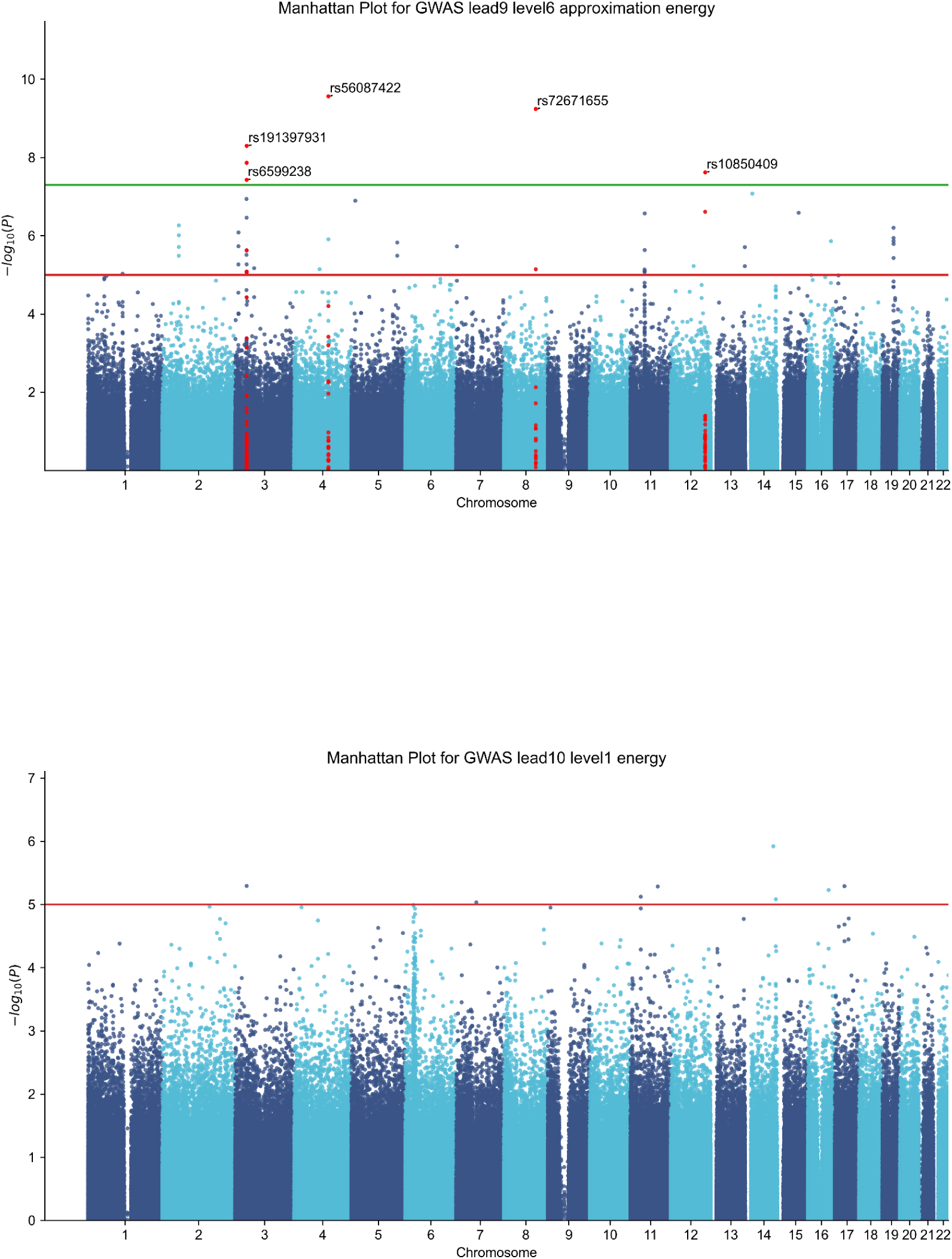
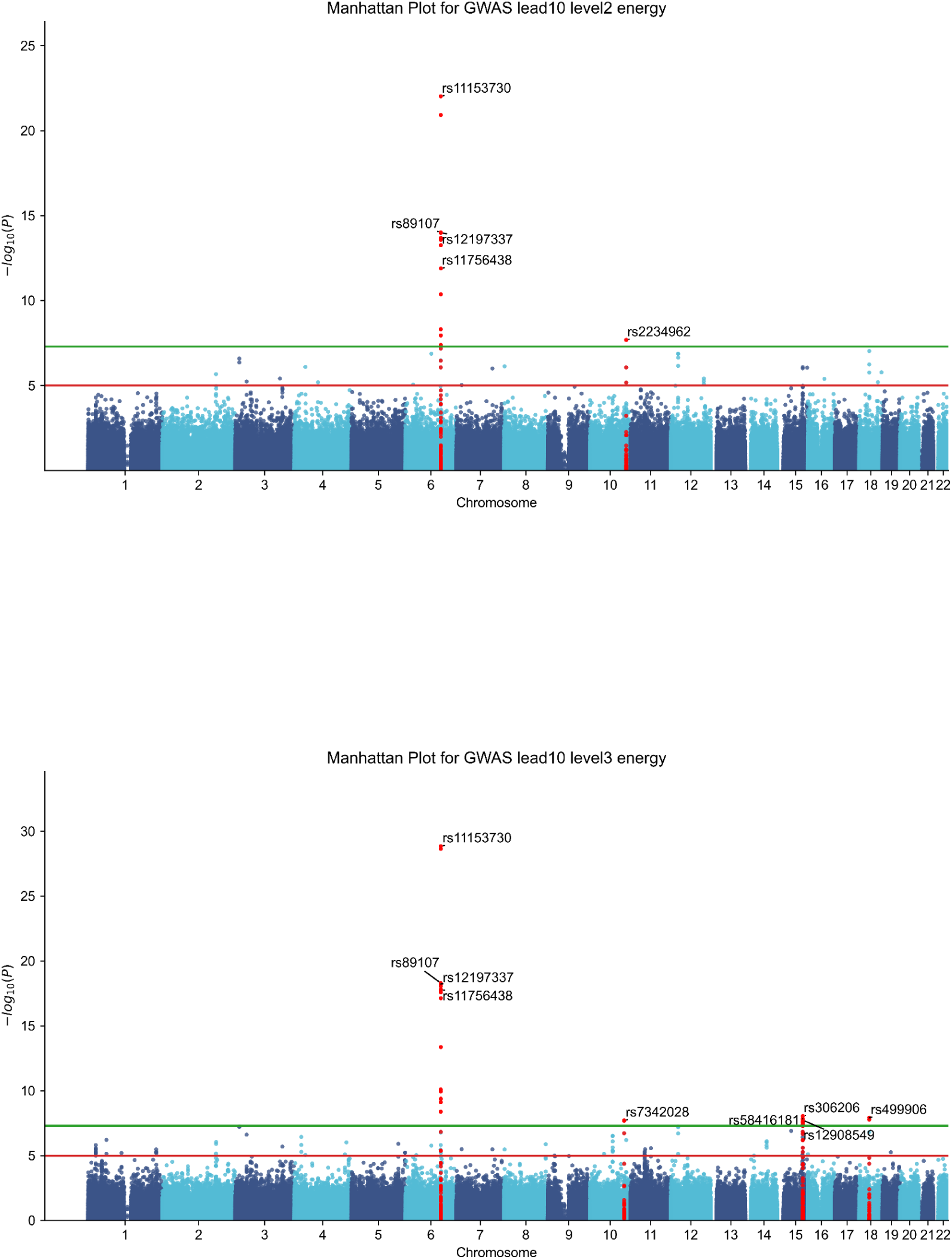
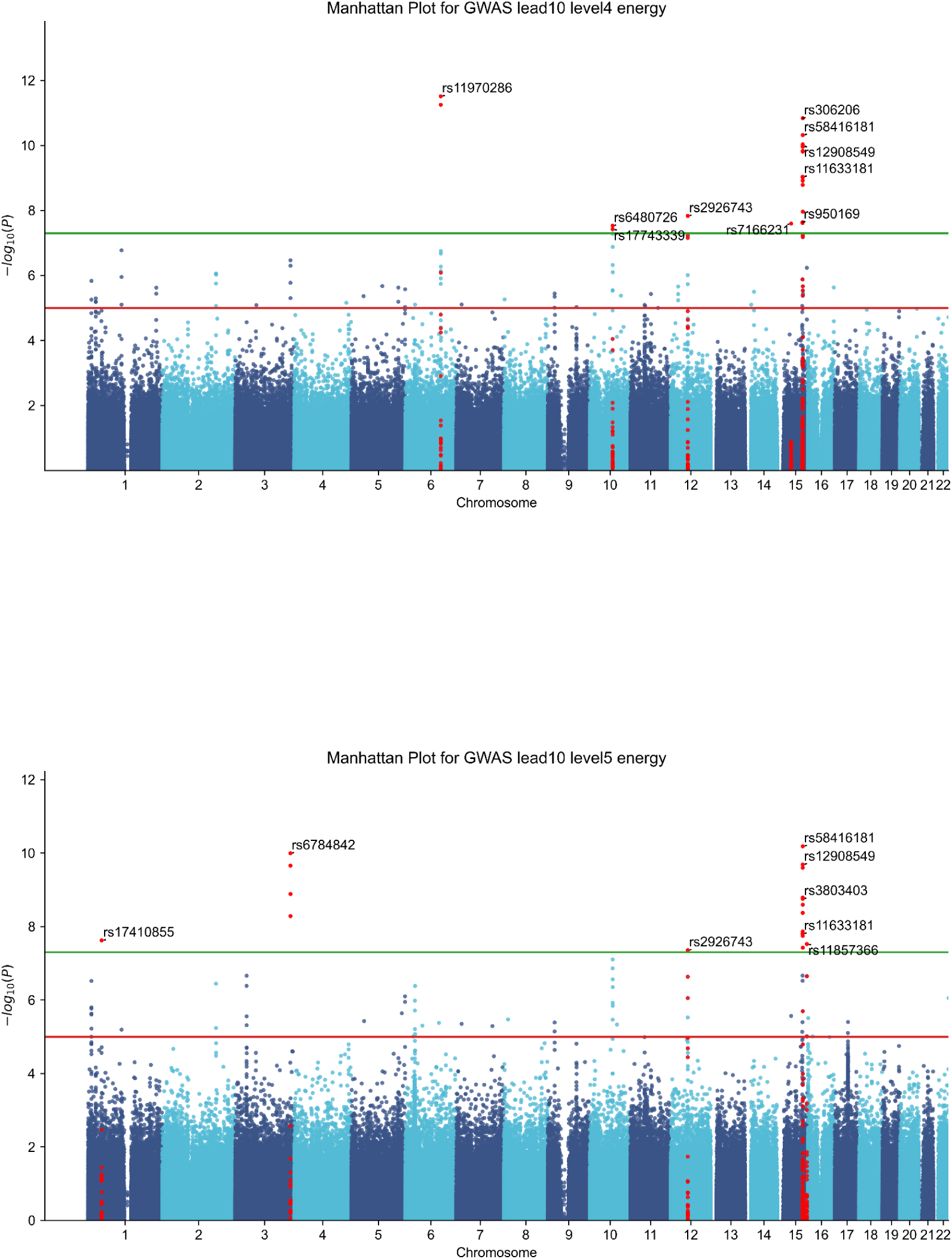
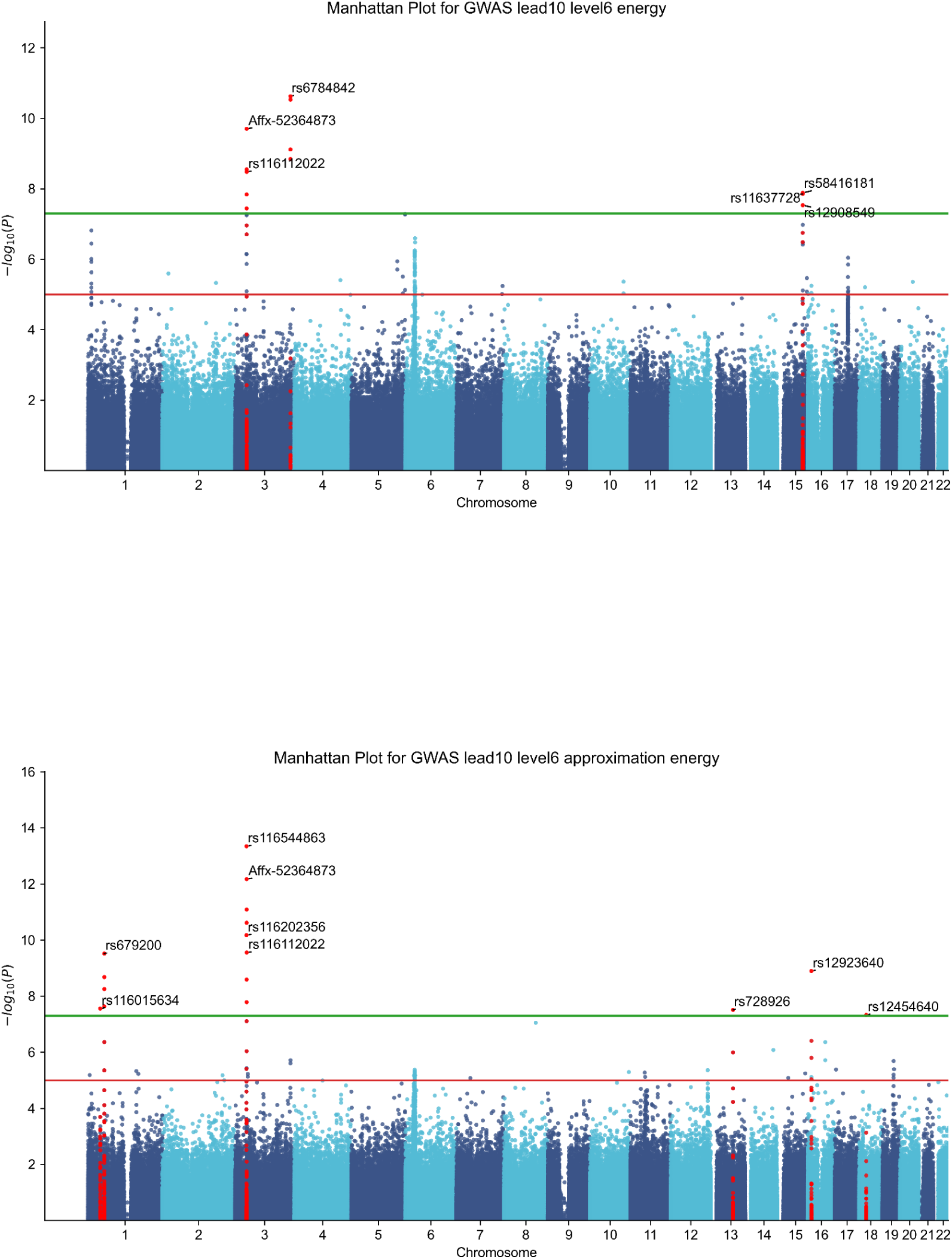
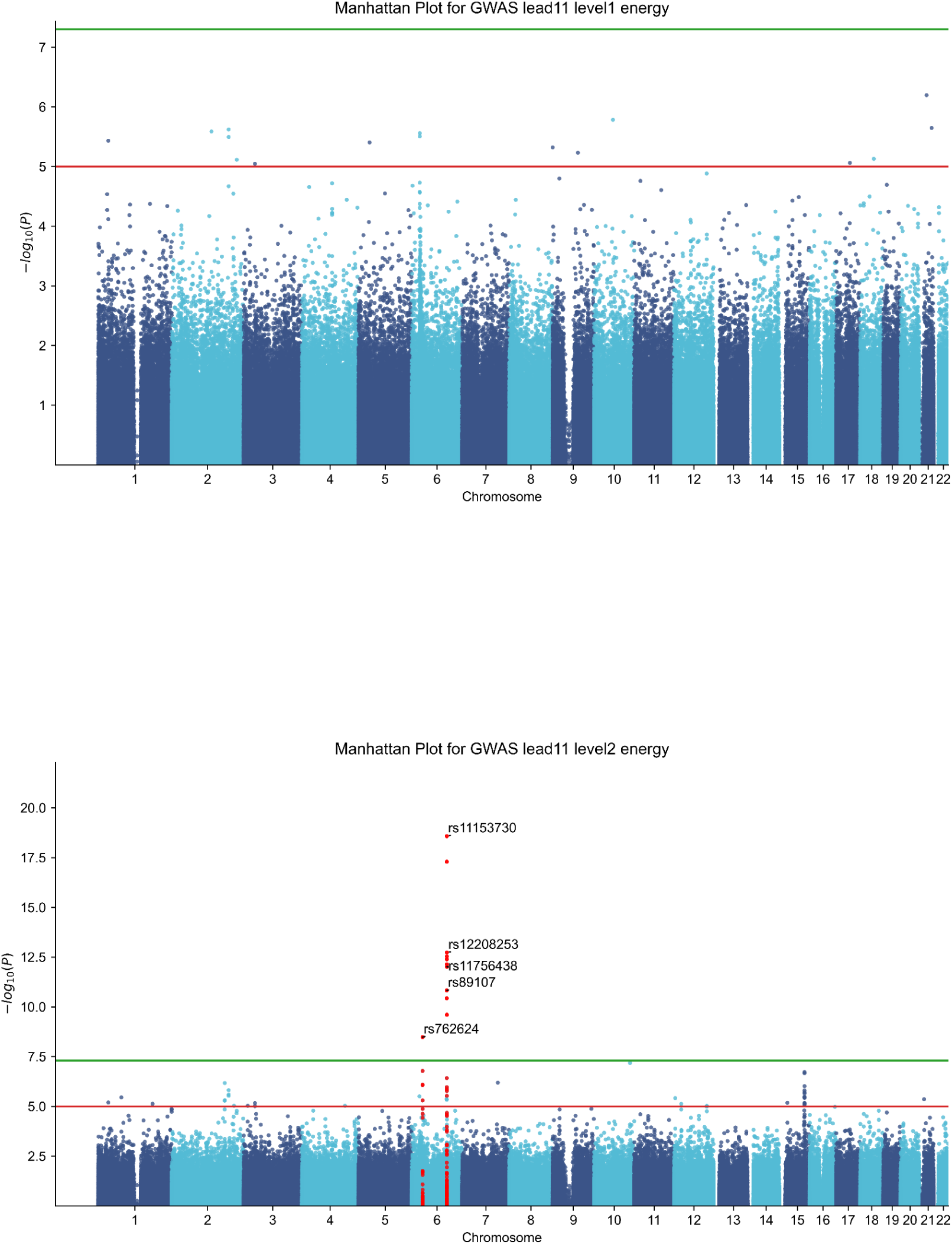
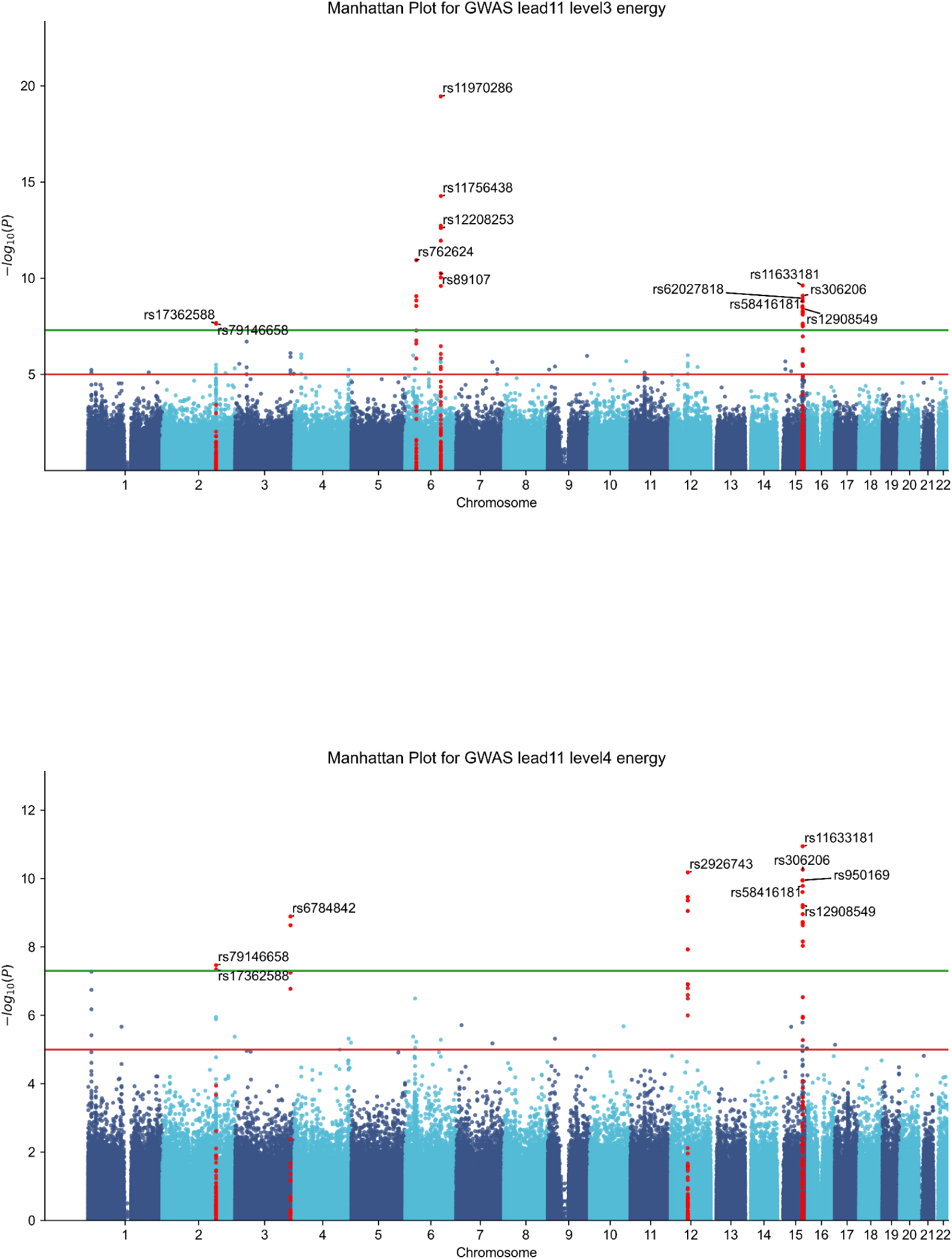
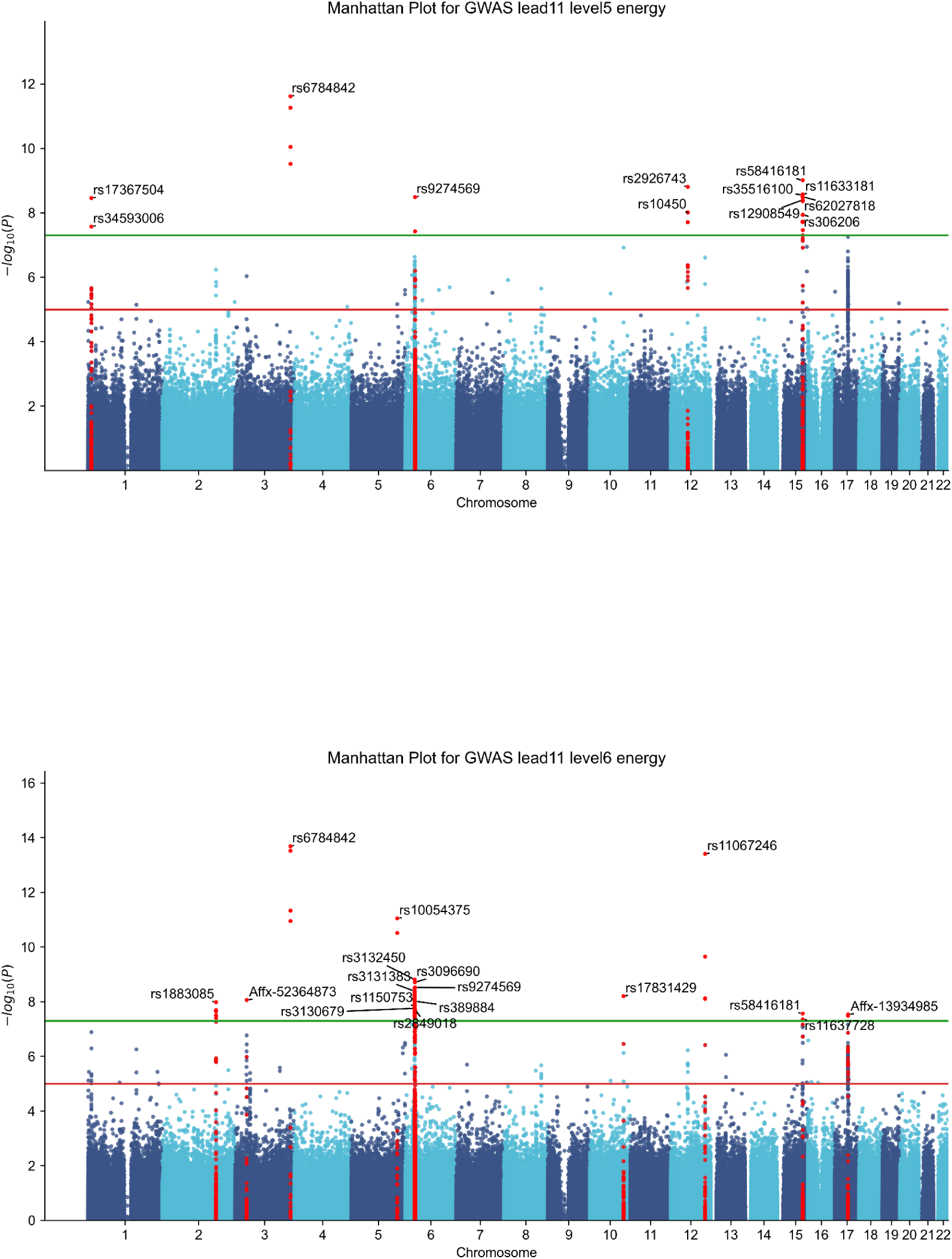
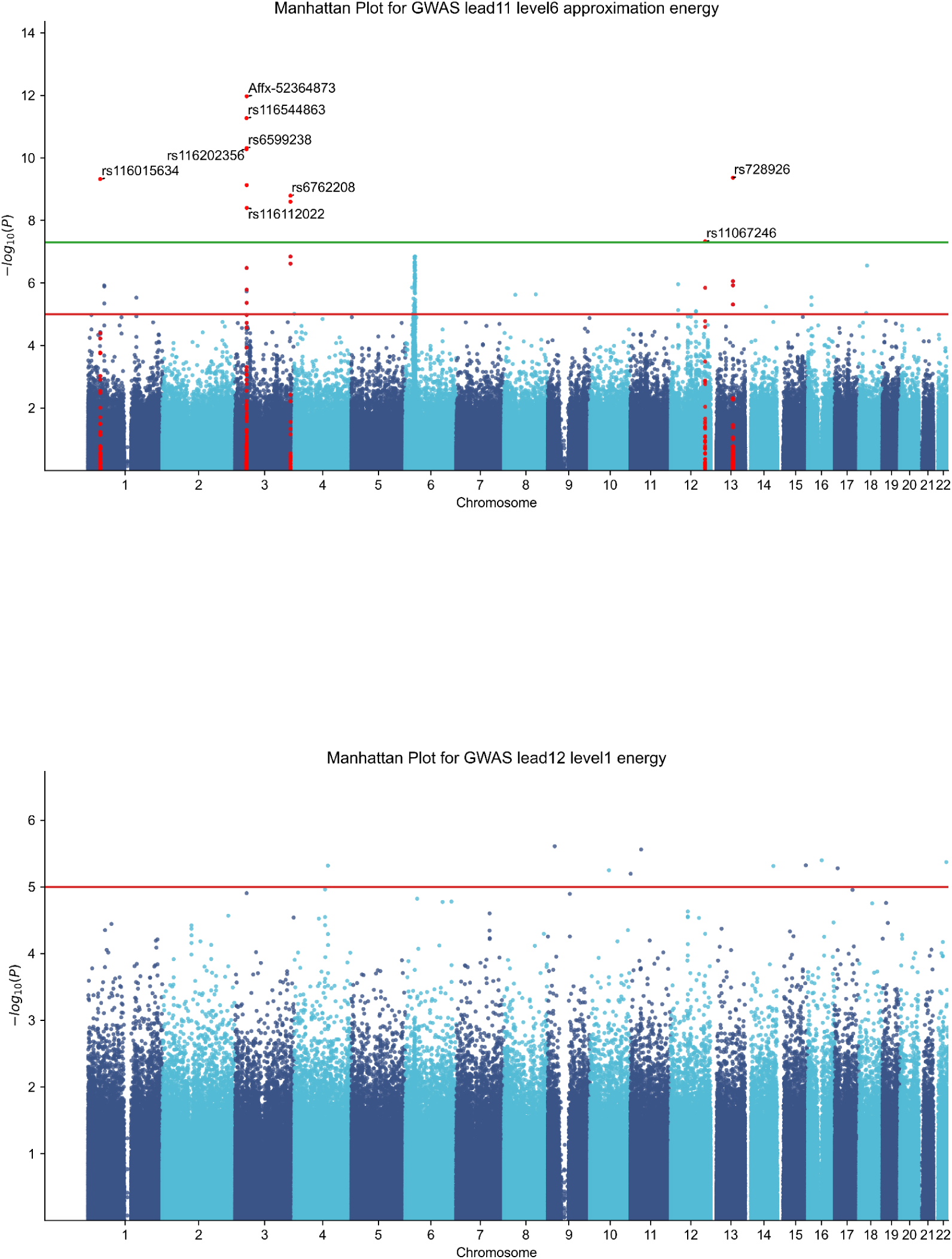
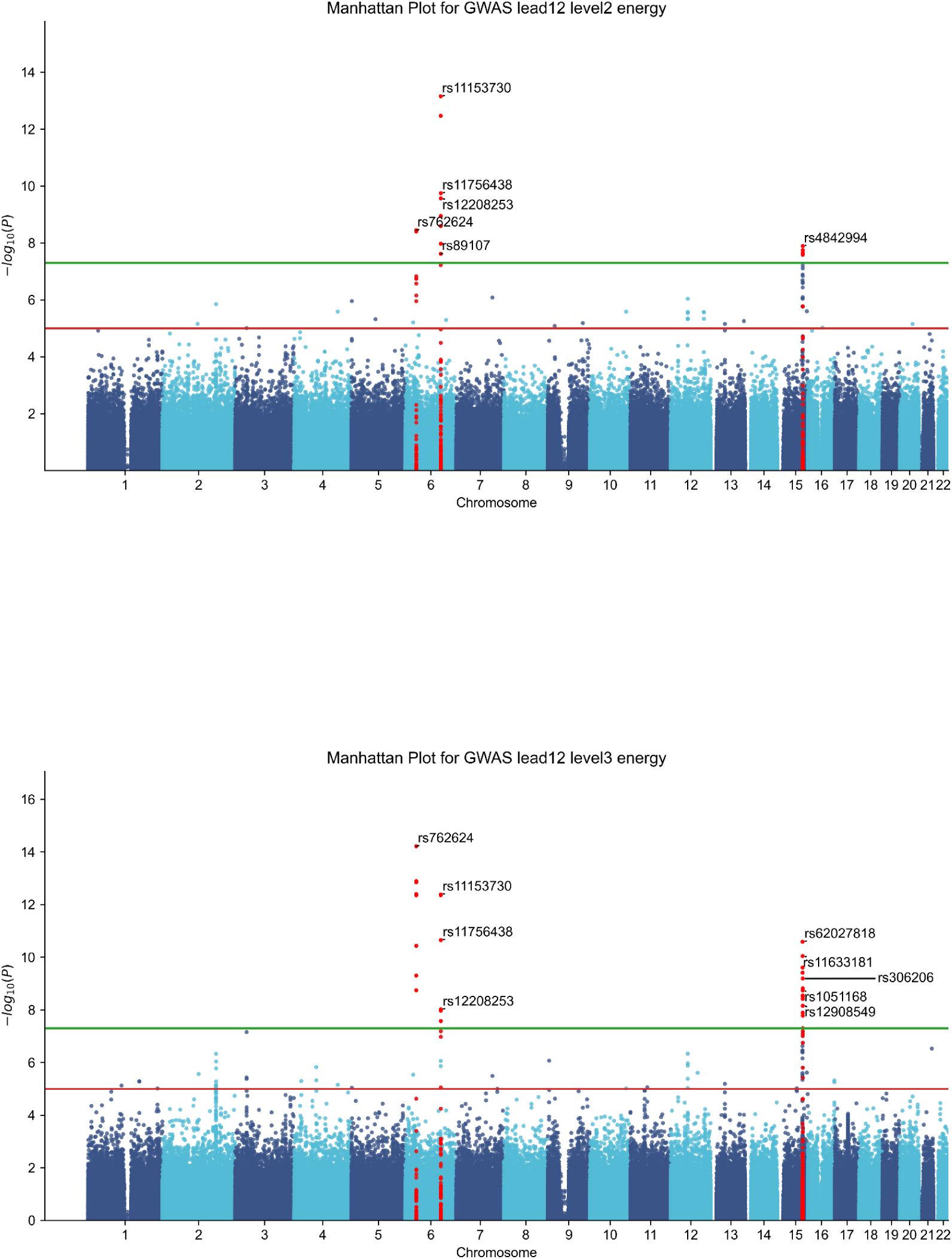
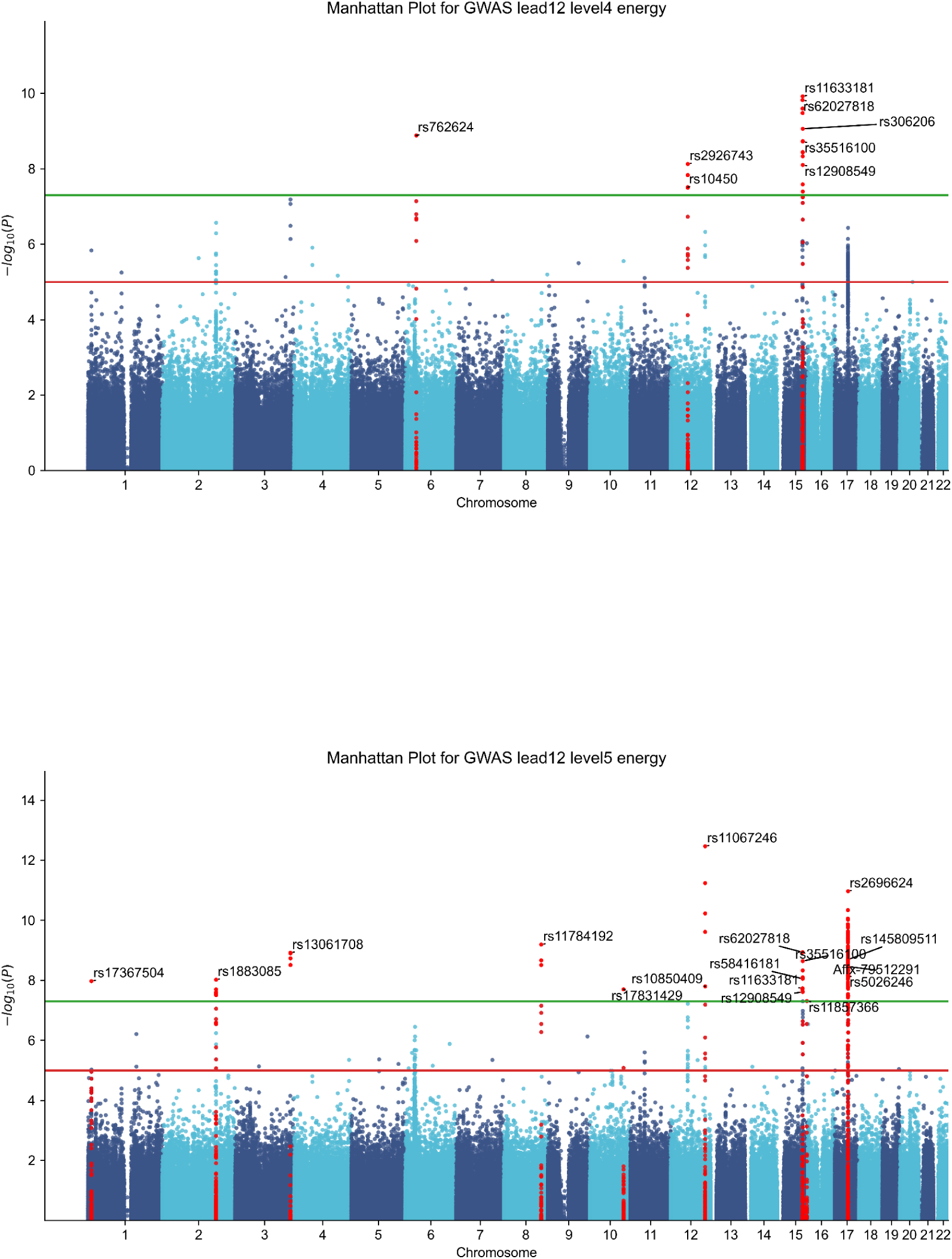
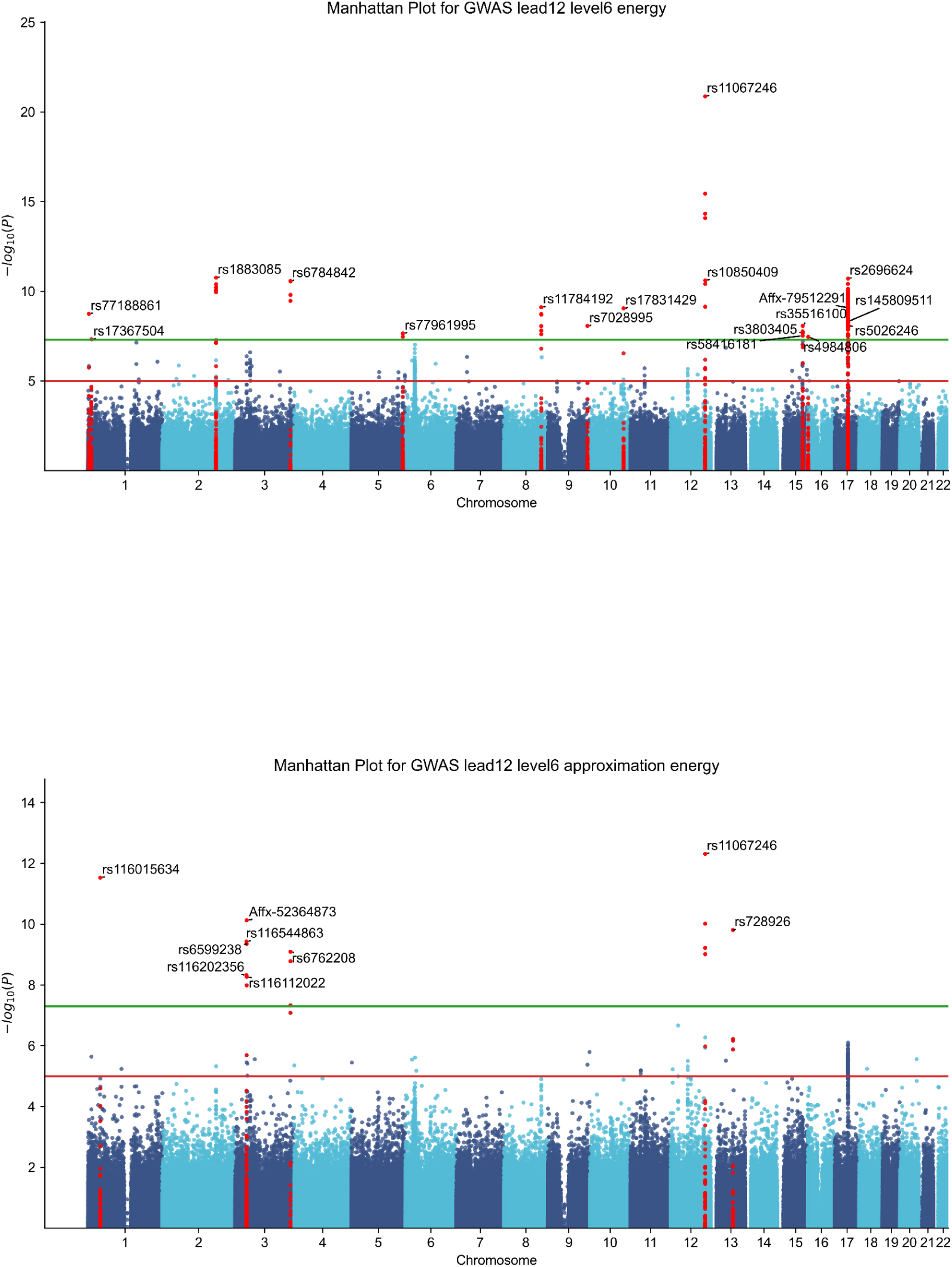
Figures S1 present the complete set of Manhattan plots from the genome-wide association analyses of all 84 wavelet-derived ECG energy features (12 leads × 6 detail levels + 1 approximation level). Each plot displays the chromosomal position of SNPs versus their association significance (–log₁₀(p)), with genome-wide and suggestive thresholds indicated. These figures complement the main Manhattan plot shown in Figure 2 by providing the full visual results for each lead and decomposition level, enabling detailed inspection of frequency- and lead-specific genetic association patterns.

**Supplementary Figure 2.**
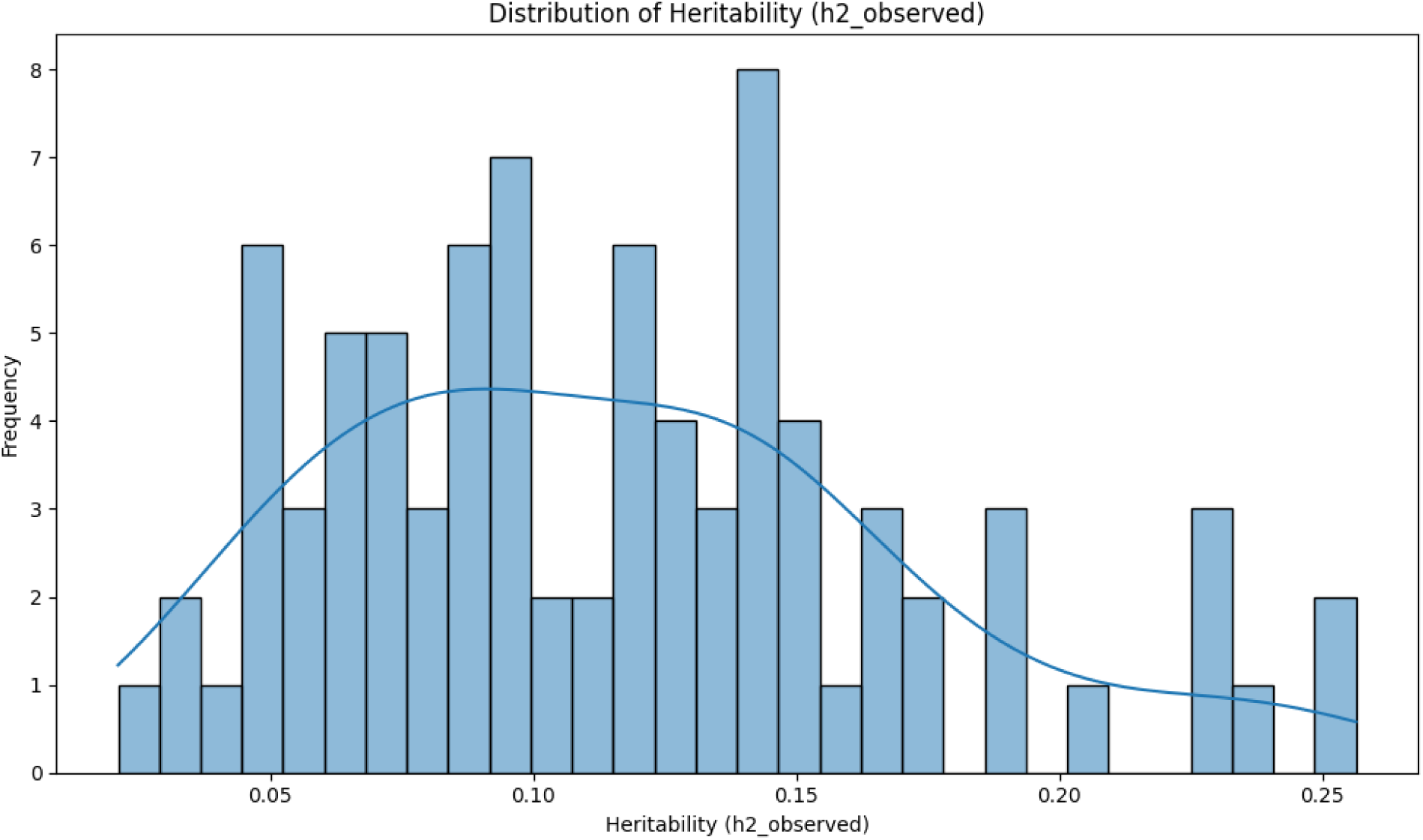
Distribution of the heritability of ECG-derived energy features. The strength of the heritability is denoted on the x-axis, and the frequency is denoted on the y-axis.

The distribution of observed heritability (h²) estimates across the 84 wavelet-derived energy features is depicted in the histogram, revealing a unimodal and right-skewed pattern with values ranging from below 0.03 to a maximum of 0.25. The majority of estimates cluster in the moderate range of 0.05–0.20, with a peak frequency around 0.12–0.15. These h² values align with GWAS-based heritability estimates for standard ECG traits, such as QRS duration (∼0.16–0.17) and QT interval (∼0.17). The right-skewed tail towards higher h² (up to 0.25) suggests a subset of features, particularly those in mid-detail levels (D6–D4), captures stronger heritable signals, thereby validating the wavelet decomposition approach as a tool for dissecting nuanced genetic influences on cardiac traits beyond traditional metrics.

**Supplementary Figures 3.**
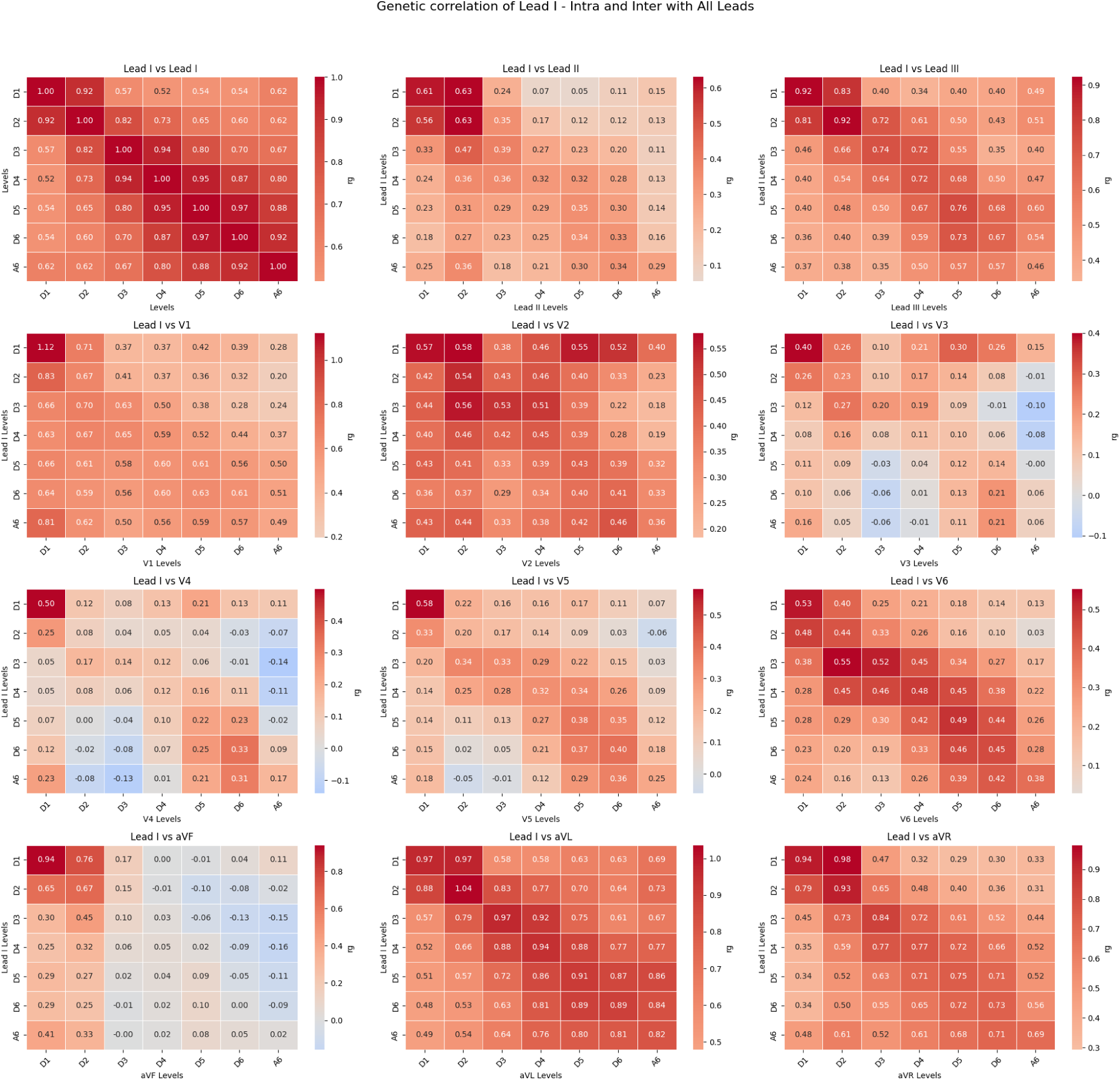
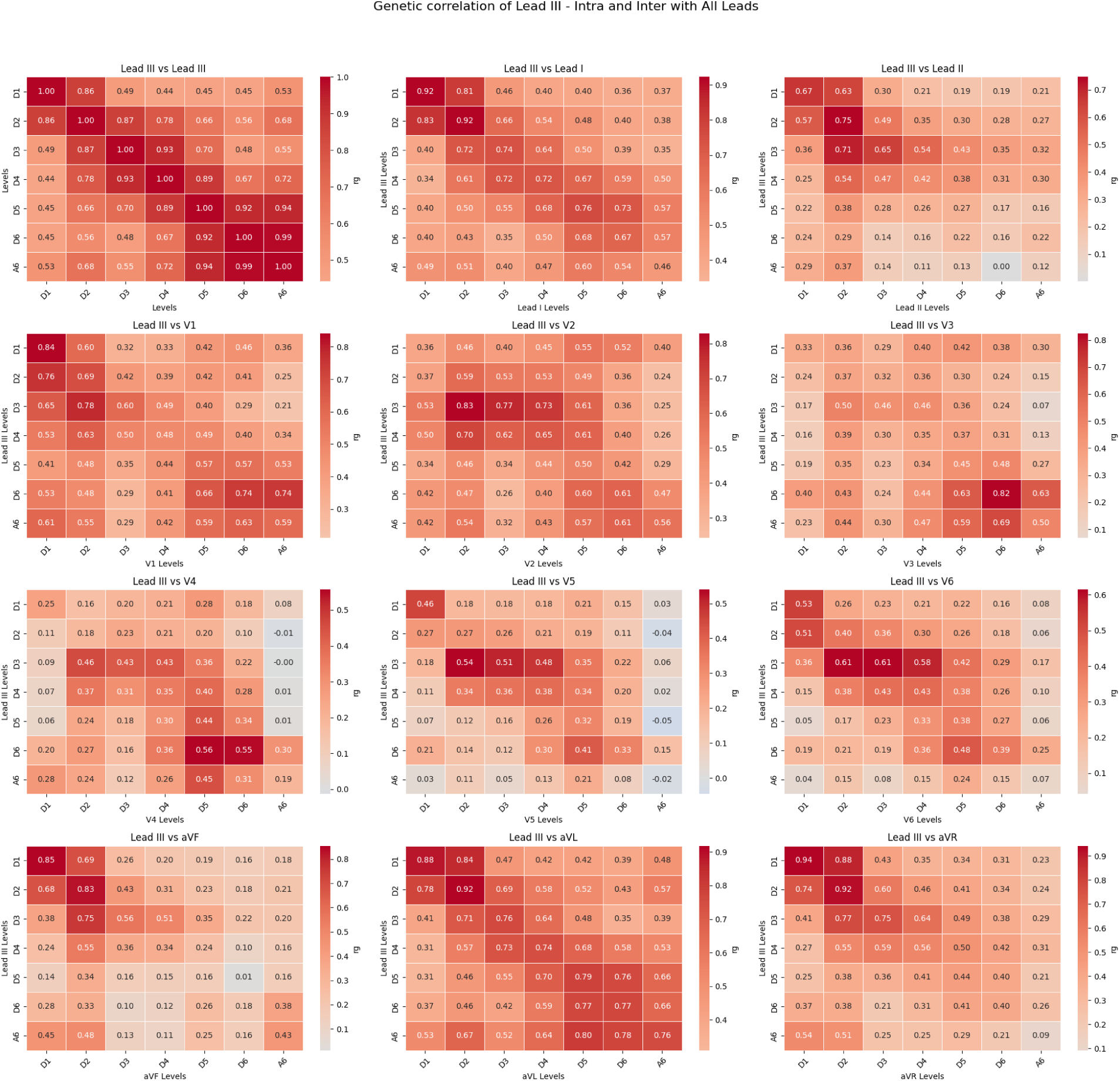
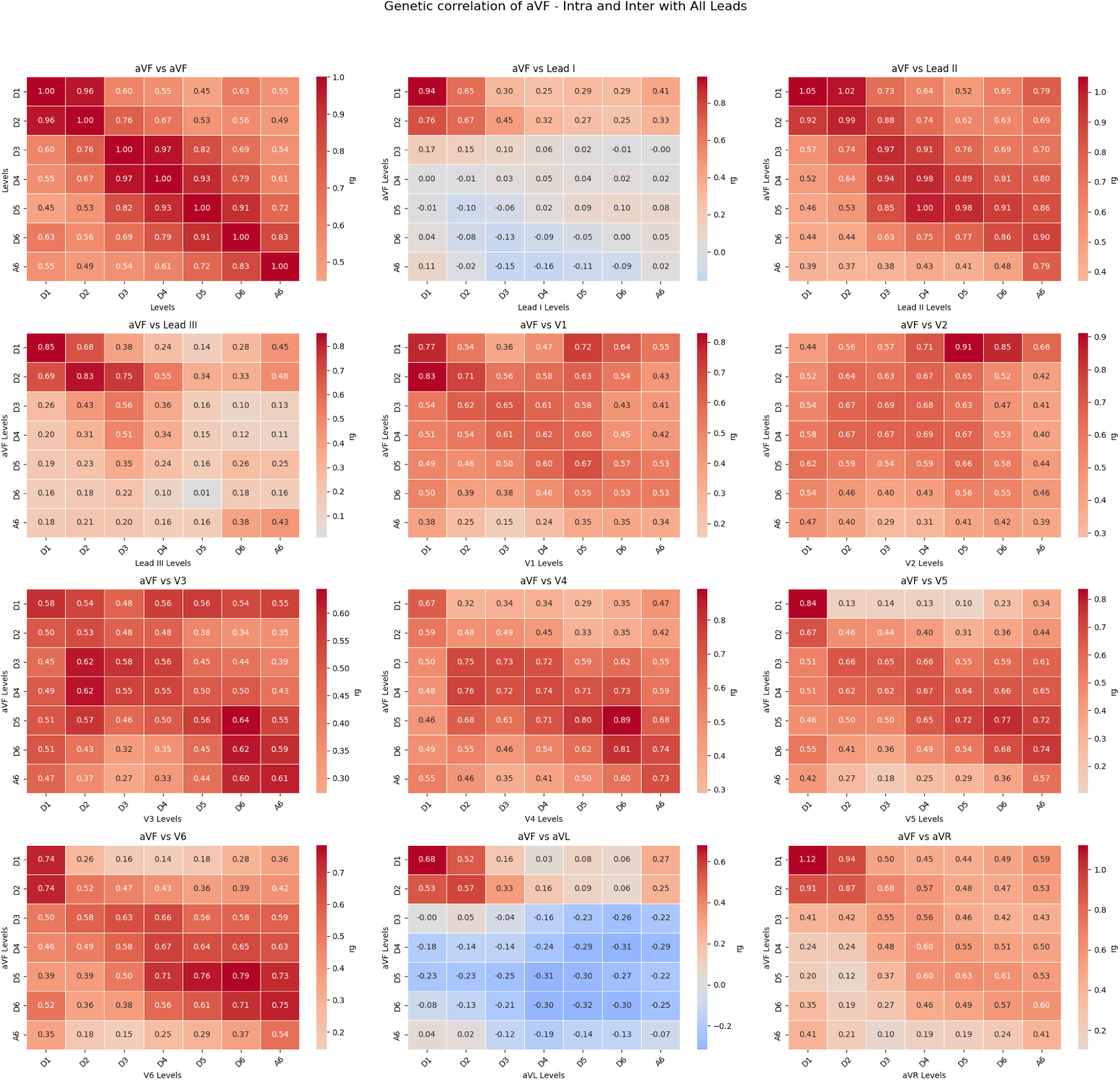
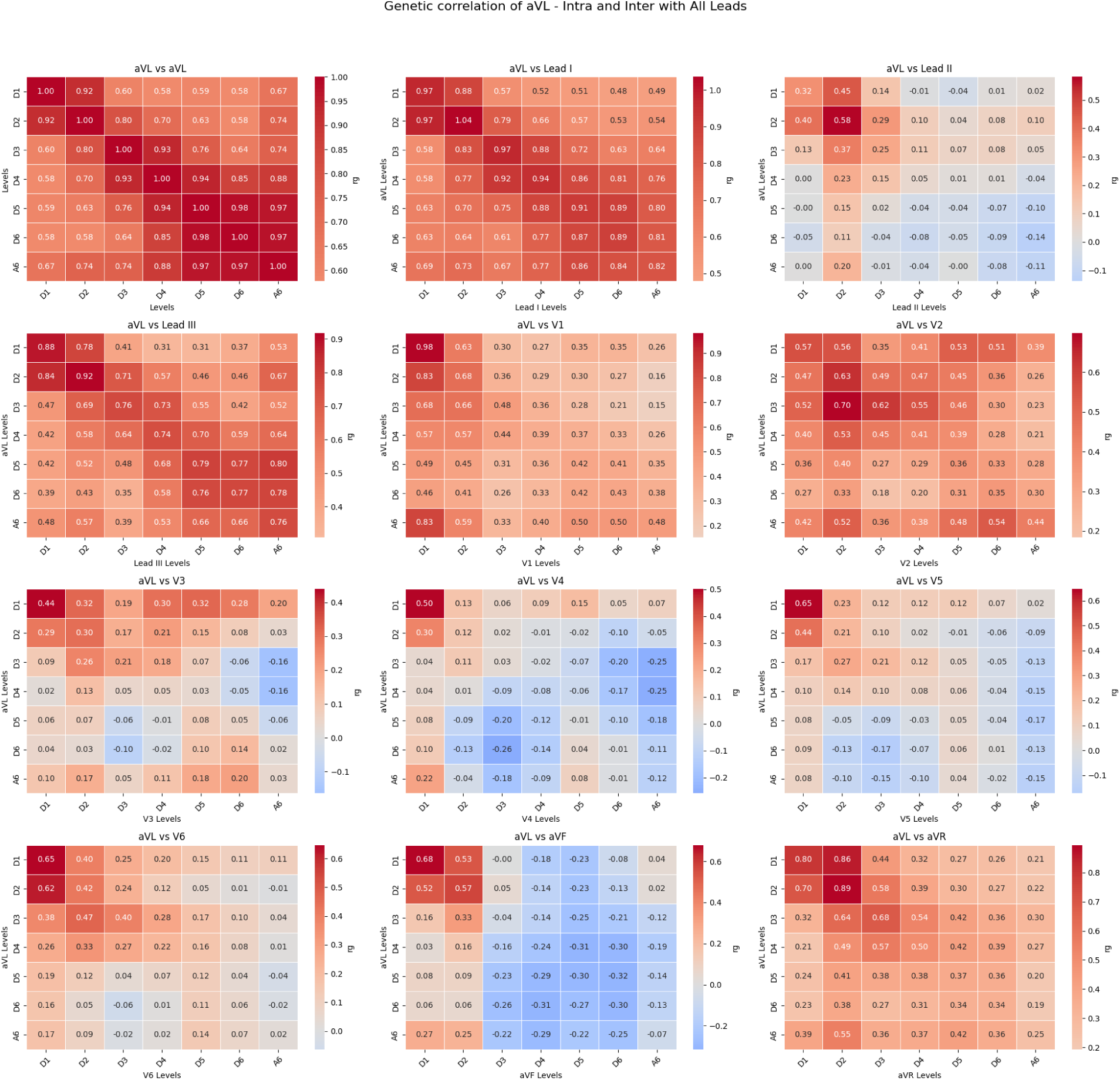
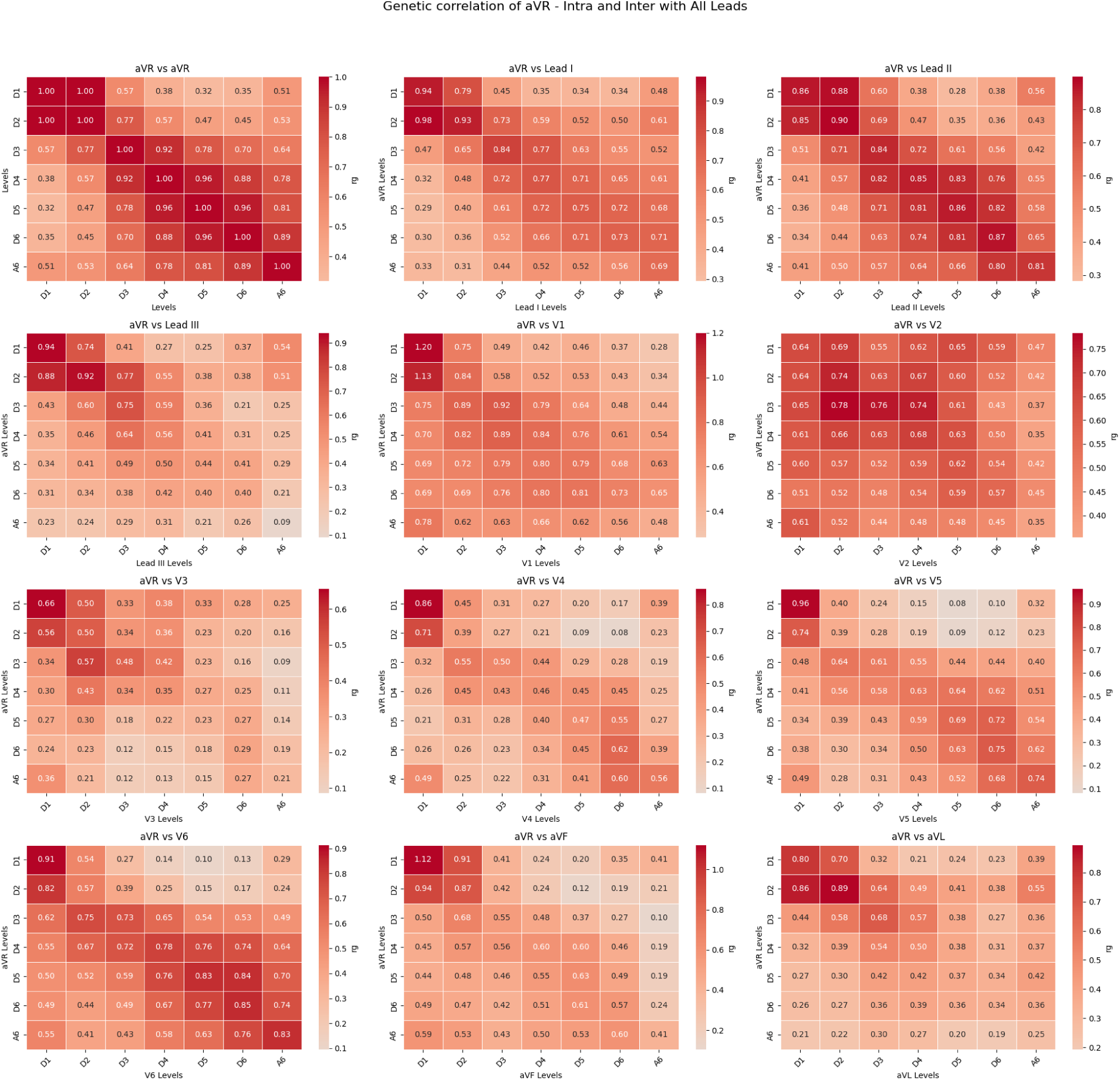
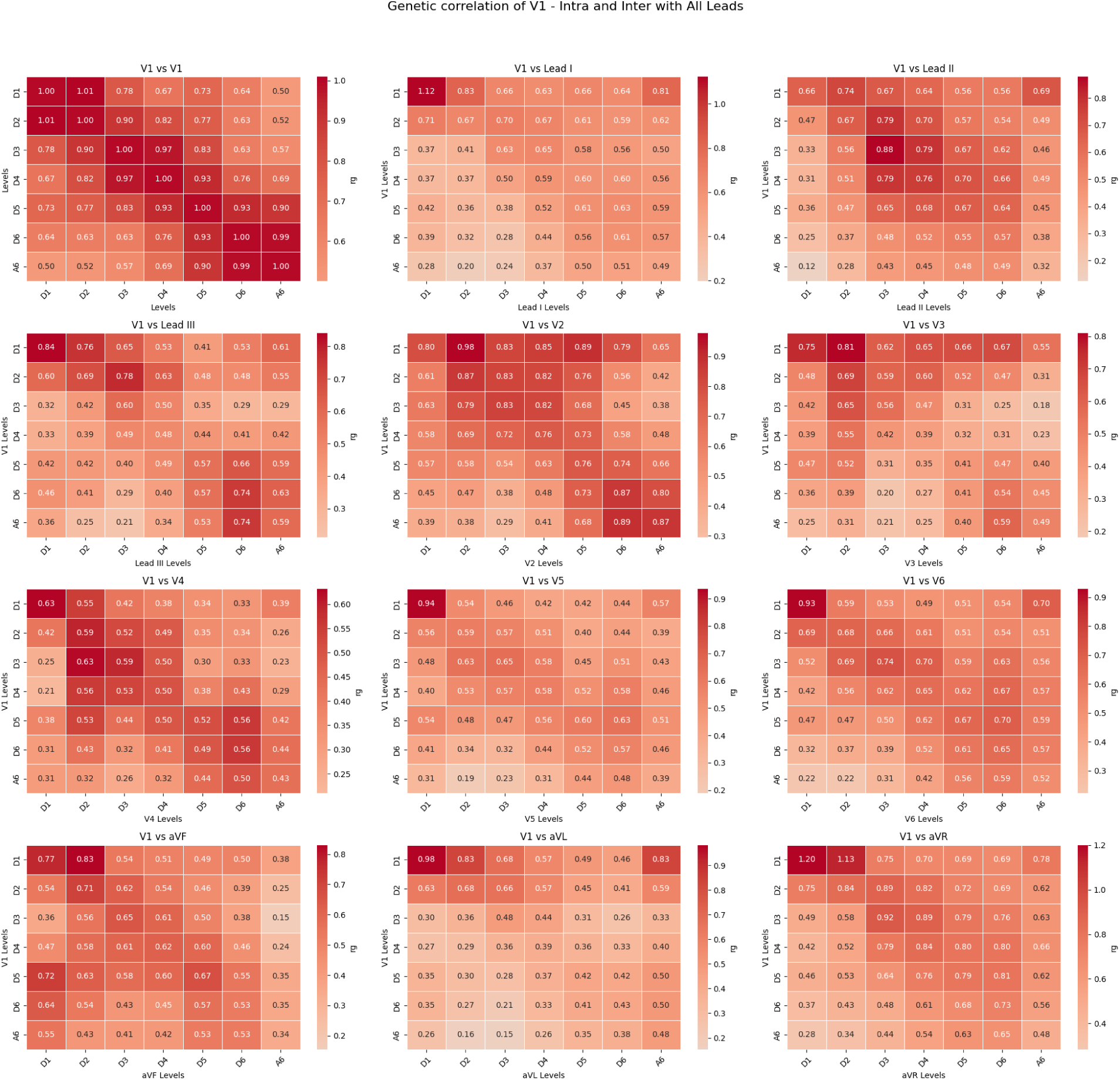
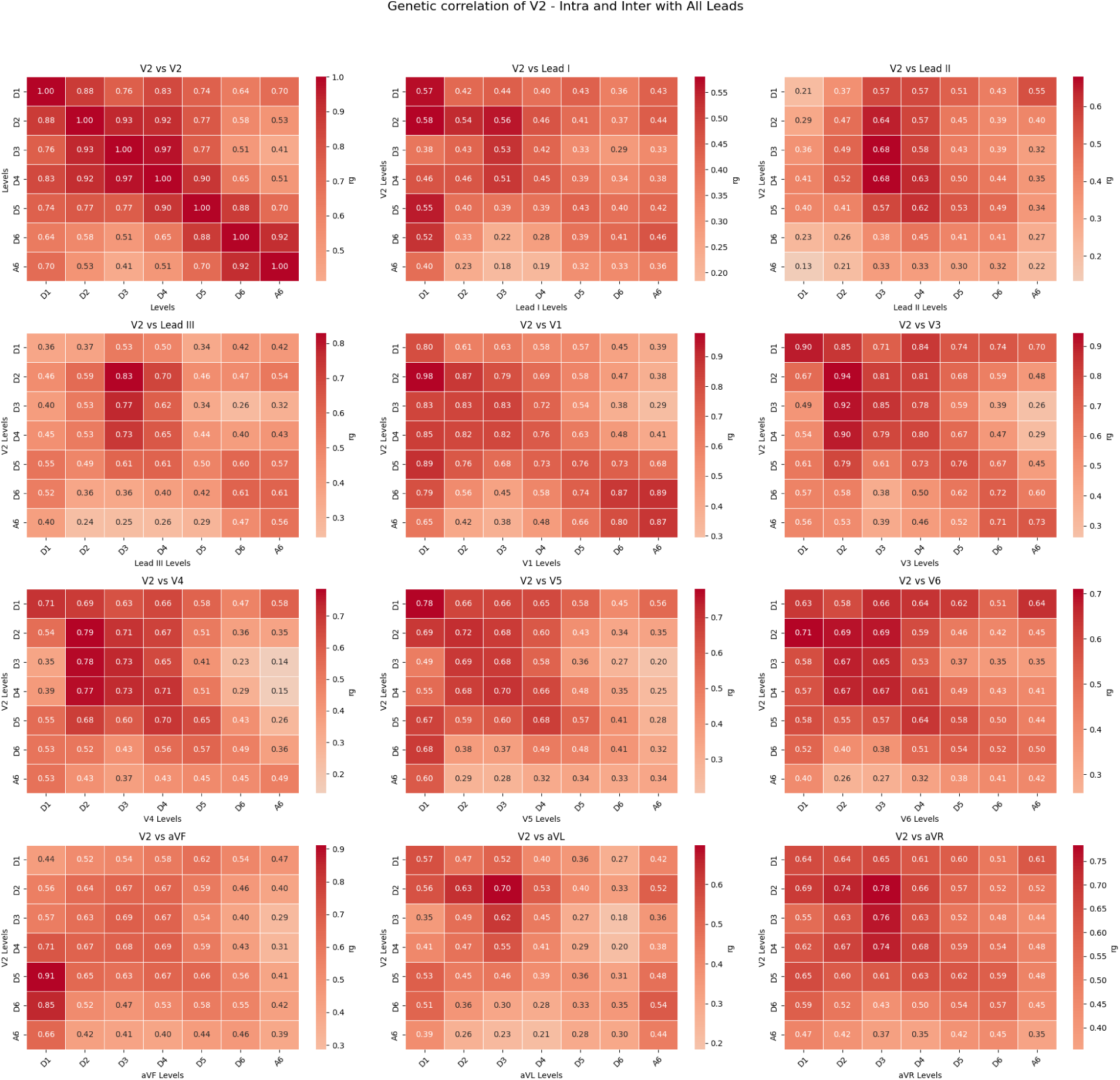
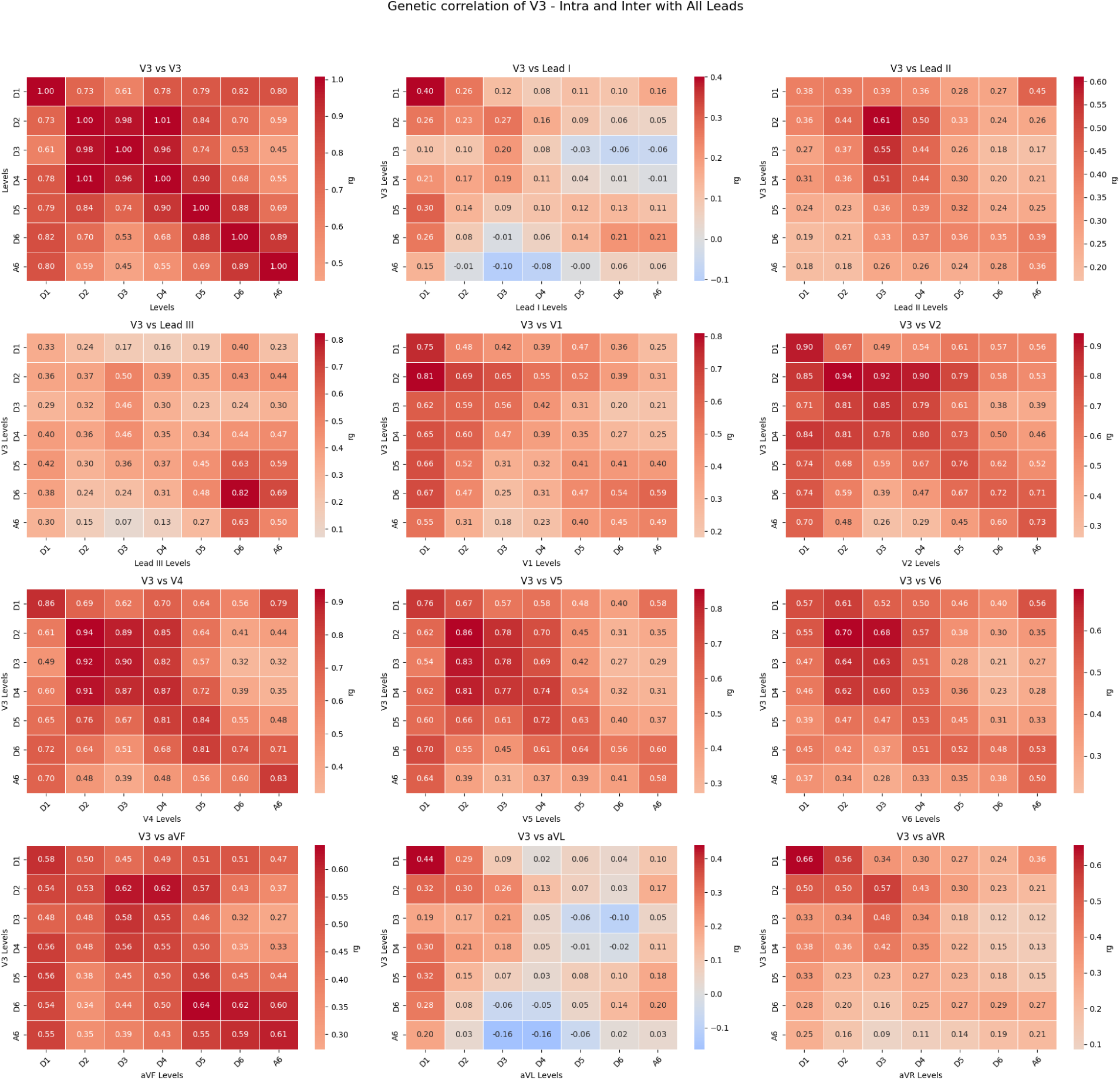
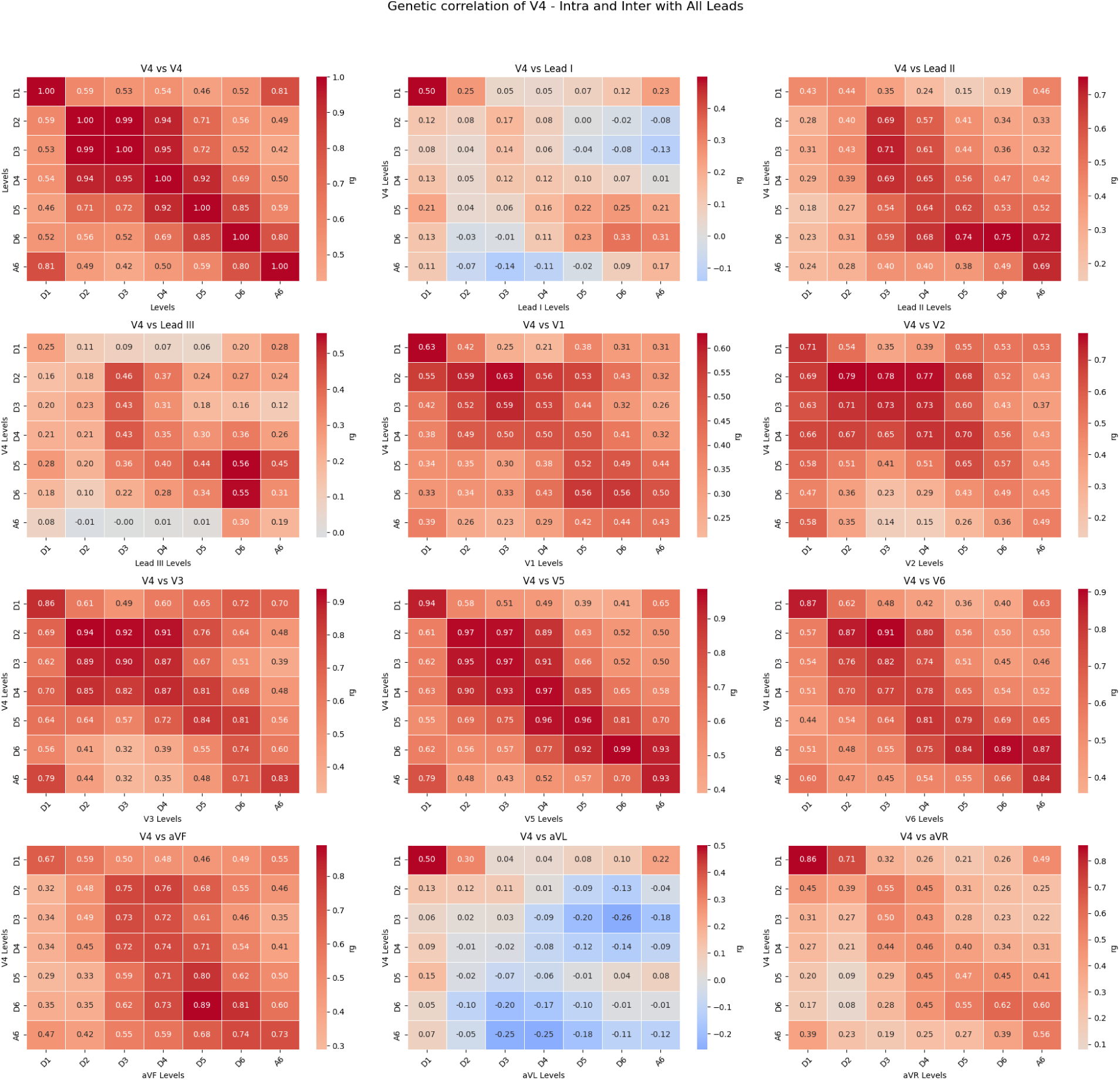
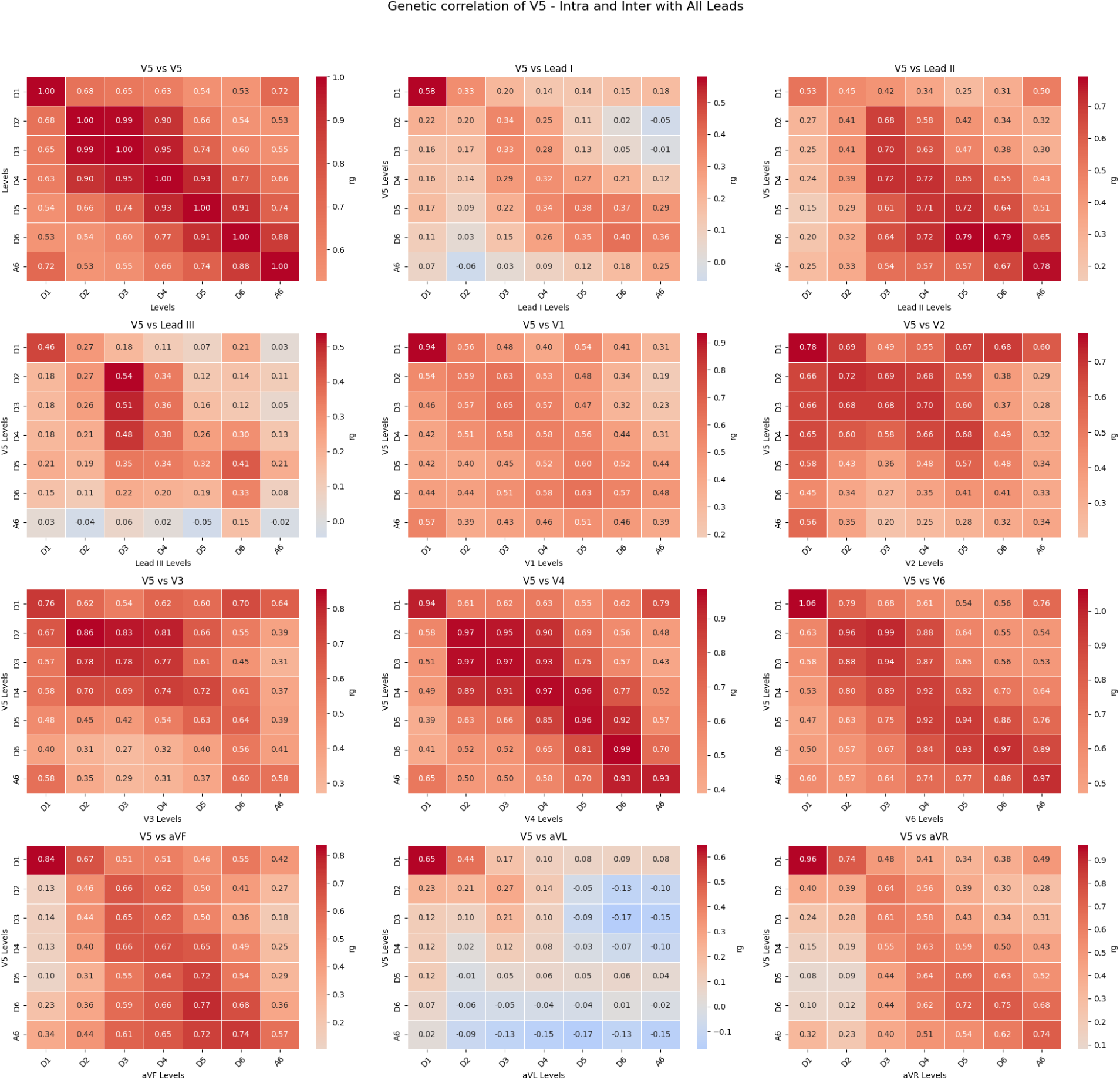
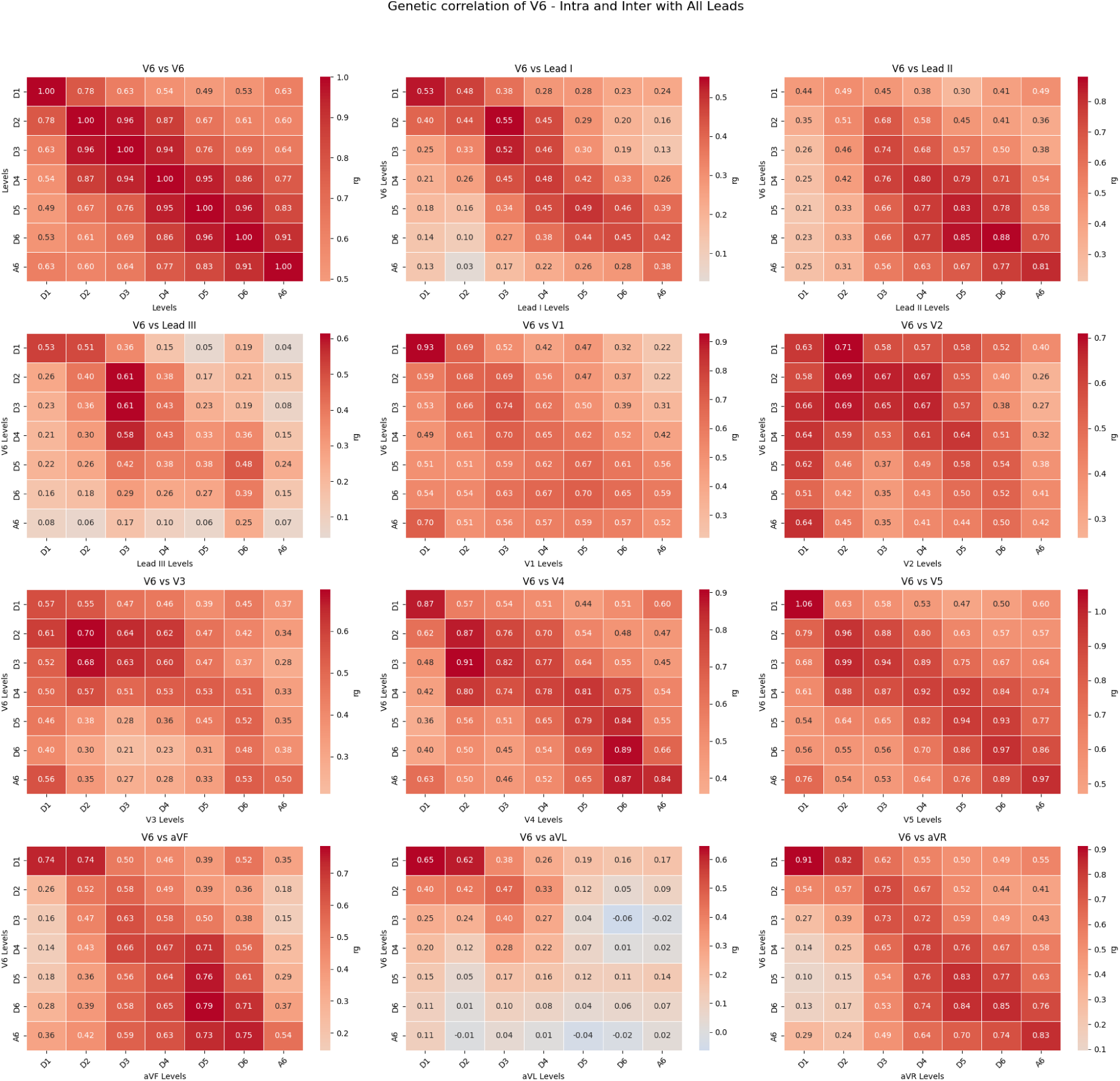
Genetic correlation estimates across leads and levels.

**Supplementary Table 1:** Significant genetic correlations between ECG energy features and cardiovascular phenotypes (p < .005 shown).

| Cardiovascular phenotypes | Energy features | rg | p |
| --- | --- | --- | --- |
| Heart failure and coronary heart disease | Lead I D1 | 0.34 | 0.0013 |
| Atherosclerosis (excluding cerebral and coronary sclerosis) | Lead I D1 | 0.3138 | 0.0016 |
| Heart failure and coronary heart disease | Lead I D2 | 0.2558 | 0.0036 |
| Atherosclerosis (excluding cerebral and coronary sclerosis) | Lead I D2 | 0.3234 | 0.0004 |
| Heart failure and coronary heart disease | Lead I D3 | 0.2065 | 0.0002 |
| Coronary heart disease wide definition | Lead I D4 | 0.1406 | 0.0034 |
| Heart failure and coronary heart disease | Lead I D4 | 0.2215 | 9.41E-05 |
| Ischaemic heart disease (wide definition) | Lead I D4 | 0.1252 | 0.0031 |
| Ischaemic heart disease (wide definition) | Lead I D5 | 0.1284 | 0.0015 |
| Coronary heart disease wide definition | Lead I D5 | 0.138 | 0.0032 |
| Coronary atherosclerosis | Lead I D5 | 0.1213 | 0.0041 |
| Heart failure and coronary heart disease | Lead I D5 | 0.2124 | 0.0001 |
| Coronary heart disease wide definition | Lead I D6 | 0.1367 | 0.0041 |
| Heart failure and coronary heart disease | Lead I D6 | 0.2185 | 0.0002 |
| Heart failure and coronary heart disease | Lead I A6 | 0.2391 | 0.0006 |
| Coronary atherosclerosis | Lead II D1 | 0.2512 | 0.004 |
| Coronary atherosclerosis | aVR D1 | 0.2952 | 0.0031 |
| Atherosclerosis (excluding cerebral and coronary sclerosis) | aVR D1 | 0.3483 | 0.0032 |
| Atherosclerosis (excluding cerebral and coronary sclerosis) | aVR D2 | 0.3682 | 0.0008 |
| Heart failure and coronary heart disease | aVR D2 | 0.3238 | 0.0044 |
| Coronary atherosclerosis | aVR D2 | 0.2469 | 0.003 |
| Coronary atherosclerosis | aVR D4 | 0.1302 | 0.0041 |
| Heart failure and coronary heart disease | aVR D4 | 0.1845 | 0.0033 |
| Heart failure and coronary heart disease | aVR D5 | 0.1819 | 0.0016 |
| Coronary atherosclerosis | aVR D5 | 0.1463 | 0.0004 |
| Ischaemic heart disease (wide definition) | aVR D5 | 0.1463 | 0.0004 |
| Coronary heart disease wide definition | aVR D5 | 0.1344 | 0.0029 |
| Heart failure and coronary heart disease | aVR D6 | 0.1854 | 0.0019 |
| Heart failure and coronary heart disease | aVL D1 | 0.3363 | 0.001 |
| Atherosclerosis (excluding cerebral and coronary sclerosis) | aVL D1 | 0.2671 | 0.0048 |
| Atherosclerosis (excluding cerebral and coronary sclerosis) | aVL D2 | 0.3062 | 0.0012 |
| Heart failure and coronary heart disease | aVL D3 | 0.2051 | 0.0028 |
| Heart failure and coronary heart disease | aVL D4 | 0.2343 | 0.0019 |
| Heart failure and coronary heart disease | aVL D5 | 0.24 | 0.0027 |
| Left bundle branch block | aVL A6 | 0.4233 | 0.0026 |
| Heart failure and coronary heart disease | aVL D6 | 0.2377 | 0.0039 |
| Left bundle branch block | aVL D6 | 0.3744 | 0.0021 |
| Heart failure and coronary heart disease | aVL A6 | 0.2954 | 0.0049 |
| Heart failure and coronary heart disease | V1 D1 | 0.5591 | 0.0039 |
| Coronary atherosclerosis | V1 D1 | 0.3894 | 0.0027 |
| Ischaemic heart disease (wide definition) | V1 D1 | 0.4187 | 0.003 |
| Ischaemic heart disease (wide definition) | V1 D2 | 0.2082 | 0.0011 |
| Coronary heart disease wide definition | V1 D2 | 0.2245 | 0.0004 |
| Coronary atherosclerosis | V1 D2 | 0.2292 | 0.0003 |
| Heart failure and coronary heart disease | V1 D2 | 0.3353 | 0.0004 |
| Heart failure and coronary heart disease | V1 D3 | 0.225 | 0.0013 |
| Coronary atherosclerosis | V1 D4 | 0.1479 | 0.0033 |
| Heart failure and coronary heart disease | V1 D4 | 0.2735 | 5.04E-05 |
| Coronary heart disease wide definition | V1 D4 | 0.1617 | 0.0026 |
| Coronary atherosclerosis | V1 D5 | 0.2178 | 0.0001 |
| Heart failure and coronary heart disease | V1 D5 | 0.3322 | 3.79E-05 |
| Ischaemic heart disease (wide definition) | V1 D5 | 0.2102 | 0.0002 |
| Coronary heart disease wide definition | V1 D5 | 0.2237 | 0.0002 |
| Coronary heart disease wide definition | V1 D6 | 0.2253 | 0.0003 |
| Coronary heart disease wide definition | V1 A6 | 0.3073 | 4.41E-05 |
| Heart failure and coronary heart disease | V1 A6 | 0.376 | 0.0001 |
| Heart failure and coronary heart disease | V1 D6 | 0.3164 | 0.0002 |
| Ischaemic heart disease (wide definition) | V1 A6 | 0.2731 | 0.0001 |
| Ischaemic heart disease (wide definition) | V1 D6 | 0.1969 | 0.0007 |
| Coronary atherosclerosis | V1 A6 | 0.2967 | 3.97E-05 |
| Coronary atherosclerosis | V1 D6 | 0.2097 | 0.0004 |
| Heart failure and coronary heart disease | V2 D2 | 0.2784 | 0.0013 |
| Coronary heart disease wide definition | V2 D2 | 0.2311 | 0.0006 |
| Heart failure and coronary heart disease | V2 D3 | 0.2253 | 0.0037 |
| Coronary heart disease wide definition | V2 D3 | 0.2061 | 0.0012 |
| Ischaemic heart disease (wide definition) | V2 D4 | 0.1922 | 0.004 |
| Heart failure and coronary heart disease | V2 D4 | 0.2479 | 0.0049 |
| Coronary heart disease wide definition | V2 D4 | 0.2265 | 0.001 |
| Ischaemic heart disease (wide definition) | V2 D5 | 0.2541 | 0.0012 |
| Coronary heart disease wide definition | V2 D5 | 0.2559 | 0.0019 |
| Coronary atherosclerosis | V2 D5 | 0.2311 | 0.0031 |
| Ischaemic heart disease (wide definition) | V2 A6 | 0.2676 | 0.0012 |
| Heart failure and coronary heart disease | V2 A6 | 0.2867 | 0.0045 |
| Coronary heart disease wide definition | V2 D6 | 0.276 | 0.0021 |
| Coronary atherosclerosis | V2 D6 | 0.2795 | 0.0015 |
| Coronary heart disease wide definition | V2 A6 | 0.264 | 0.0017 |
| Coronary atherosclerosis | V2 A6 | 0.259 | 0.0015 |
| Ischaemic heart disease (wide definition) | V2 D6 | 0.2883 | 0.001 |
| Valvular heart disease | V5 D6 | 0.2248 | 0.0016 |
| Coronary heart disease wide definition | V6 D1 | 0.3458 | 0.0017 |
| Ischaemic heart disease (wide definition) | V6 D1 | 0.3709 | 0.0004 |
| Coronary atherosclerosis | V6 D1 | 0.3778 | 0.0004 |
| Heart failure and coronary heart disease | V6 D1 | 0.419 | 0.0021 |
| Heart failure and coronary heart disease | V6 D2 | 0.2372 | 0.0025 |
| Coronary atherosclerosis | V6 D2 | 0.1971 | 0.0017 |
| Ischaemic heart disease (wide definition) | V6 D2 | 0.1964 | 0.0016 |

**Supplementary Table 2:**
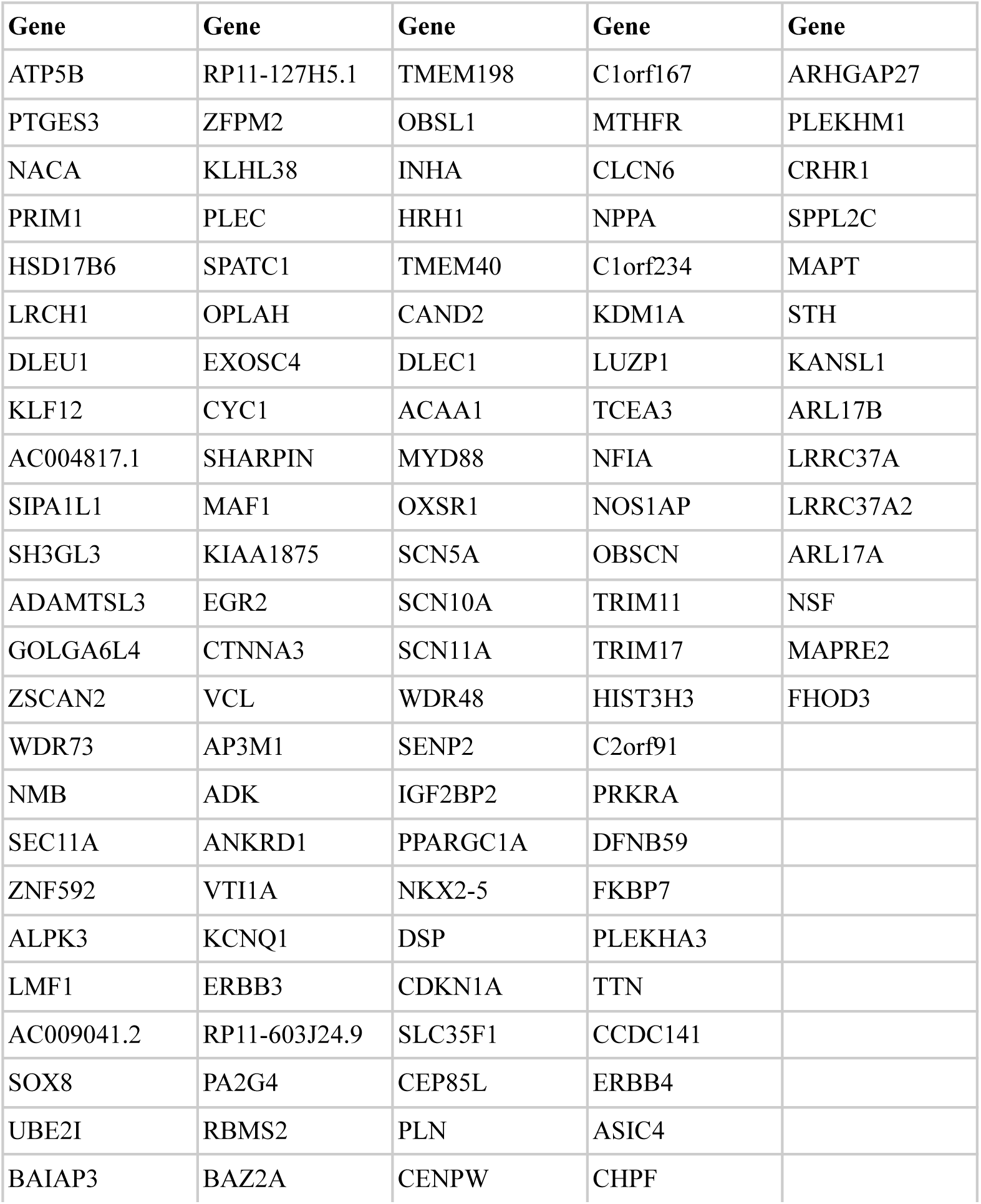
List of 110 genes mapped from clumped leading SNPs via the FUMA snps2genes tool.

**Supplementary Table 3:**
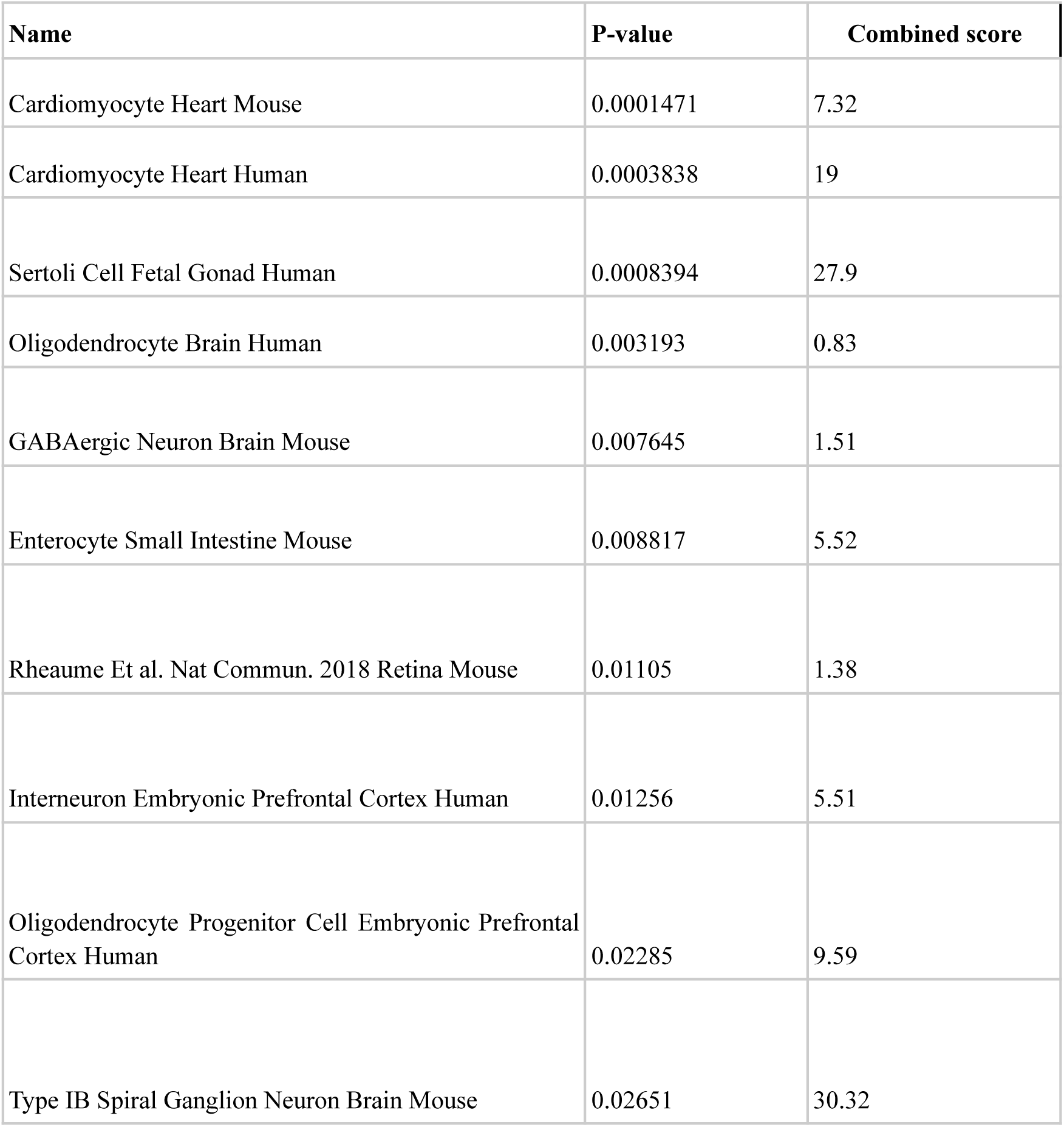
Cell type enrichment analysis results from Enrichr.

**Supplementary Table 4:** Sensitivity analysis for reverse causation in D1 genetic correlations.

| Phenotype | Lead I | Lead II | Lead III | aVR | aVL | aVF | V1 | V2 | V3 | V4 | V5 | V6 |
| --- | --- | --- | --- | --- | --- | --- | --- | --- | --- | --- | --- | --- |
| Heart failure & CHD | 0.32*<br>* | 0.21 | 0.12 | 0.30* | 0.24* | 0.21 | 0.38*<br>* | 0.26* | 0.06 | 0.14 | 0.60 | 0.52 |
| Coronary atherosclerosis | 0.27*<br>* | 0.29*<br>* | 0.03 | 0.32*<br>* | 0.14 | 0.17 | 0.31*<br>* | 0.14 | 0.07 | 0.24* | 0.68 | 0.57* |
| Coronary heart disease | 0.24*<br>* | 0.26*<br>* | 0.05 | 0.26*<br>* | 0.15* | 0.21 | 0.26*<br>* | 0.12 | 0.08 | 0.23* | 0.57 | 0.46* |
| Ischaemic heart disease | 0.24*<br>* | 0.23* | -0.01 | 0.29*<br>* | 0.12 | 0.14 | 0.31*<br>* | 0.17* | 0.07 | 0.21 | 0.66 | 0.56* |
| Atherosclerosis | 0.30*<br>* | 0.14 | 0.11 | 0.27* | 0.22* | 0.07 | 0.13 | 0.12 | -0.02 | 0.07 | 0.36 | 0.40 |
| Atrial fibrillation | 0.13 | 0.07 | -0.02 | 0.10 | 0.06 | -0.01 | 0.10 | 0.02 | -0.03 | 0.01 | 0.41 | 0.32 |
| Cardiomyopathy | 0.14 | -0.02 | 0.17 | 0.06 | 0.13 | 0.10 | 0.10 | 0.14 | 0.03 | 0.05 | 0.36 | 0.27 |
| Conduction disorder | 0.16 | -0.26 | 0.00 | -0.06 | 0.10 | -0.36*<br>* | 0.00 | -0.07 | -0.02 | -0.13 | 0.08 | -0.16 |
| AV block | 0.15 | -0.13 | 0.03 | 0.04 | 0.09 | -0.22 | -0.02 | -0.07 | 0.00 | -0.06 | 0.29 | 0.02 |
| Left bundle branch block | -0.02 | -0.31 | -0.14 | -0.26 | 0.01 | -0.29 | -0.15 | 0.05 | -0.17 | -0.27 | -0.31 | -0.29 |
| Paroxysmal tachycardia | 0.13 | 0.18 | 0.01 | 0.19 | 0.00 | 0.16 | 0.04 | 0.07 | 0.16 | 0.10 | 0.53 | 0.58 |
| Valvular heart disease | 0.06 | -0.08 | -0.09 | -0.04 | -0.04 | -0.11 | -0.12 | 0.07 | 0.19 | 0.11 | 0.28 | 0.13 |

\* p<0.05 \*\* p<0.005 \*\*\* p<0.0005

### Supplementary Analysis 1: Predictive Modeling With Wavelet-Derived ECG Features

We tested if wavelet energy features capture heart-specific electrical patterns by training LASSO logistic regression models for three UK Biobank outcomes: atrial fibrillation, acute myocardial infarction and myocardial infarction defined by diagnostic codes. We included physician-diagnosed asthma as a negative-control phenotype. Models used all 84 wavelet-derived energy features together with age, sex and four genetic principal components as covariates. To provide an orthogonal view of feature–phenotype relationships, we also performed univariate association testing on covariate-residualized energies (Energy ∼ age + sex +4PCs), enabling direct visualization of lead- and band-specific patterns.

For atrial fibrillation (AUC = 0.75), predictive signal concentrated in mid-to-high frequency detail bands (D5–D1), most prominently in V1 and adjacent precordial leads, consistent with the loss of organized atrial activity and increased fragmentation of atrial electrical dynamics; conversely, low-frequency components in Lead II and aVR carried negative weights, aligning with preserved, structured atrial activity under sinus rhythm.

For myocardial infarction (AUC = 0.78), selected coefficients were enriched in mid-frequency bands (D6–D4) across anterior–lateral precordial leads (V3–V6), implicating altered QRS morphology and early repolarization dynamics in the predictive signal, with smaller contributions from intermediate bands (D3–D2) suggestive of localized changes in waveform complexity. Notably, multivariable selections were concordant with the univariate lead–band association maps, indicating that sparse models recover coherent, physiologically interpretable patterns rather than diffuse correlates.

In contrast, the asthma negative control showed near-null discrimination (AUC = 0.564) with low-magnitude, spatially unstructured coefficients and weak univariate signals, supporting the specificity of the cardiovascular patterns to electrophysiological remodeling rather than generic confounding.

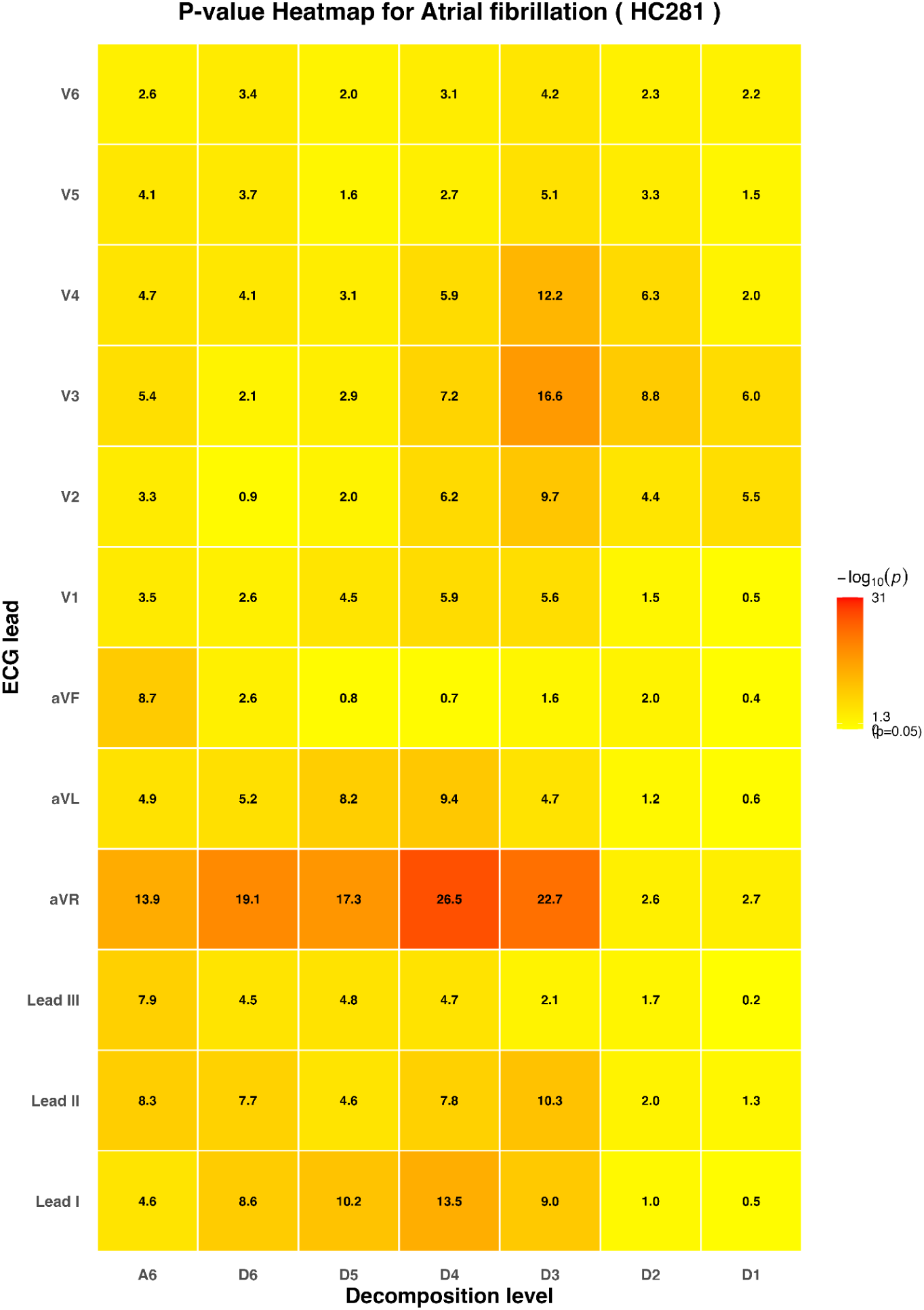
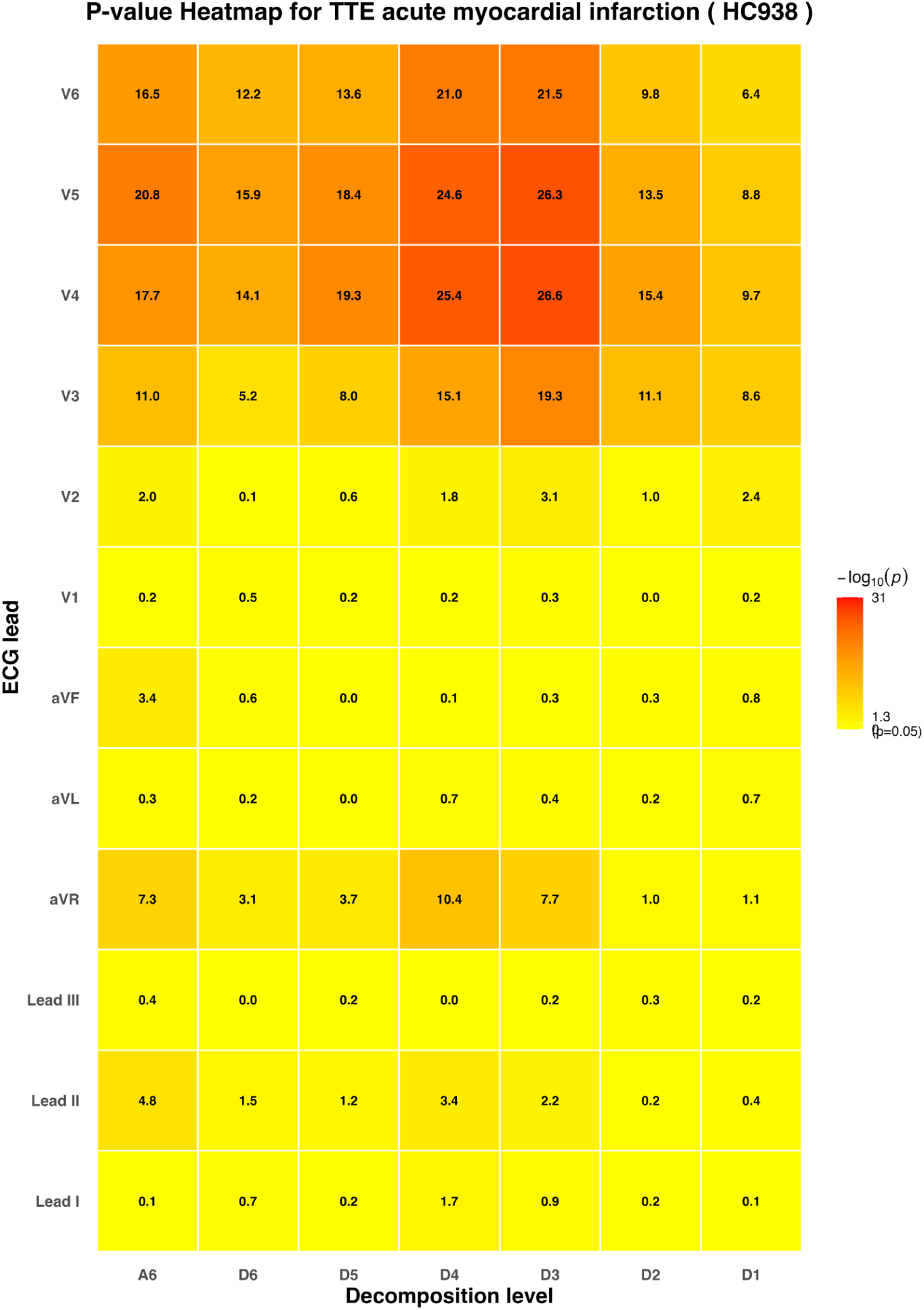
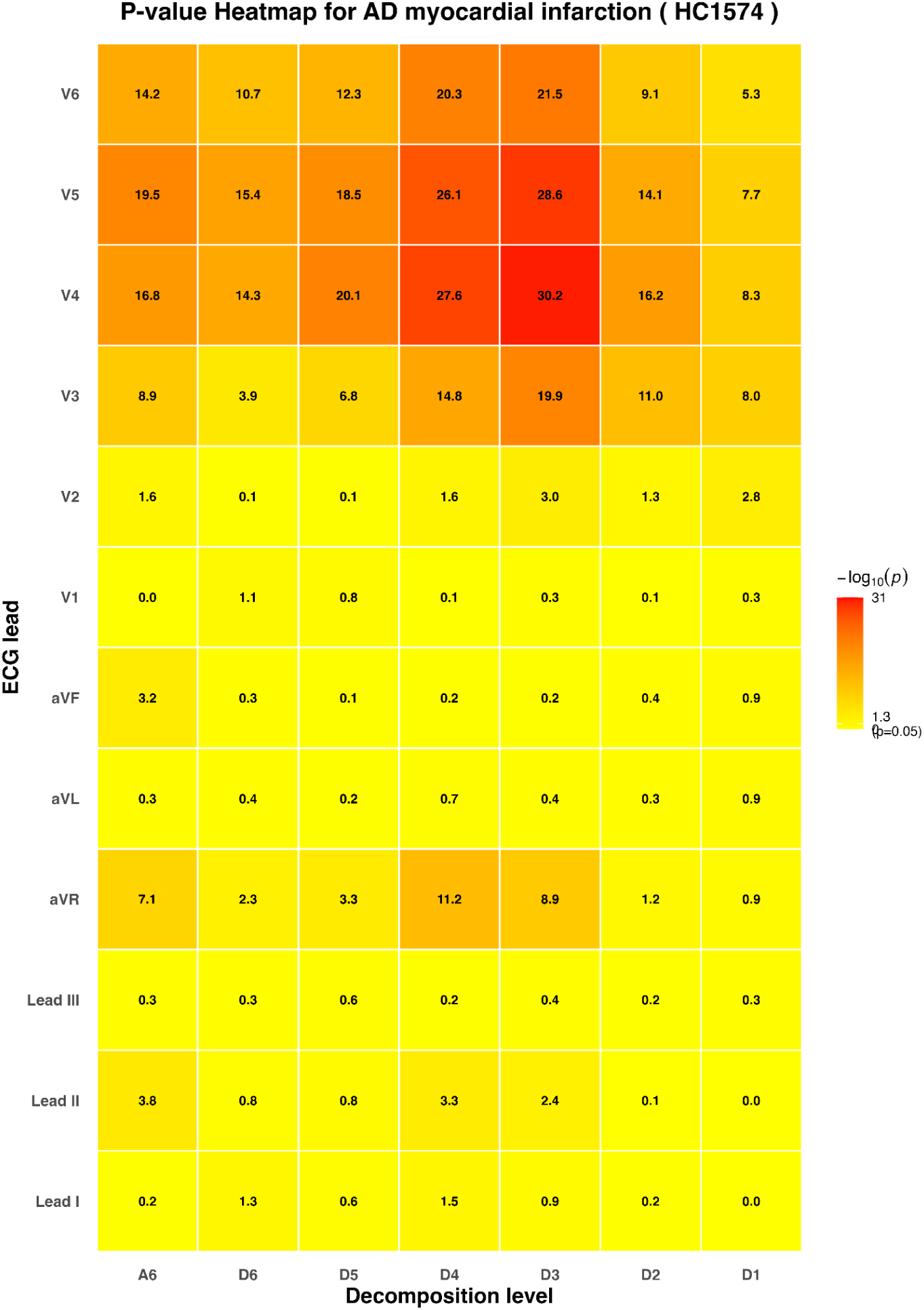
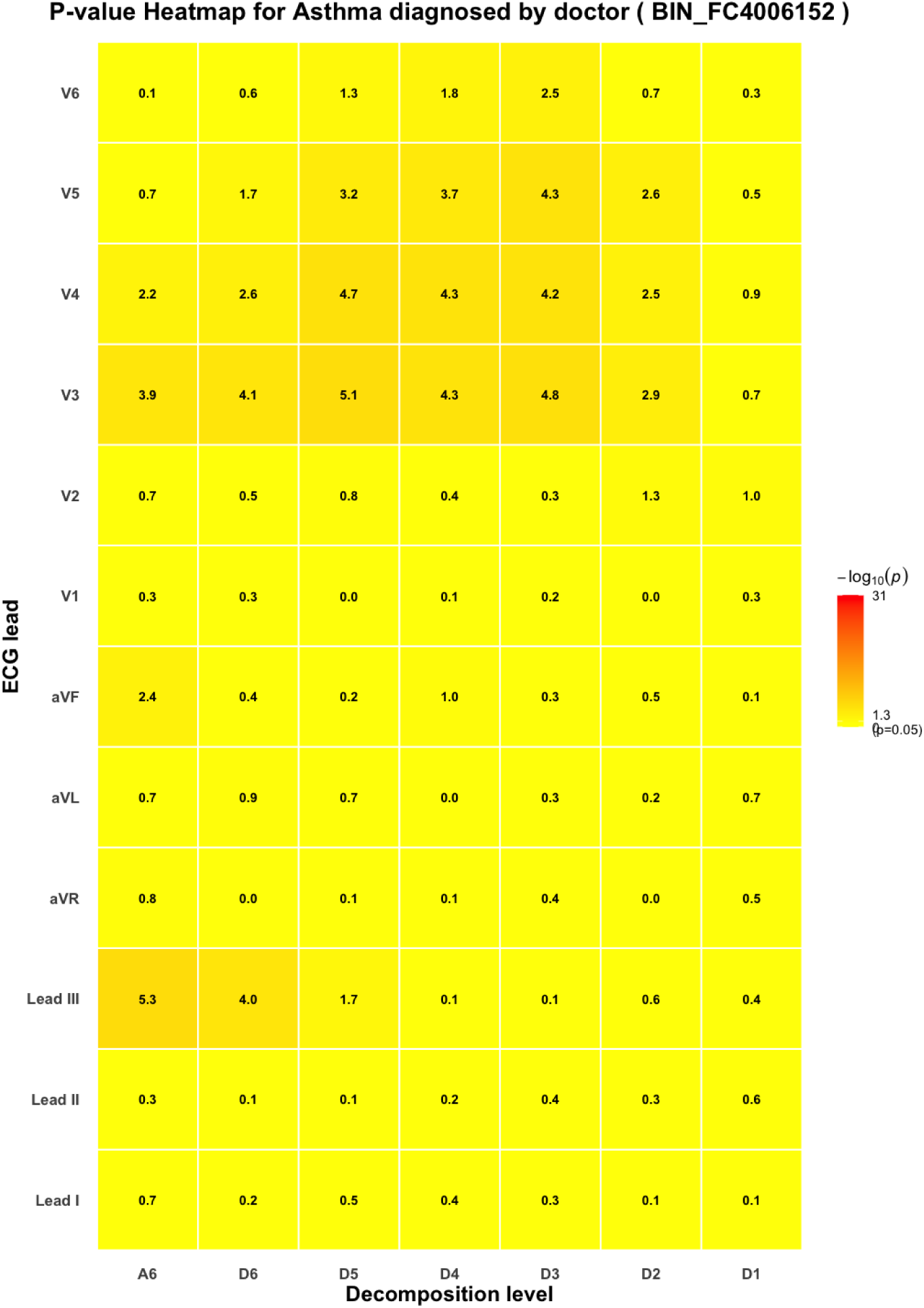
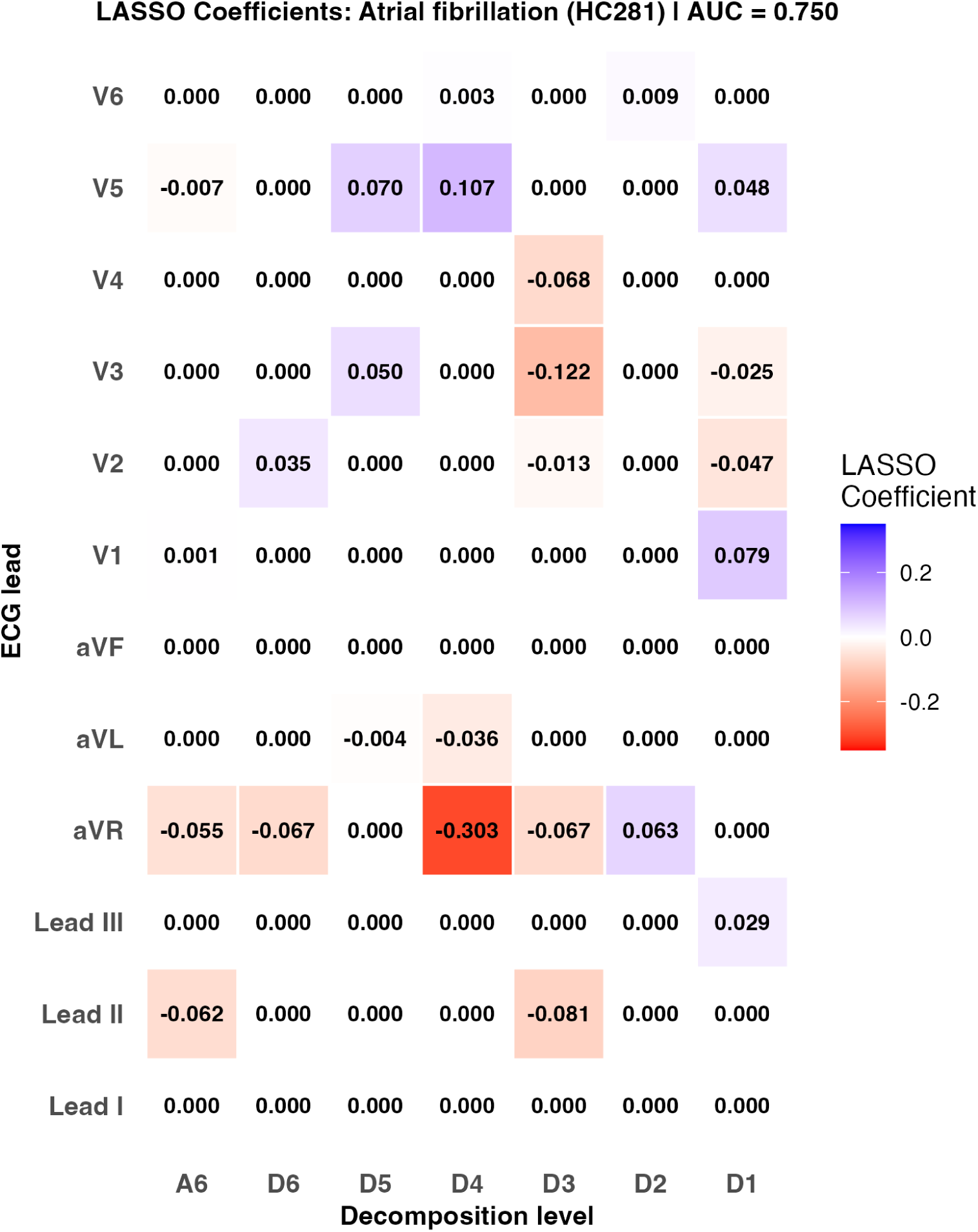
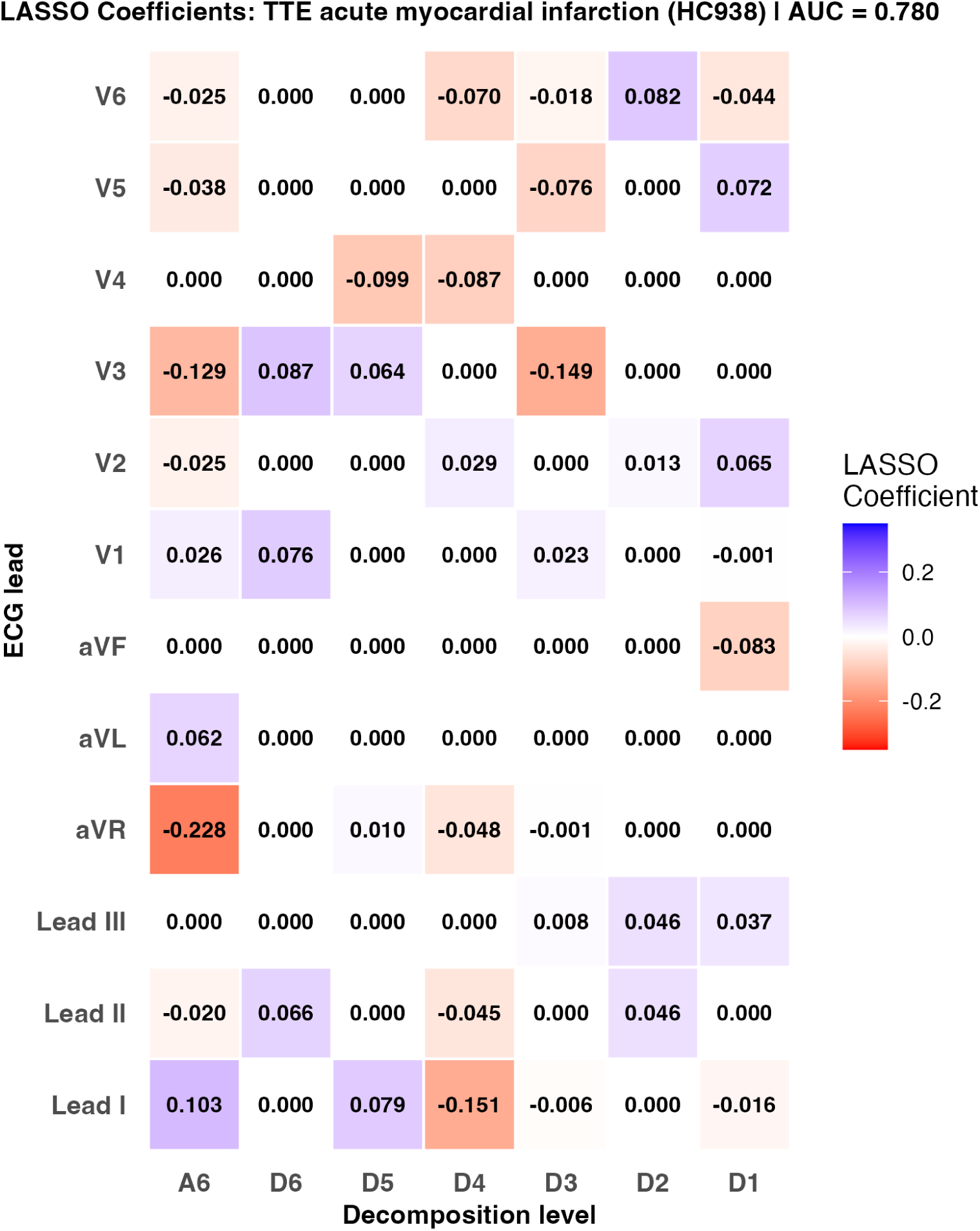
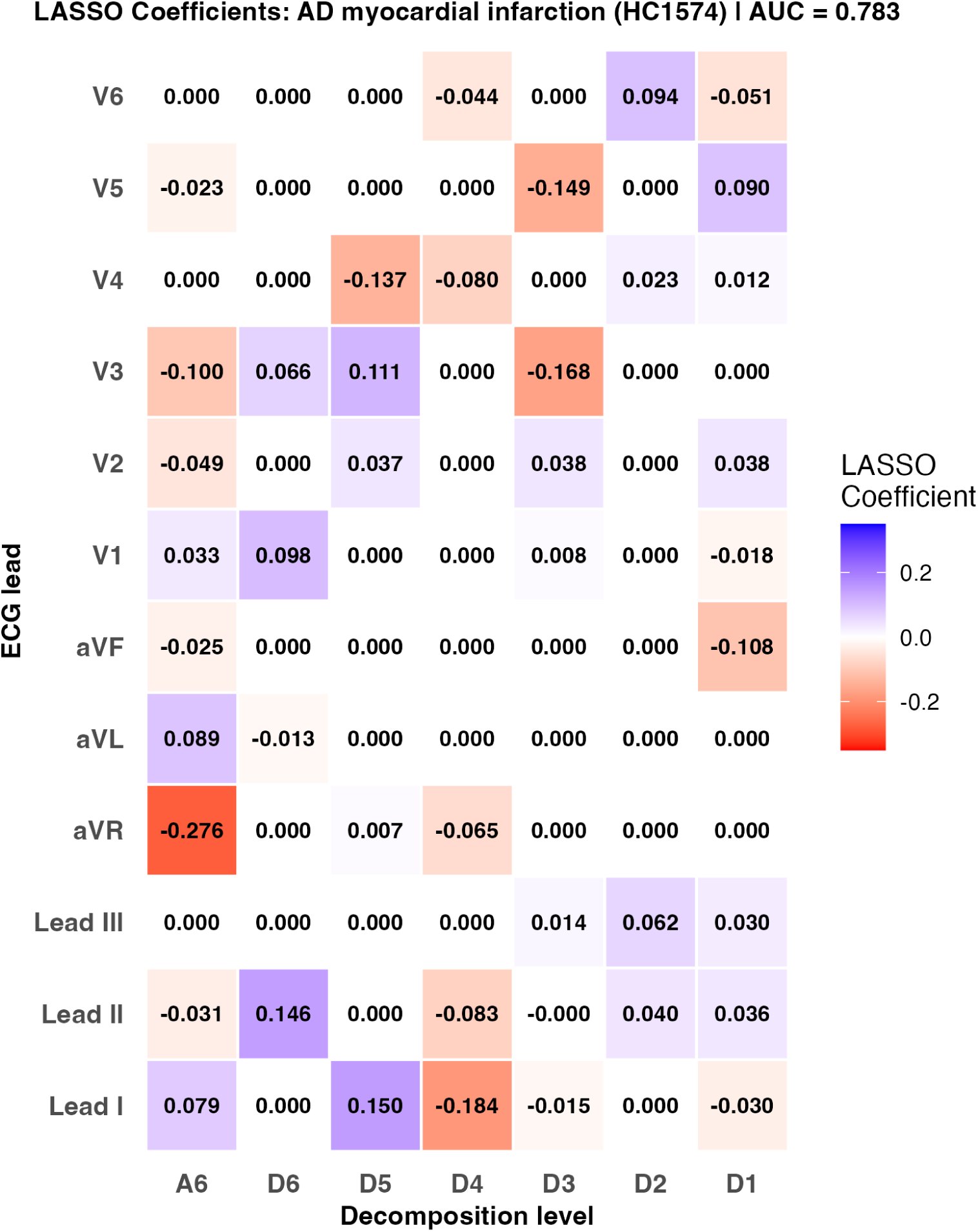
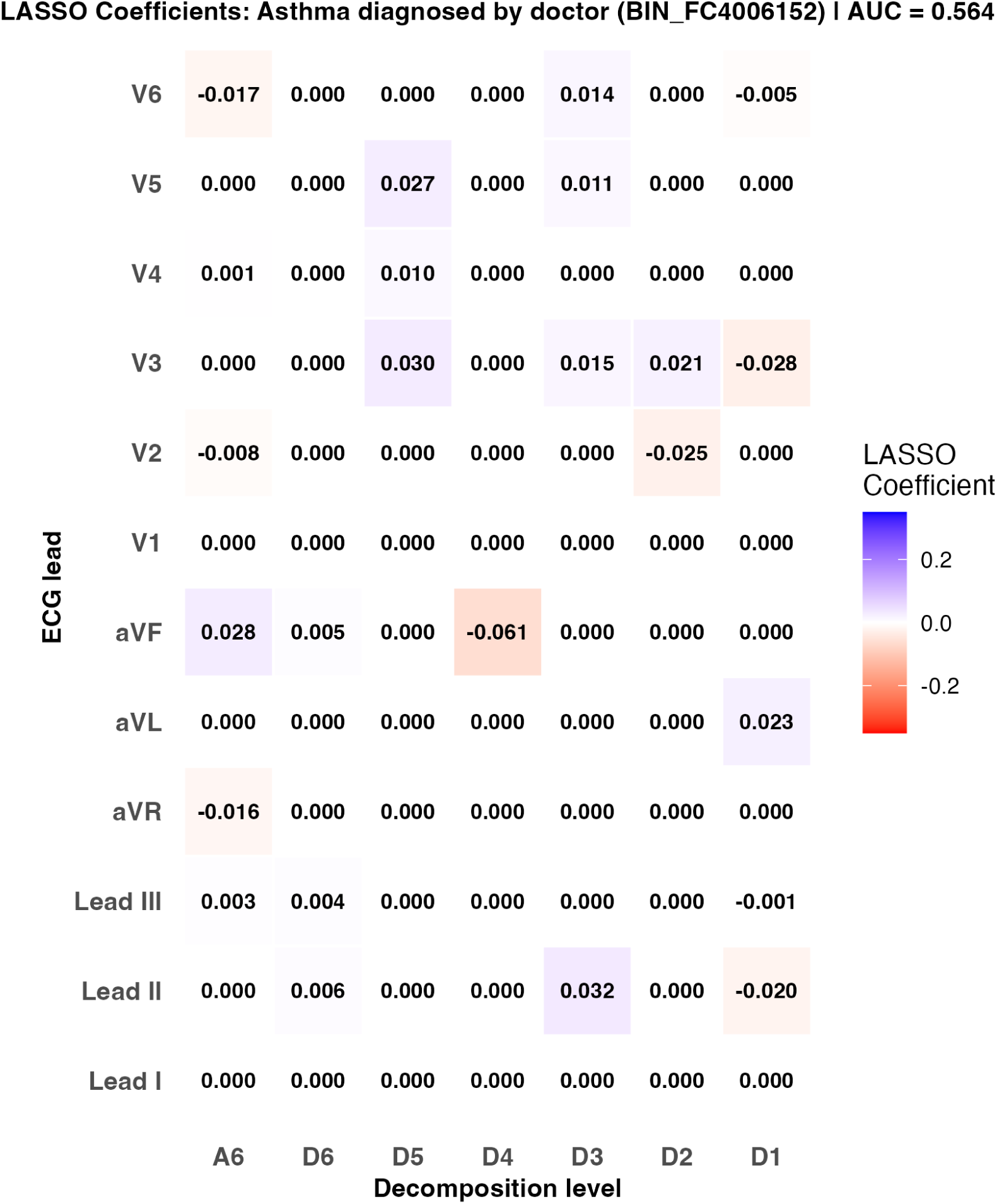

### Supplementary Analysis 2: Independence of D1 cardiac signal from BMI genetics

To determine whether the genetic associations identified in the D1 frequency band (125–250 Hz) reflect cardiac biology rather than adiposity-related artifact, we conducted three complementary analyses.

#### Partial genetic correlation

We computed the partial genetic correlation between D1 energy and heart failure (HF) conditioning on BMI for all 12 leads, using the trivariate formula

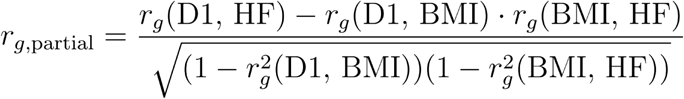

where rg(BMI, HF) = 0.354 (SE = 0.041, p = 1.2×10^−17^, estimated by LDSC against FinnGen R12). The partial rg remained substantial across all leads with significant D1–HF associations. For V1 — the lead with the strongest association — the partial rg was 0.548 vs total rg = 0.559, a reduction of 2%. For leads V5 and V6, the partial rg exceeded the total rg (V5: 0.487→0.547; V6: 0.419→0.463), indicating that BMI acts as a suppressor for precordial leads rather than a confounder. Results for all 12 leads are shown in **Figure SA2a**.

#### BMI-conditioned GWAS

We repeated all 12 D1 GWAS with BMI as an additional covariate (Model B: energy_D1 ∼ SNP + age + sex + PC1–10 + BMI) and compared effect sizes at the top loci against the primary GWAS (Model A). The lead SNP in the SCN5A region (Affx-89018181, chr3p22.2), encoding the cardiac sodium channel Nav1.5, showed beta attenuation of −1.9% to +1.6% across all 12 leads — negligible across the board. The only genome-wide significant TTN variant (rs34940894, V2 D1, p = 4.6×10^−9^) showed attenuation of −2.7%, with the effect slightly strengthened after conditioning, consistent with BMI acting as noise at this locus. All points fall within the ±10% band **(Figure SA2b)**.

#### Genetic correlations after BMI conditioning

LDSC genetic correlations between BMI-adjusted D1 summary statistics and FinnGen R12 HF were re-estimated for all 12 leads. V1 retained a significant association (rg = 0.363, SE = 0.128, p = 0.0045). Lead I (rg = 0.237, p = 0.033) and V6 (rg = 0.541, p = 0.037) also showed nominal significance. Results are shown in **Figure SA2a** .

Together these analyses demonstrate that D1 energy associations with cardiovascular disease are not explained by shared adiposity genetics, and that the top D1 GWAS loci encode cardiac rather than metabolic biology.

**Figure SA2a.**
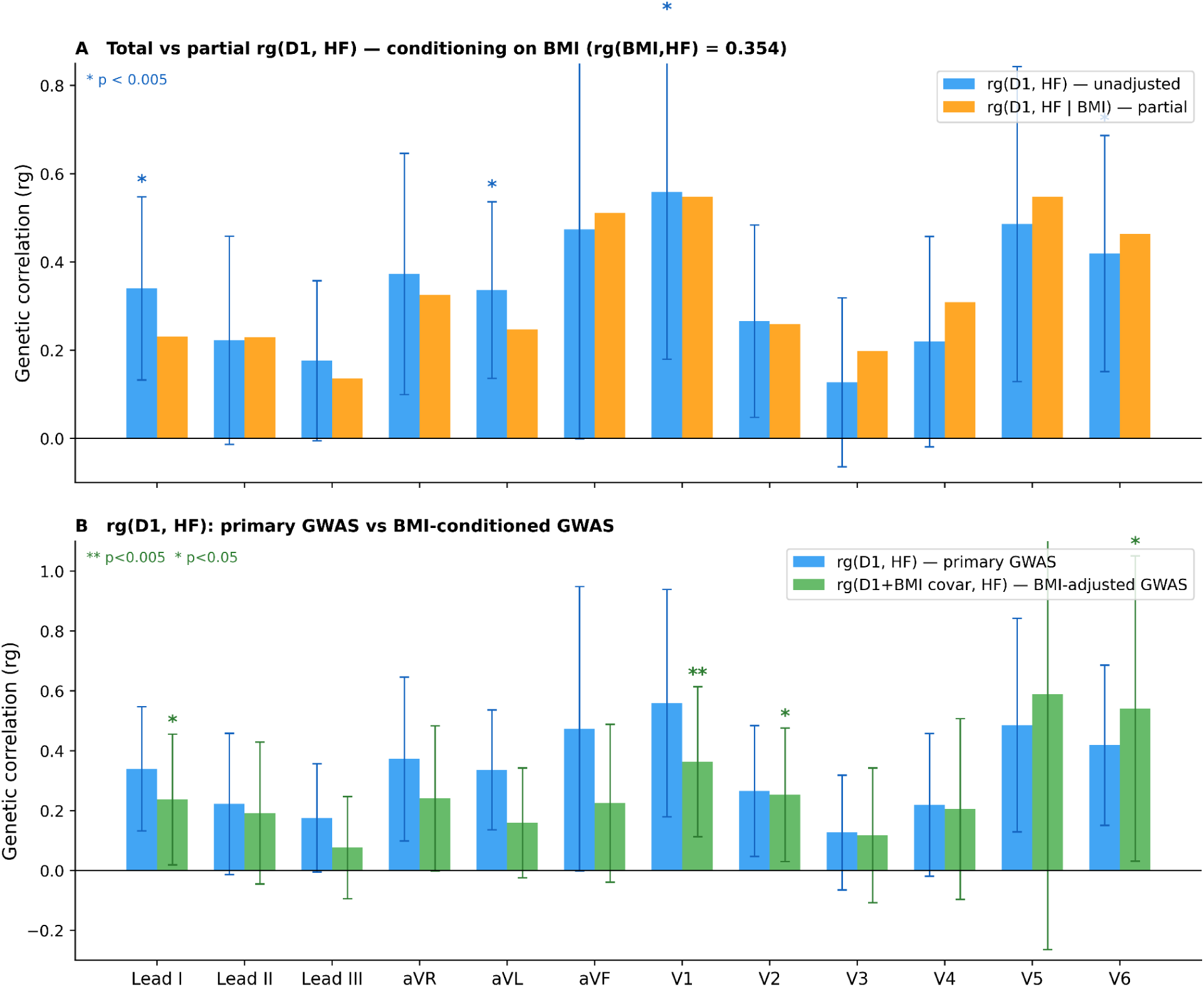
Two-panel bar plot comparing genetic correlations between D1 energy features and heart failure across 12 ECG leads. Each bar group shows three estimates: the primary genetic correlation rg(D1, HF) estimated from the original GWAS (blue), the partial genetic correlation rg(D1, HF | BMI) conditioning on BMI using the trivariate LDSC formula with rg(BMI, HF) = 0.354 (orange), and the genetic correlation estimated from BMI-conditioned GWAS summary statistics rg(D1+BMI, HF) (green). Error bars represent 95% confidence intervals (1.96 × SE). Significance markers indicate p < 0.05 (*) and p < 0.005 (**). Panel A shows the total vs partial rg, demonstrating that conditioning on BMI genetics produces negligible attenuation for most leads, with V1 retaining the strongest association (rg partial = 0.548 vs rg total = 0.559). Panel B shows the primary vs BMI-conditioned GWAS rg, with V1 remaining the only lead significant at p < 0.005 after BMI conditioning (rg = 0.363, p = 0.0045). Together, these panels demonstrate that the D1–heart failure genetic overlap is not mediated through shared adiposity genetics.

**Figure SA2b.**
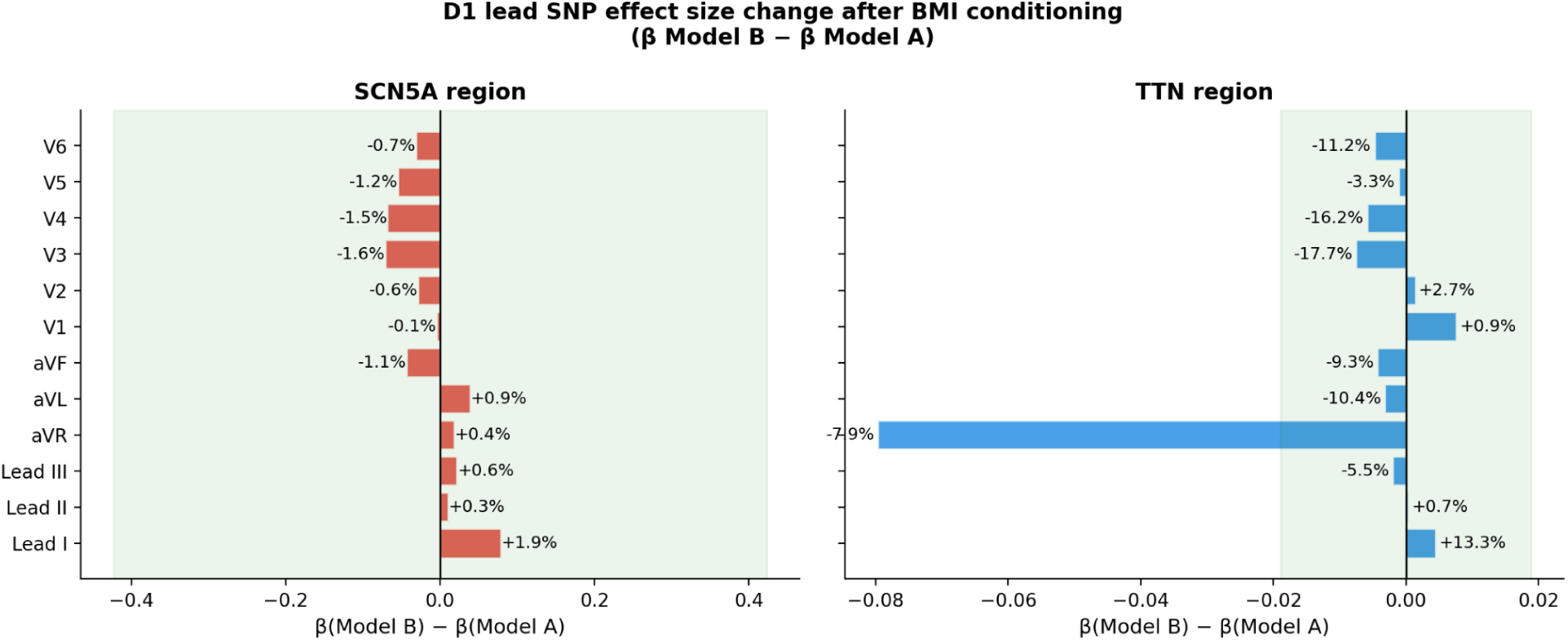
Horizontal bar plots showing the change in lead SNP effect size (Δβ = β Model B − β Model A) after adding BMI as a covariate, at the SCN5A region (chr3p22.2, left panel) and TTN region (chr2q31.2, right panel), across all 12 D1 leads. Model A: energy_D1 ∼ SNP + age + sex + PC1–10; Model B: energy_D1 ∼ SNP + age + sex + PC1–10 + BMI. Each bar represents one ECG lead. The green band indicates the ±10% attenuation zone. For SCN5A (Affx-89018181), all 12 leads show Δβ within ±2% (median attenuation 0.3%), confirming that the cardiac sodium channel locus is unaffected by BMI conditioning. For TTN, the median attenuation is 6.7%, with all leads remaining within the ±10% band. Bars near zero indicate that BMI does not explain the D1 genetic associations at these cardiac loci.

